# Statistical Inferences and Analysis based on the COVID-19 Data from the United States

**DOI:** 10.1101/2021.11.30.21267096

**Authors:** Nivedita Rethnakar

## Abstract

This paper investigates the mortality statistics of the COVID-19 pandemic from the United States perspective. We bring out several exciting and glaring aspects of the pandemic, otherwise hidden, using empirical data analysis and statistical inference tools. First, specific patterns seen in demographics such as race/ethnicity are analyzed qualitatively and quantitatively. We looked at the role of factors such as population density in mortality rates. A detailed study of the connections between COVID-19 and other respiratory diseases is also covered. Finally, we examine the temporal dynamics of the COVID-19 outbreak and vaccines’ stellar impact in controlling the pandemic. Statistical inference such as the ones gathered in this paper would be helpful for better scientific understanding, policy preparation, and thus adequately preparing, should a similar situation arise in the future.

## 1 Corona virus and COVID-19 Pandemic

A severe global crisis emerged at the start of the year 2020, in the form of a virus outbreak called the novel coronavirus 2019 disease, now widely known by the acronym COVID-19. COVID-19 is an infectious disease caused by the virus Severe Acute Respiratory Syndrome Coronavirus 2 (SARS-CoV-2)^1^^1^. Believed to have transitioned from animals to humans in 2019, the first reported human infection of COVID-19 came up in Wuhan, a region in central China, on December 1, 2019^3–5^. Since then, the virus has traveled to different parts of the world and infected over 250 million people. On March 11, 2020, the World Health Organization (WHO) officially declared COVID-19 a pandemic^8^. In the preceding weeks and months, the virus swiftly spread and thus created a sense of alarm around the globe, which continues to haunt the world population to date. During this period, the pandemic has resulted in the loss of lives, upset many aspects of social dynamics, and caused unprecedented damages to the economy and livelihood of people around the world. As of 12 November, 2021, the COVID-19 pandemic consumed over 5 million lives around the globe. Table 1 shows a tabular summary of the COVID-19 global situation in terms of the aggregate number of infected cases and deaths^6^ (There likely is some under-reporting of the data from Africa^25^).

**Table 1.** Estimated COVID-19 cases and deaths: A global summary as of 12 November 2021. Source^6^.

| COVID-19 summary |  |  |
| --- | --- | --- |
| Geography | Cases | Deaths |
| Africa | 8,627,508 | 220,823 |
| Asia | 80,496,301 | 1,187,853 |
| Europe | 67,642,656 | 1,346,080 |
| North America | 57,234,738 | 1,166,627 |
| South America | 38,633,222 | 1,174,470 |
| Oceania | 332,321 | 3,916 |
| World | 252,967,467 | 5,099,784 |
| United States | 47,701,670 | 780,833 |

COVID-19 is primarily a respiratory disease (like asthma, lung cancer, and pneumonia) affecting the lungs and the immune system. People with mild to moderate respiratory trouble may recover without needing special treatment or medical attention. However, specific infected individuals may develop a severe illness that will require medical intervention. While we are still in the learning process when it comes to uncovering all risk factors for the extreme form of COVID-19, it is apparent a weakened immune system can potentially accelerate the disease into severity. In particular, people with underlying medical conditions such as cancer, cardiovascular ailments, chronic respiratory disease, or diabetes are at increased risk of developing a severe illness^12–15^. Similarly, the older populace tends to be more vulnerable in illness progression, leading to fatal situations^9,10^.

The landscape of COVID-19 is diverse and complex. Certain aspects of the disease and its functioning mechanisms are still under active research. Moreover, unlike most standardized conditions, a perplexing attribute of this pandemic is the variation in symptoms that is seen among the affected individuals. While some people get through with minor invisible signs, others – even relatively young and otherwise healthy subjects – get highly sick and sometimes border on death^16^.

Overall, the pattern of the coronavirus pandemic has been a series of waves. Each wave identified with surges of infected cases and deaths. Around the world, the heave in cases and deaths has varied across geography, as evident in Figure 1. In the U.S context, the country witnessed four distinct waves, as explicitly shown in Figure 2. At its height, the biggest wave, which took place between October 2020 through April 2021, was consuming nearly 3500 lives in a given day and a weekly toll accounting for as many as 26000 deaths.

**Figure 1.**
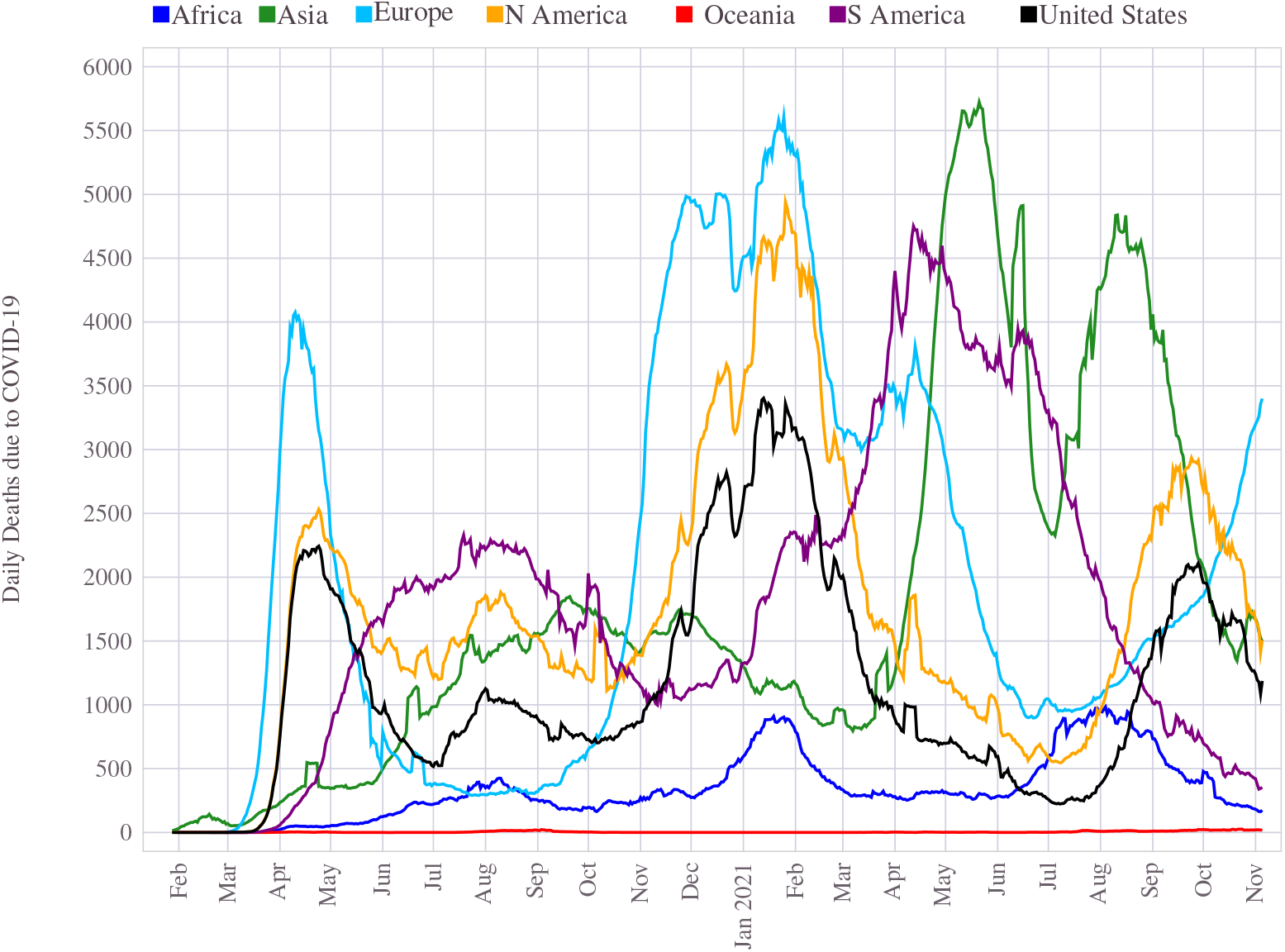
The coronavirus pandemic in different continents. The timing of surge and the peak number of cases and deaths have varied around the globe.

**Figure 2.**
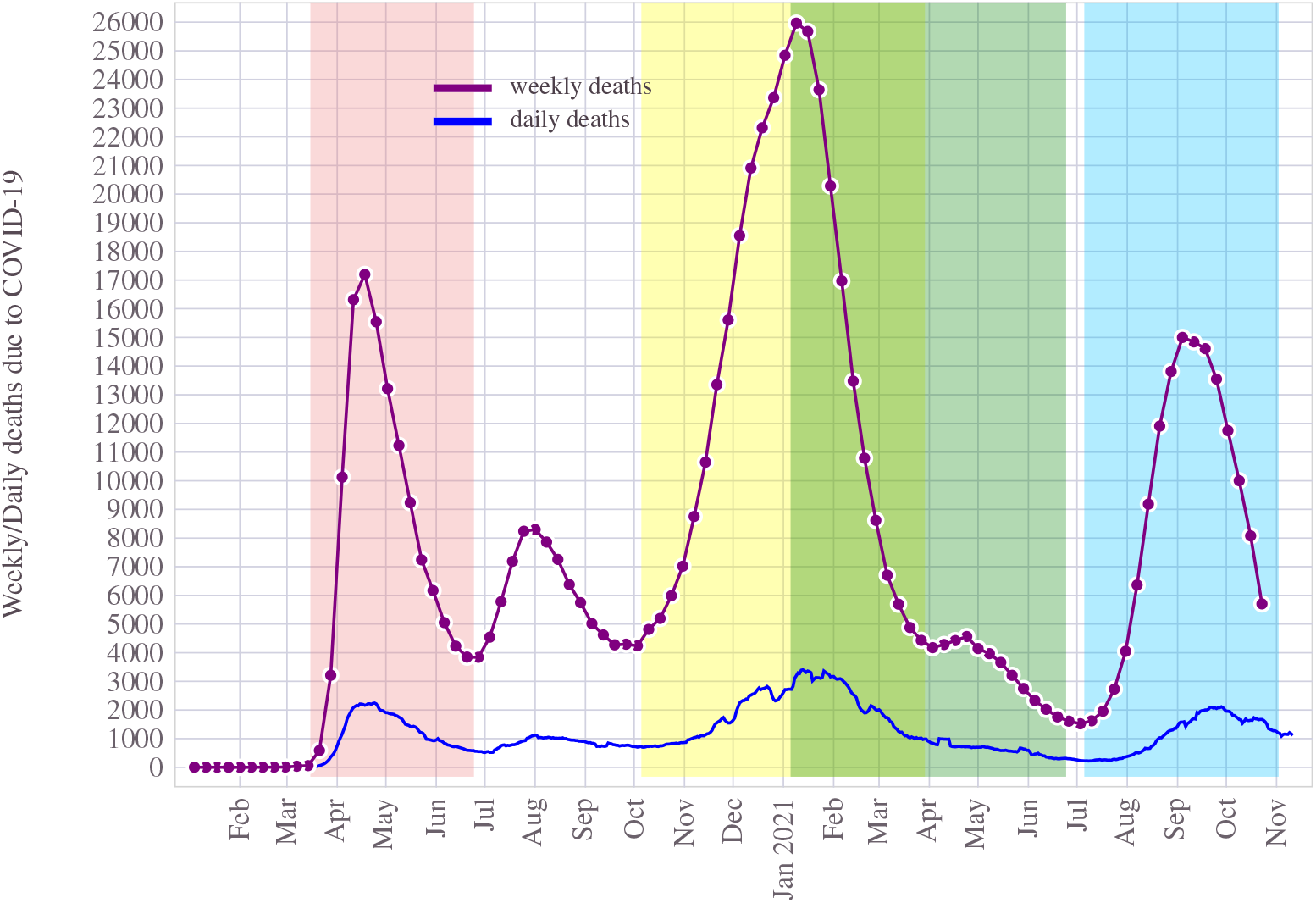
Covid deaths in U.S: Weekly and Daily deaths are shown against the duration of the COVID-19 pandemic (to date). Each color band demarks a specific timeline of interest, such as surge in deaths (i.e., wave). The period between Jan-July 2021 is specifically shown to highlight the impact of vaccines.

A significant aspect of COVID-19 is the changing dynamics of the virus itself, driven by its genetic mutations. During the pandemic, several new variants emerged in different parts of the world, at different epochs, some of which are classified as more damaging and threatening than others. In particular, the delta variant that emerged in 2021 has caused a severe impact in Europe and the United States.

For much of the early 2020, the world had to cope with the crisis without any medicines or cure. It was clear from the early stage that the most appropriate way to deal with the pandemic is through a vaccine. Active work on vaccine research and development has happened throughout 2020^18,41^. In December, the first vaccine approval occurred in the U.S. Access to vaccines to the public began in Jan 2021 in the U.S and other parts of the world, which helped control the pandemic to a manageable level.

Thanks to the internet infrastructure and increased support from institutions, more extensive societal participation, and cooperation between entities, the pandemic sparked a wave of research, data collection, and sharing. The availability of data and enhanced open access to information have greatly helped track, study, and analyze various disease dynamics. Some behavioral patterns, geographic and demographic trends, etc., could thus be gathered by analyzing such data, which in turn help further to support scientific understanding and aid in policy decisions to control and possibly prevent similar outbreaks in the future.

Much of the analysis done in this paper have relied heavily on publicly available data such as the one maintained by the Centers for Disease Control and Prevention(CDC)^26,27^, John Hopkins Coronavirus Resource Center^28,29^ and Our World in Data^7^. We outline the broader scope of the work discussed in this paper below.

## 2 Scope

In this work, we focus attention on the United States and assess the mortality situation triggered by the COVID-19. From the publicly available data, we gather, analyze and quantify some interesting correlations between the pandemic’s behavior and its impact on individuals across different demographics such as age, race/ethnicity, and gender. We further investigate how the pattern evolved geographically (over states in the U.S). Finally, we compare the pandemic trend to that of the population. Given that the pandemic is still active and has witnessed multiple waves over the last 20 months, we discuss some of the controlling factors and their role in its time evolution.

Identifying specific patterns emerging from data can be valuable in public policy decisions and mitigation efforts to prevent further impact on the disproportionately affected population. The lesson learned from this work may also act as a guideline to adequately prepare for future outbreaks of this nature.

Before looking at data specifics, we define some relevant metrics used for the analysis in this work.

### 2.1 Quantitative measure of mortality

The main emphasis of this paper is on mortality. Several mortality measures (sometimes referred to as mortality rate) are in place to quantify deaths. Broadly put, the mortality rate measures the frequency of occurrence of death in a defined population during a specified interval of interest. A similar metric is often used also for morbidity where the measured item is illness instead of death. The mathematical definition of the mortality rate *μ* of a defined population *P*, over a specified period Δ*T*, is given by,

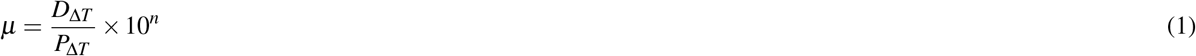

Where *D*_Δ*T*_ is the death count during the time interval Δ*T* and *P*_Δ*T*_ is the population size during the same time interval. Given that the population measure can vary slightly over moderate intervals, it is common to adopt the value at the midpoint of the measuring epoch. The factor 10^*n*^ is a normalization factor to help quantify the metric to a rounded decimal level than a fractional number. An example *n* = 1000 will translate the death count normalized over a subset of population with size equal to 1000 on the average.

Under the same metric framework, a few mortality measures are used in the context of an epidemic, such as COVID-19. Center for Disease Control and Prevention (CDC) outlines the definition of these metrics in^23^. For this investigation, we relied on two of the following metrics,

1. Cause-specific mortality ratio (CSR). This measure is the mortality rate from a specified cause (COVID-19 in our case) for a population. The numerator in Eq. 1 is the number of deaths attributed to a specific cause (COVID-19). The denominator remains the size of the population at the midpoint of the period.
2. Case to fatality ratio (CFR). This is the proportion of people with a particular condition (e.g., COVID-19) who die from that condition. It is a measure of the severity of the disease. The formula is adapted from Eq. 1 with the following changes: Numerator has the number of cause-specific (COVID-19) deaths and denominator accounts for the incident (or confirmed COVID-19) cases.

In this paper, we normalize CSR with a population size of *n* = 3, 2^*n*^ = 1000, which translate to *n* = 3, and define *μ*_CSR_ as.

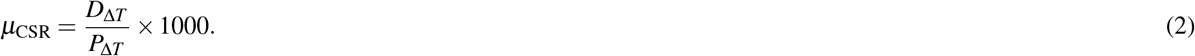

For CFR, *μ*_CFR_, we normalize the metric with *n* = 2, 2^*n*^ = 100 and thus express it as percent (%) throughout this document.

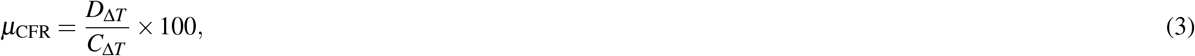

Where *C*_Δ*T*_ is the number of (infected) cases during the same measuring time interval Δ*T*.

The above metrics explicitly computed for an age group are sometimes called age-specific mortality rates. Therefore, when we study the behavior of the COVID-19 across age groups (see 4), we rely on such classifications. Likewise, when other demographics are analyzed, the same methodology is adopted.

CFR is easier to compute but is not a strict measure of mortality. The accuracy of these measures heavily depends on the exactness of the data accounting, recording, and availability. Even with this caveat, CFR offers valuable insights in estimating the severity of a pandemic such as COVID-19.

### 2.2 Outline of the paper

The rest of the paper is organized as follows: Begin with section 3 where it capture a high-level summary of the COVID-19 pandemic situation in the United States. Compared to the national level, specific trends that emerged at the state level are discussed in section 3.1. Details on age-specific trend in mortality is covered in section 4, where both the national and state-level data is compared. The influence that gender had in the COVID-19 statistics is the focus of section 5. Section 6 investigates details of how the pandemic disproportionately impacted specific ethnic communities.

The COVID-19 data with other respiratory illnesses such as Influenza and pneumonia are the focus in section 7. Virus genealogy is discussed next in section 8. The emergence of vaccines has been a vital factor in effectively controlling and containing the pandemic. Section 9 is dedicated to this topic. Section 10 gets into the temporal and dynamic nature of the pandemic. We end the paper with some concluding remarks and notes on aspects of reliably reproducible results presented in this work in Section 13.

There is a huge body work in the literature focusing on many important aspects of the pandemic. The economic impact is studied in several recent works including^77–79,81^. An up to date global database of research work done on the pandemic is maintained in by the World Health Organization^83^.

## 3 COVID-19 situation in United States

Having impacted nearly one-fifth of the national population, COVID-19 is officially marked as the deadliest pandemic in history for the United States of America, surpassing the 1918 Influenza outbreak, which had impacted more than a quarter of its overall population.

The first COVID-19 case in the United States was reported on 19 January 2020 in Washington state. A 35 year man, a Washington state resident, had recently returned from Wuhan, China. In the immediate aftermath of this event, the Seattle area became the epicenter of an early outbreak of COVID-19. The first official death in the United States due to COVID-19 is recorded in February 2020. Since early 2020, the virus quickly marched across the country, disrupting normalcy and forcing the country to a near shutdown.

United States is one of the most heavily impacted countries by the COVID-19 pandemic in terms of the aggregate volume of reported cases and the total number of deaths. The U.S also had the longest reign of the highest number of daily cases. As of 11 November 2021, there are 47.7 million (± 150K) people (approximately 14.5 percent of the total population) infected by the virus, among which 780833 ± 3400 reported dead (at an average case to fatality ratio of 1.62%). As per a report by CDC, for the year 2020, COVID-19 turned out to be the 3rd leading cause of death in the United States^30–32^. With these alarming death counts, the COVID-19 has officially dethroned the influenza pandemic of 1918 and 1919^2,33–35^, which consumed 675, 000 deaths. Figure 3 highlights the number of fatalities in each state in U.S.

**Figure 3.**
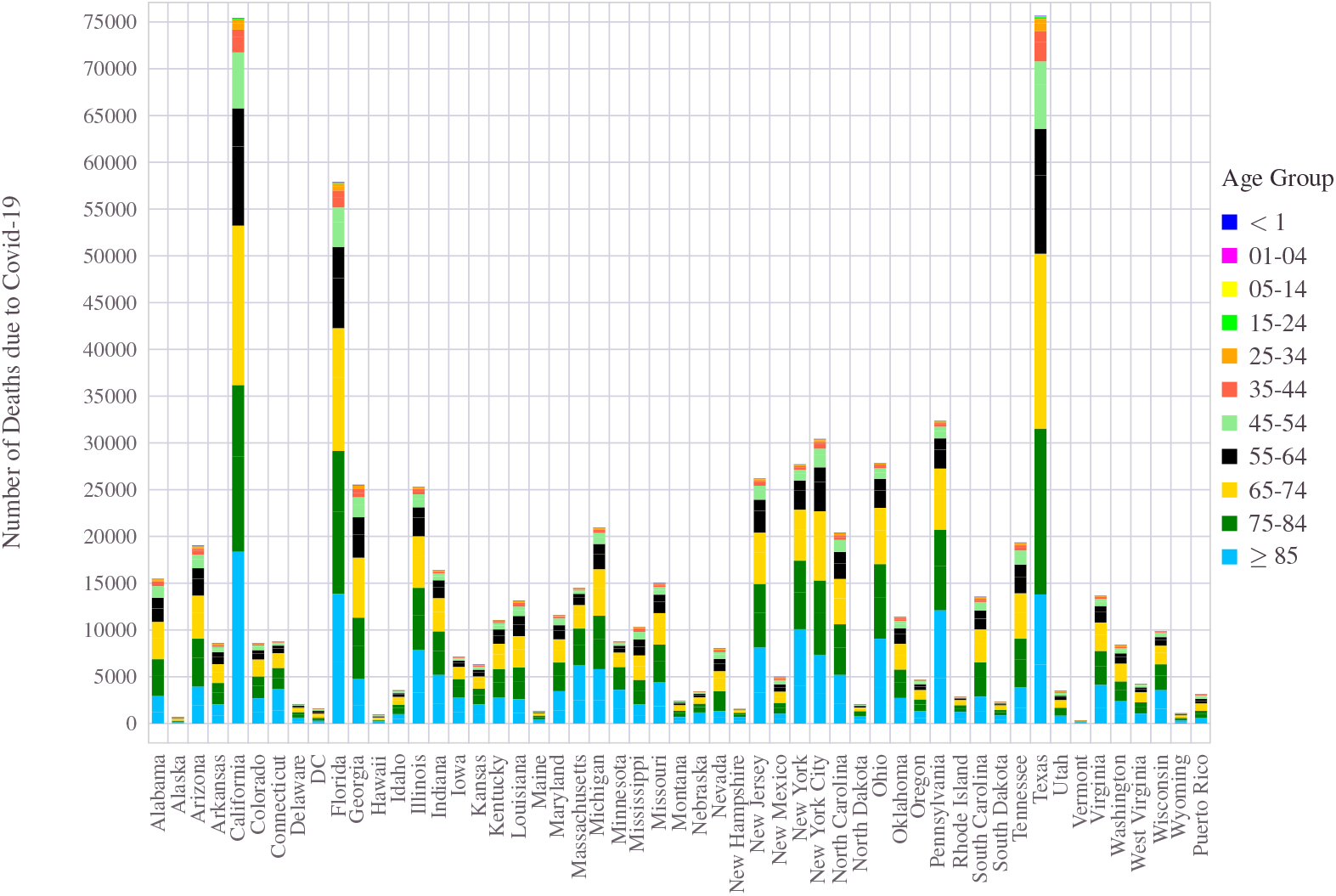
Number of deaths due to COVID-19 across U.S states (based on data as of 11 November 2021).

A timeline view of the coronavirus outbreak in the U.S is pictorially shown in Figure 2. It shows both the daily and weekly total number of deaths caused by the coronavirus in the country. Each color band indicates a specific surge (aka waves) in deaths (and cases) that the country witnessed during the pandemic. Since the first case reporting in February 2020, the surge in COVID-19 cases began in March 2020. When the WHO declared it a pandemic, the United States was already witnessing the first wave of COVID-19. Soon, a series of events followed, including sealing borders, enforced business shutdown, closure of schools and colleges, cancellation of sporting events, and enforcement of travel restrictions. The immediate aftermath of the outbreak also witnessed massive job losses and uncertainty. In addition, people began wearing masks and started practicing social distancing.

These measures made an impact that resulted in a drop in both infected cases and deaths during May 2020. However, experts emphasized the need to “flatten the curve.” A flattened curve reflects a reduction of cases on the graph of the cumulative number of COVID-19 cases over time^54,55^. This is highlighted by the first wave band, timed between March-July 2020 in Figure 2. Around this time, the states slowly began a phased reopening, hoping that the pandemic was nearing the recovery lane. Increased effort in understanding the disease and finding an effective treatment was picking momentum on the science front. In addition, some encouraging signs began emerging from different vaccine trials.

Relaxing the restrictions and partial opening resulted in the second wave during the summer months. This is reflected by the small hill in the in Figure 2 between July-October 2020. By September, schools were opened in a hybrid manner. Leading up to the 2020 presidential election in November 2020 and into the holiday season, the United States witnessed the most significant surge in COVID-19 cases. As cold weather drove people indoors, record-breaking daily cases/deaths were the norms for many days. As the holidays approached, the CDC urged people to stay home, limit the gatherings, and avoid mixing with people outside their households. The giant wave that stood between October 2020 and April 2021 in Figure 2 heralded this phase of the pandemic, its most deadly face so far. The period between July and November 2021 witnessed the so-called fourth leg of the pandemic surge, driven largely by the Delta variant.

A quantitative summary of the epic damage done by the pandemic across the United States is illustrated in Figure 4. Each state has the two-letter acronym and the four metrics; the total number of reported cases, deaths, CFR, and the percentage of the state population that the disease has infected. For example, for Montana (MT), 179770 individuals have been infected, and 2405 people have died due to the coronavirus disease. The number of infected people translates to 16.64% of the state population and the mortality rates corresponds to a CFR = 2405*/*179770 = 1.34%. The morbidity and mortality metrics discussed earlier in section 2.1, for each U.S state is shown in Figure 5.

**Figure 4.**
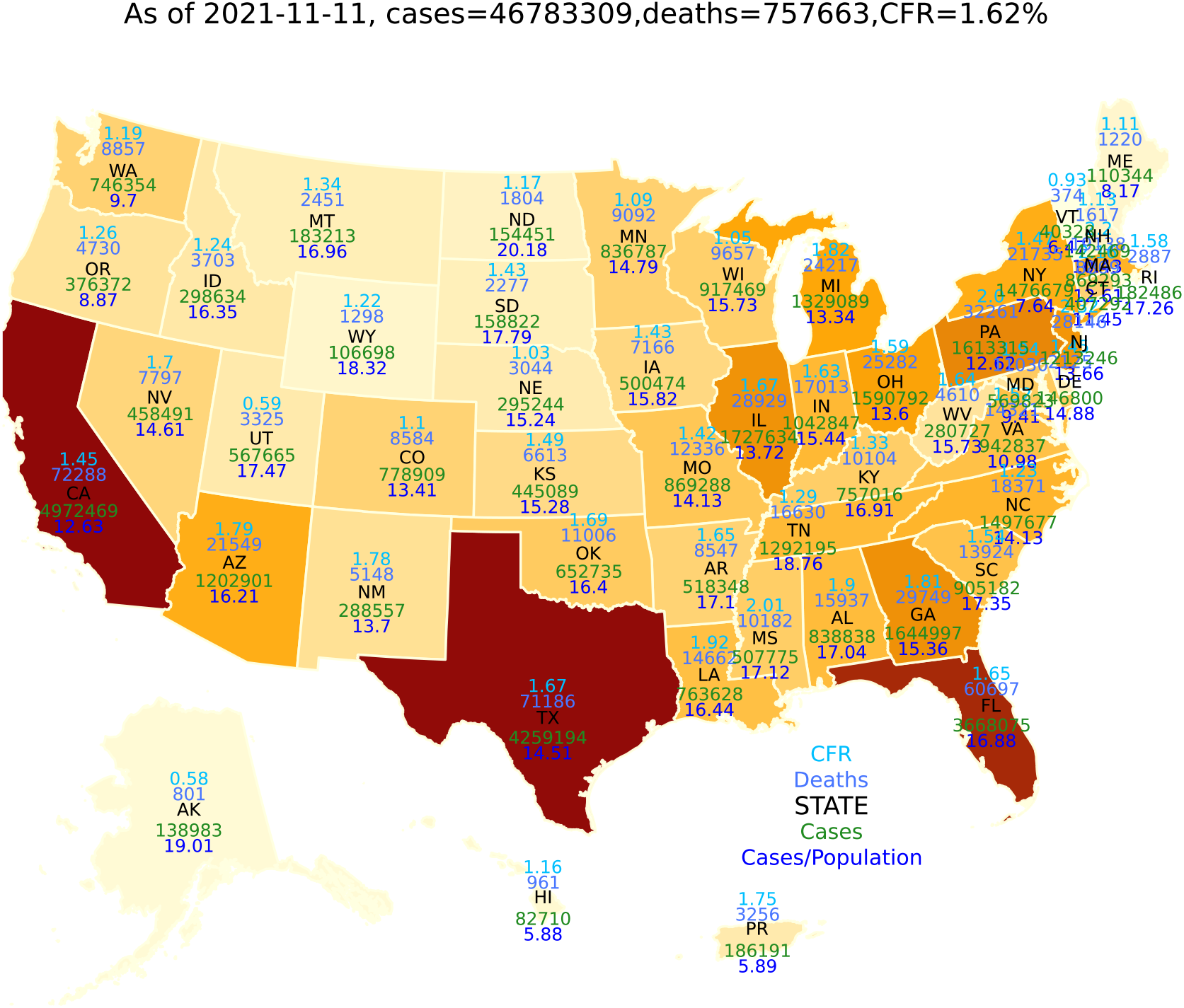
Covid-19 in USA: A summary of cases, deaths, CFR (%) and cases per capita (in percent) are shown on the state map.

**Figure 5.**
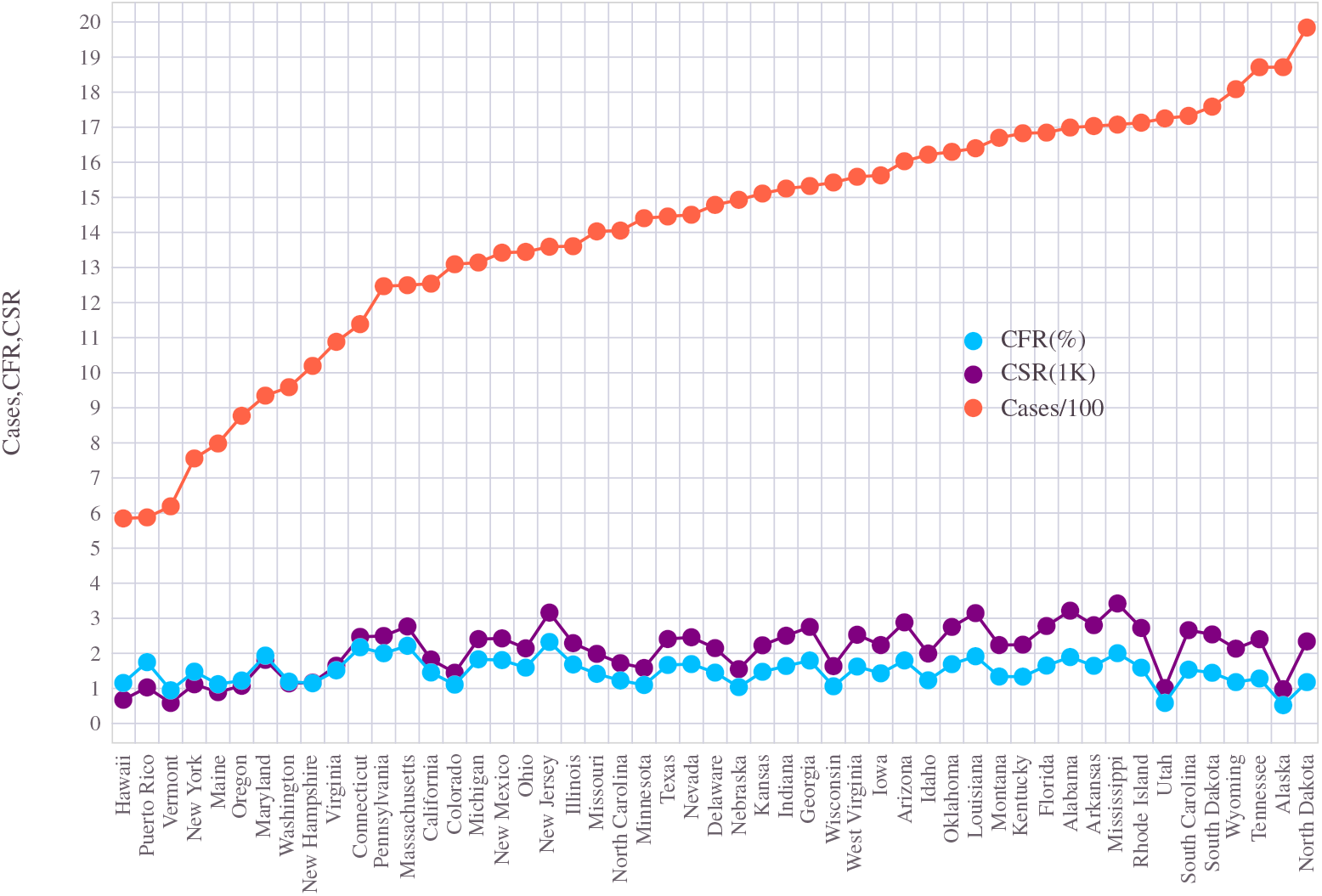
The mortality measures from the states of U.S. The morbidity(cases) and CFR are normalized per 100 people and hence expressed as % whereas the CSR is expressed per 1000 people.

Among the states (including the union territory District of Columbia (D.C)), the three large states California, Texas, and Florida, have altogether 13 million confirmed COVID-19 cases and a combined death count above 205, 000. California and Texas each reported more than 75, 000 deaths per state as of 8 November 2021. Although COVID-19 impacted every nuke and corner of the country, specific trends stayed the same across the vast geographical landscape. At the same time, certain variations on a few aspects of the pandemic also could be seen. New Jersey has been the state with the highest average CFR (averaged over all ages, gender, and ethnicity), CFR = 2.4%, and Alaska the lowest, with CFR = 0.5%. Approximately half of the states have an average CFR of 1.5% or higher (see Figure 5). The national average CFR is about 1.6%.

### 3.1 State specific patterns

Regarding the pandemic situation, from Figure 5, one can quickly infer that geographically, the infection rates have changed from state to state. The fraction of the state population impacted by COVID-19 has varied between 5% to 17.5%. Vermont registered a low 5% while Tennessee the highest Figure of 17.5%. Among the bigger states (in terms of population and land size), Florida saw an infection rate over 16% of its inhabitants.

The average mortality rates paint a somewhat different picture. Although many less infected states also ended up with lower CSR and CFR, no clear correlation exists between infection rate and mortality rates. Vermont remained the best state in terms of both infection rate and death rate per capita. It has an average of 1 death due to COVID per every 2000 inhabitants. The CFR = 1% for Vermont is also the lowest - 1 death per 100 infected cases - among all states. Both Massachusetts and New

Jersey both have reported a CFR over 2%. The per capita death figures (CSR) above 3 (per 1000 residents) came from New Jersey, Utah, and Mississippi. Looking at the age-wise statistics (see section 4), we will see how averaging over the entire set often ends up hiding the dominant demographic correlations. As we shall soon see in Figure 13, instead of using a single parameter, by incorporating multiple factors (such as age, population density), a reasonable correlation between the fatality rate and state demography can be established.

We will now look at various demographic trends and compare the national level metrics with the state-wise statistics.

## 4 Age and COVID-19 mortality

A clear trend visible in COVID-19 inflicted mortality is the way it has skewed towards older people. Nationwide, 28% of the death reported due to COVID-19 are more senior than 85 years. Over 90% of the COVID-19 deaths belonged to the age group 55 or higher. The national average age of people who died due to the pandemic is 55 years. The distribution of deaths has followed a monotonically increasing behavior with age. Among the deceased, people with age ≥ 55 accounted for more than 82% of total deaths in states throughout the U.S. In the northeastern part of the country, in New Hampshire, nearly all of the reported deaths due to COVID-19 are from this age group (See Figure 9).

The skew in mortality towards older age groups draws a stark contrast to how the population is distributed. The 2020 Census data show that the older people have accounted for a much lower population share. Despite occupying only a 15% share of the population, adults older than 55 years ended up a giant portion equalling 90% of deaths in the country. Individuals of age ≥ 75 had registered 55% of the total deaths, at a tune when this cohort garnered a mere 5% proportion of U.S inhabitants. This contrast is illustrated in Figure 6. While the bulk of the U.S. individuals are in the 5− 64 age bracket, this group only accounted for less than a quarter of the total deaths reported. On the positive side, despite occupying 2*/*3 of the United States population, the people younger than 25 years could afford to wither the COVID storm by bracing merely 0.5% of the overall COVID-19 death counts. In Figure 7, the complementary cumulative density function (CCDF) of the population is shown along with that of deaths. The CCDF is a statistical metric that computes the probability ℙ (*X* ≥ *x*) that the random variable *X* exceeds a threshold *x*^21^. Using the age group as a discrete random variable, we can compute the CCDF of the population distribution and that of the distribution of the deaths due to the pandemic. The green bar represents the CCDF of the population distribution, and the sky blue line (with circle marker) captures that of the COVID induced deaths, expressed as a percentage. A population CCDF value of 15 at age group 55 − 64 means 15% of the United States population are of age 55 or older, i.e., the probability ℙ (population whose age ≥ 55) = 0.15. For the same age threshold, the CCDF of the distribution of COVID deaths shows 76%, which means the probability ℙ (deceased whose age ≥ 55) = 0.76.

**Figure 6.**
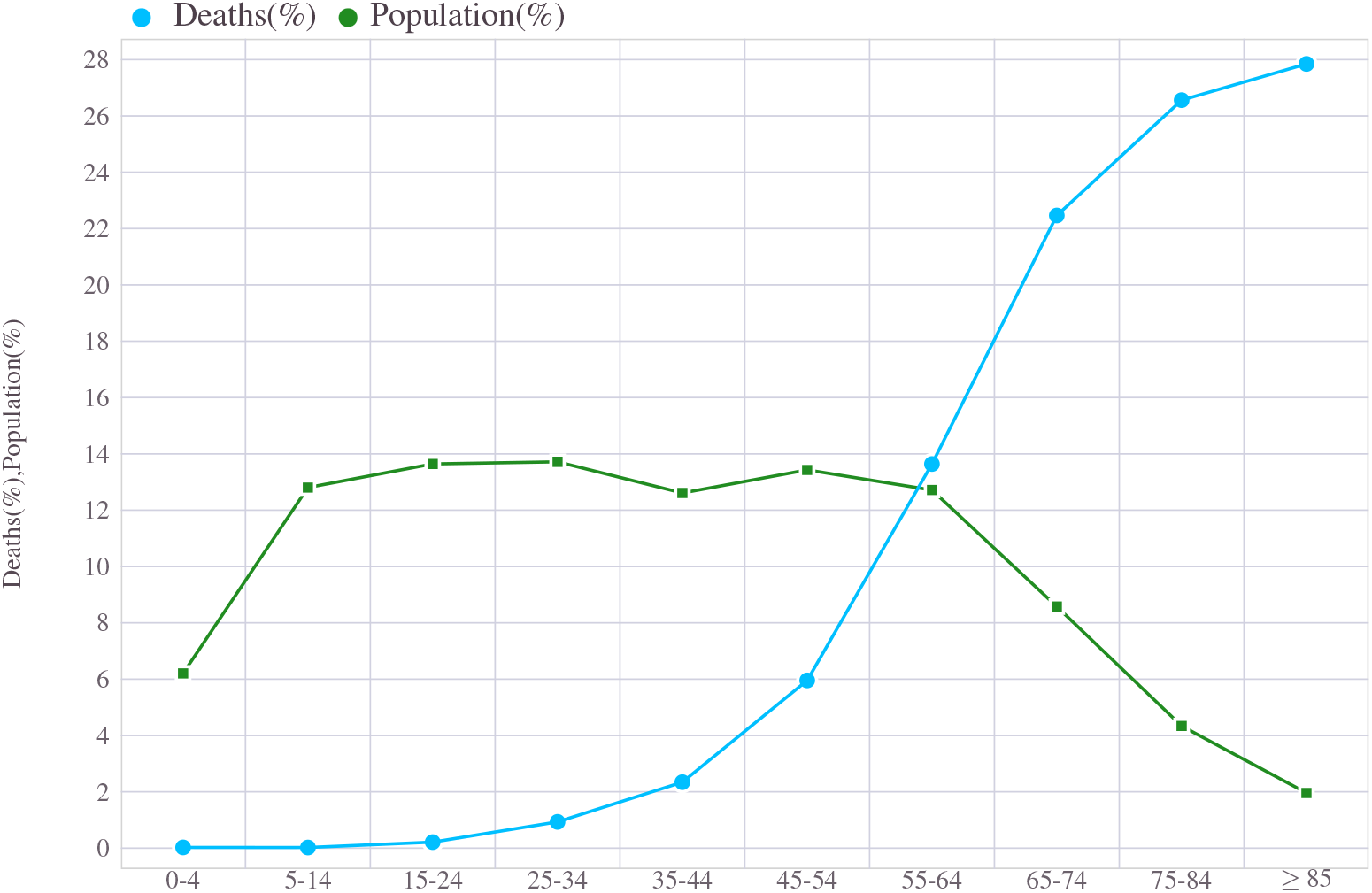
Age wise distribution of of total U.S deaths due to COVID-19 in comparison with the national population split across age groups. *x*-axis correspond to age of individuals.

**Figure 7.**
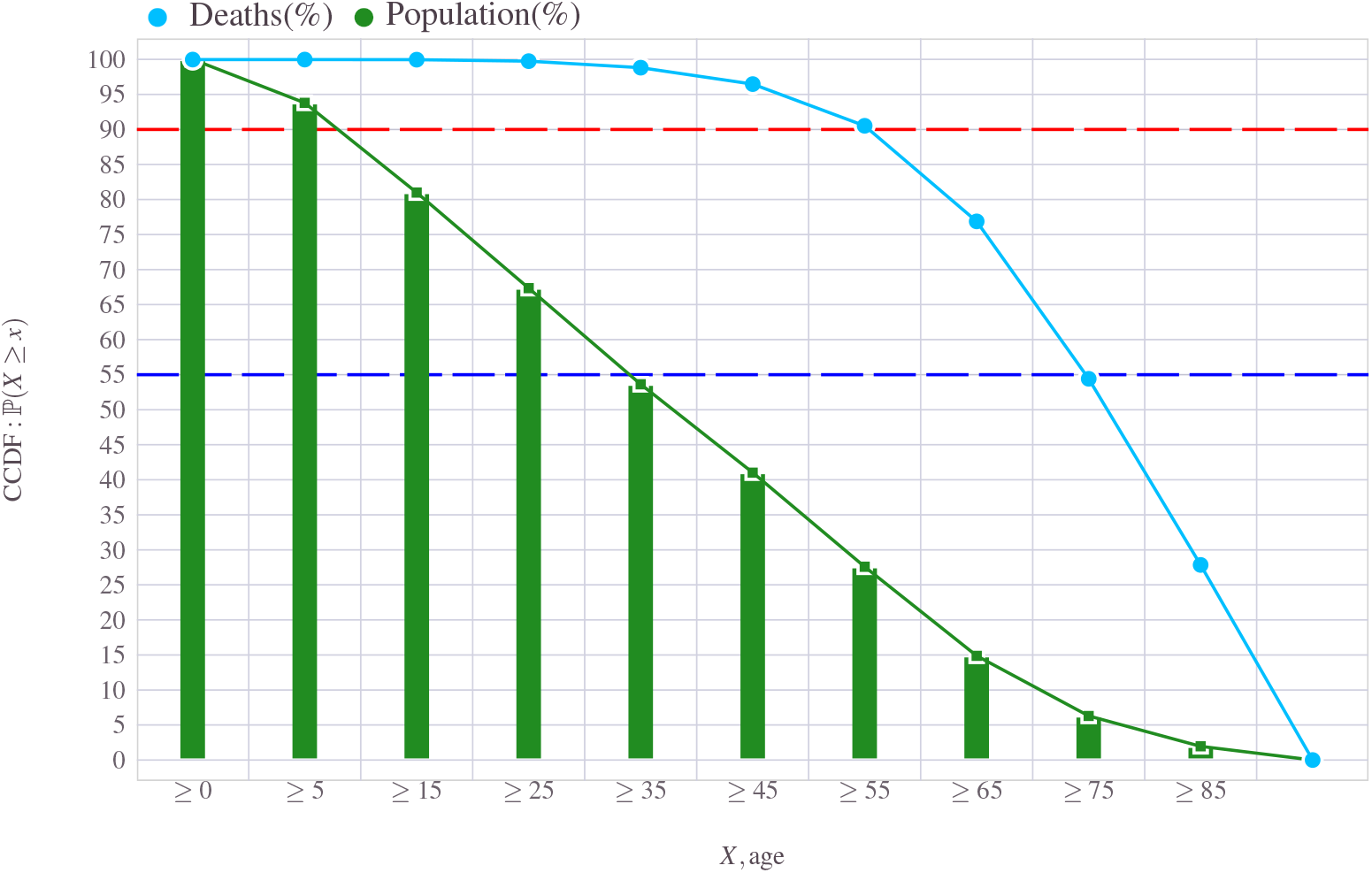
CCDF plot of Death and CCDF of Population, both distributed among age groups

Besides the distribution of deaths, the other two metrics we consider are case-to-mortality rate (CFR) and case-specific ratio (CSR). CFR and CSR per age group are shown in Figure 8. Recall that CFR is an average measure of how vulnerable infected people can be in eventually dying from the illness. In simple terms, this is the ratio of the reported number of deaths to the reported count of infections, usually expressed as a percentage. On the other hand, CSR is a pure count of the number of deaths normalized to 1000 people of the same demography (in this case, age group). As an example 30 deaths taken place from population of 10, 000 all belonging to a specific demographic section maps to CSR of *μ*_CSR_ = 30*/*10000 *×* 1000 = 3 (normalized per 1000). In the same demography if the disease had infected 2000 individuals, the corresponding CFR will be *μ*_CFR_ = 30*/*2000 *×* 100 = 1.5%. The key inference from the COVID dynamics seen from the United States point to a steep increase in the mortality figures towards the older segment of the U.S. society. All three of these metrics, split across age group is shown in Figure 8. Even though the normalization factor is different for CFR and CSR, a striking similarity of its distribution across age narrates the statistical consistency of age in governing the dynamics of the pandemic. The figure also reflects that the average metric such as CFR often hides the true impact of the age demography in the death counts. Even though the average CFR is 1.62 for the country, the true measure is almost 16 times higher than this average for the oldest segment of its society. Similarly, in the COVID-19 deaths, the lion’s share of the burden fell on older ones. In particular, the 85 and older people alone sliced 28% share of total deaths due to the disease.

**Figure 8.**
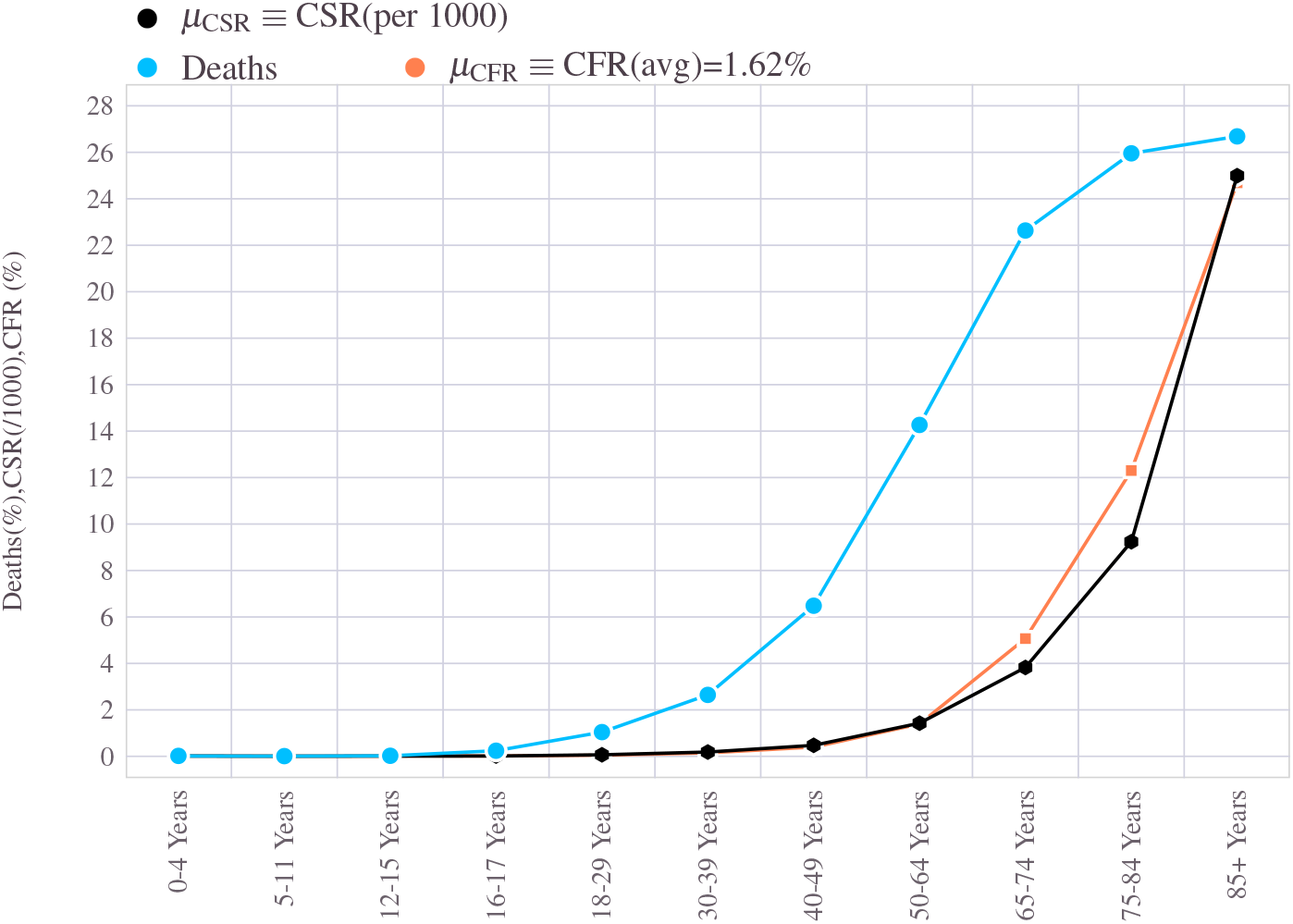
The COVID-19 mortality measures and the distribution of deaths across age groups. The *μ*_CFR_,*μ*_CSR_ are computed for each age group and plotted along with the way death count distribution among age cohort.

**Figure 9.**
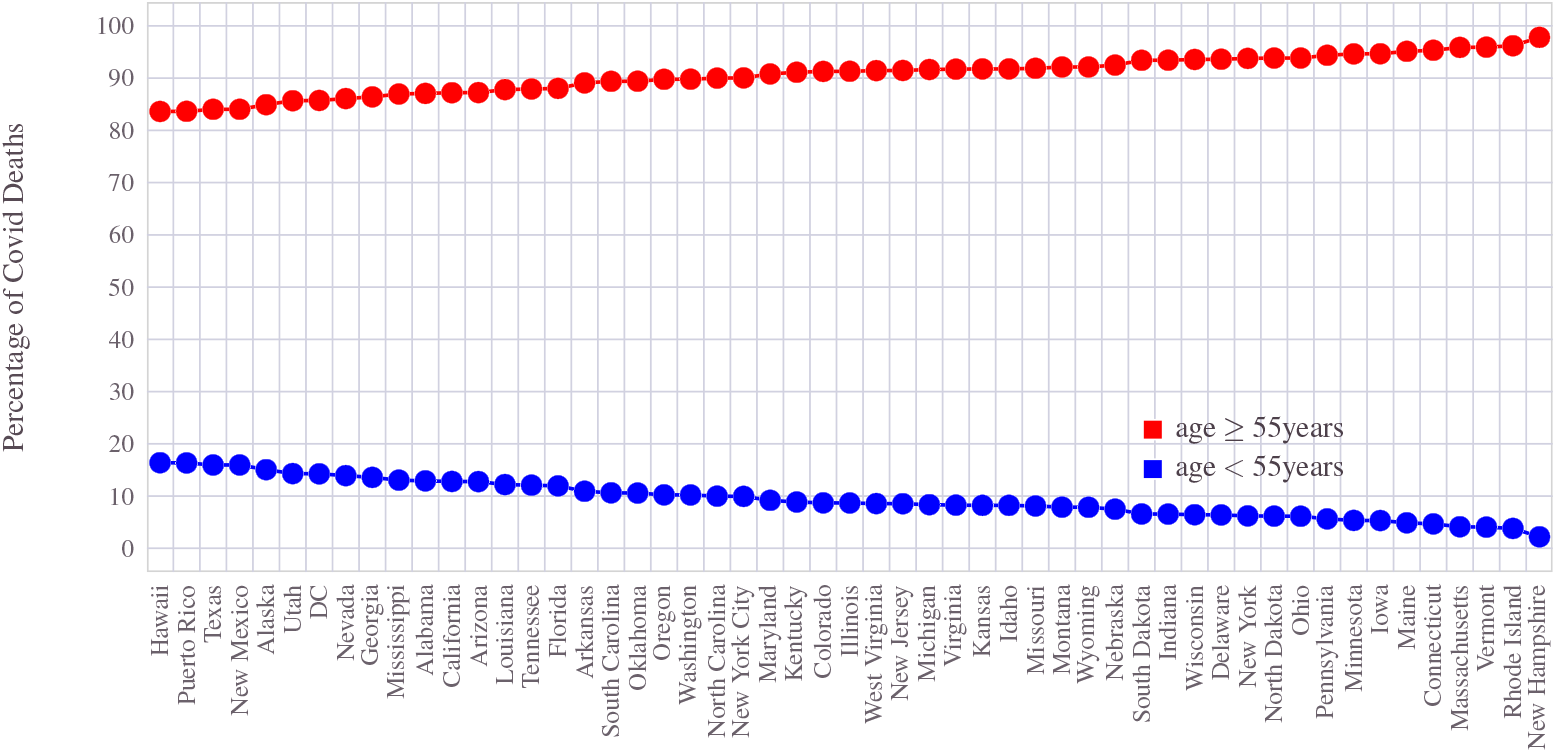
COVID-19 deaths in the U.S states when split into two groups based on an age threshold of 55 years: Overall, the older people accounted for 82 − 99 percent share of total deaths among individual states.

### 4.1 State wise trend on the impact of Age in COVID-19 mortality

The United States has a near equitable distribution of resources, albeit having minor variations between regions or among states depending on size and population. The state-wise population distribution across age groups is shown in Figure 10. By and large, the age-wise population demography fairly stays the same across the provinces. A few states such as Utah and Alaska have approximately 5% fewer older adults than most states (and the national average). On the other hand, states such as Maine, Vermont, and West Virginia possess a slightly higher percentage of the older population than the national average figures.

**Figure 10.**
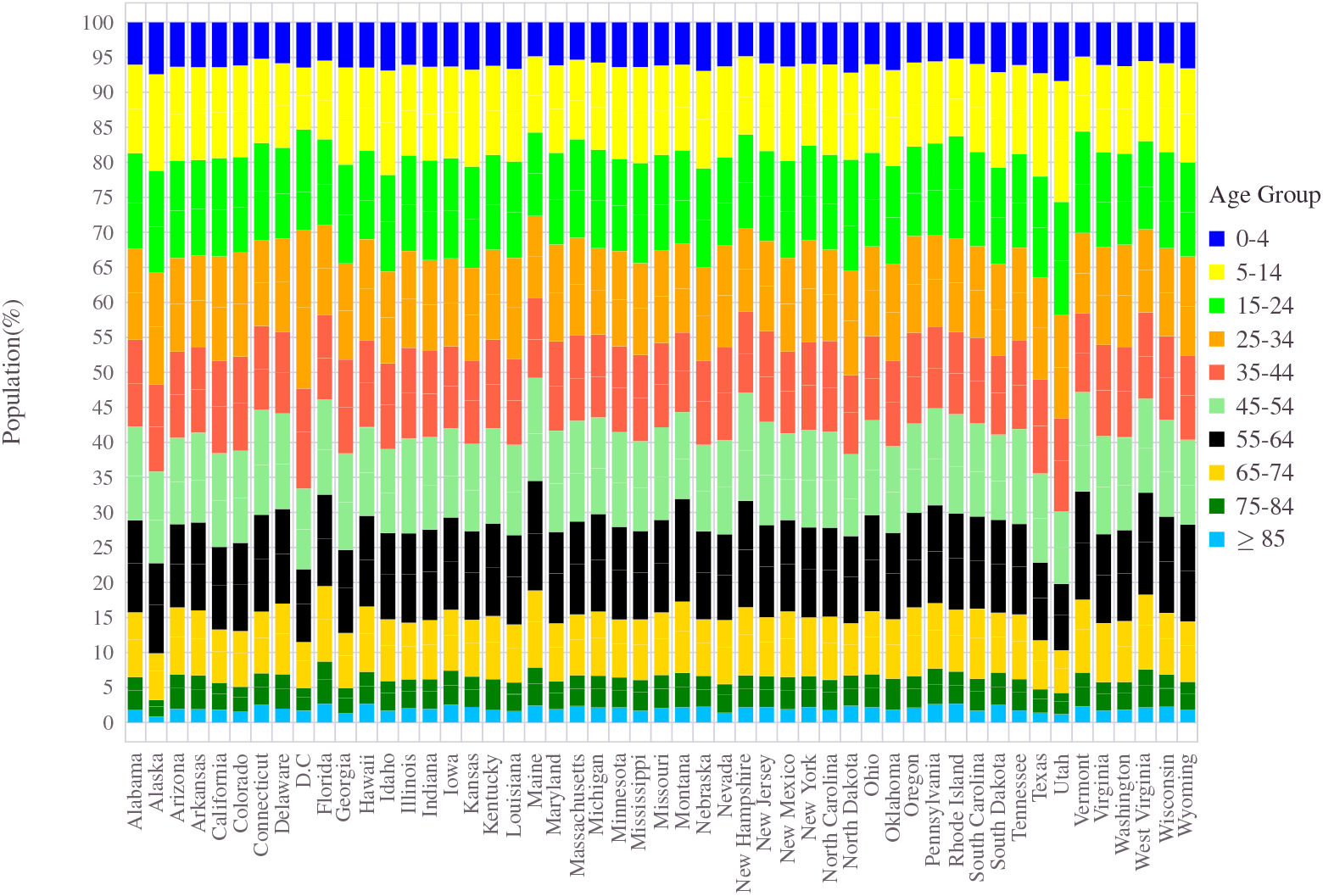
Age wise distribution of state population. For each U.S state, the population divided among age groups is expressed as a percentage.

In Figure 11 the age-wise split of deaths in various states is shown as stacked bar plot (The absolute percentage of each cohort is shown in the marker plot version Figure 12). The 2020 census data captured into different age brackets for every state is shown in Figure 10 for reference. Even though people of age 65 took the lion share of the death counts, the exact amount varied quite slightly across the states, as seen in Figure 11. For example, in some states such as Alabama, Alaska, Arizona, Nevada, Texas (and D.C and Puerto Rico), the percentage of deaths stayed approximately 20%. In contrast, that number is north of 40% in Connecticut, Iowa, Massachusetts, Michigan, New Hampshire, North Dakota, Pennsylvania, South Dakota, Utah, and Wisconsin. New Hampshire, in particular, has more than 3*/*4 deaths accounted by people who are older than 75 years old. A few states had a giant percentage count of deaths that belonged to people of age 85 and above. Connecticut, Iowa, Minnesota, Massachusetts, New Hampshire, North Dakota, Rhode Island, and Vermont are among those with more than 40% of deaths belonging to 85 or older. When it comes to the big states, Texas amounted to 45000 deaths, where 95% of them were 65 years or older.

**Figure 11.**
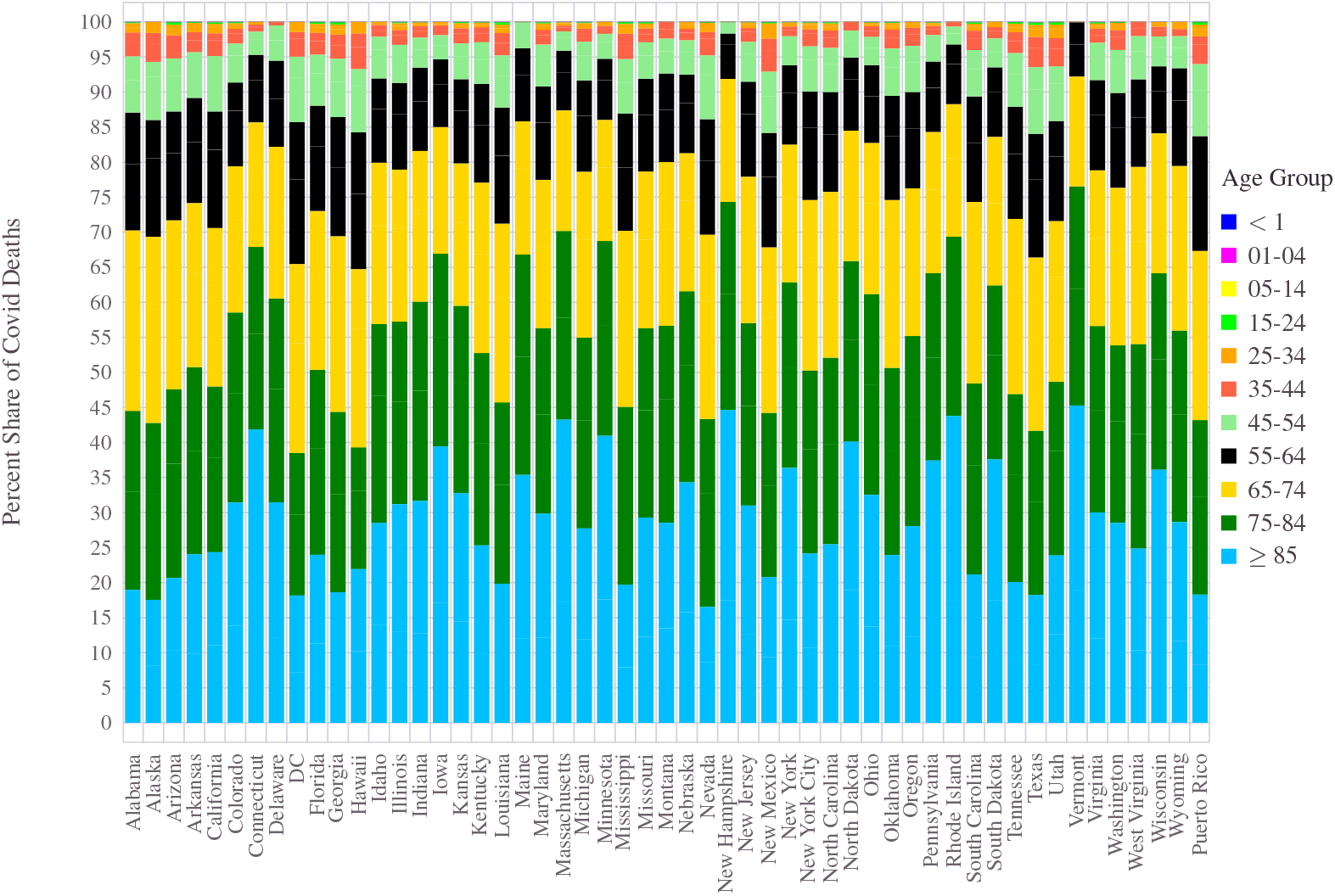
Age wise distribution of COVID-19 fatality figures in U.S states.

**Figure 12.**
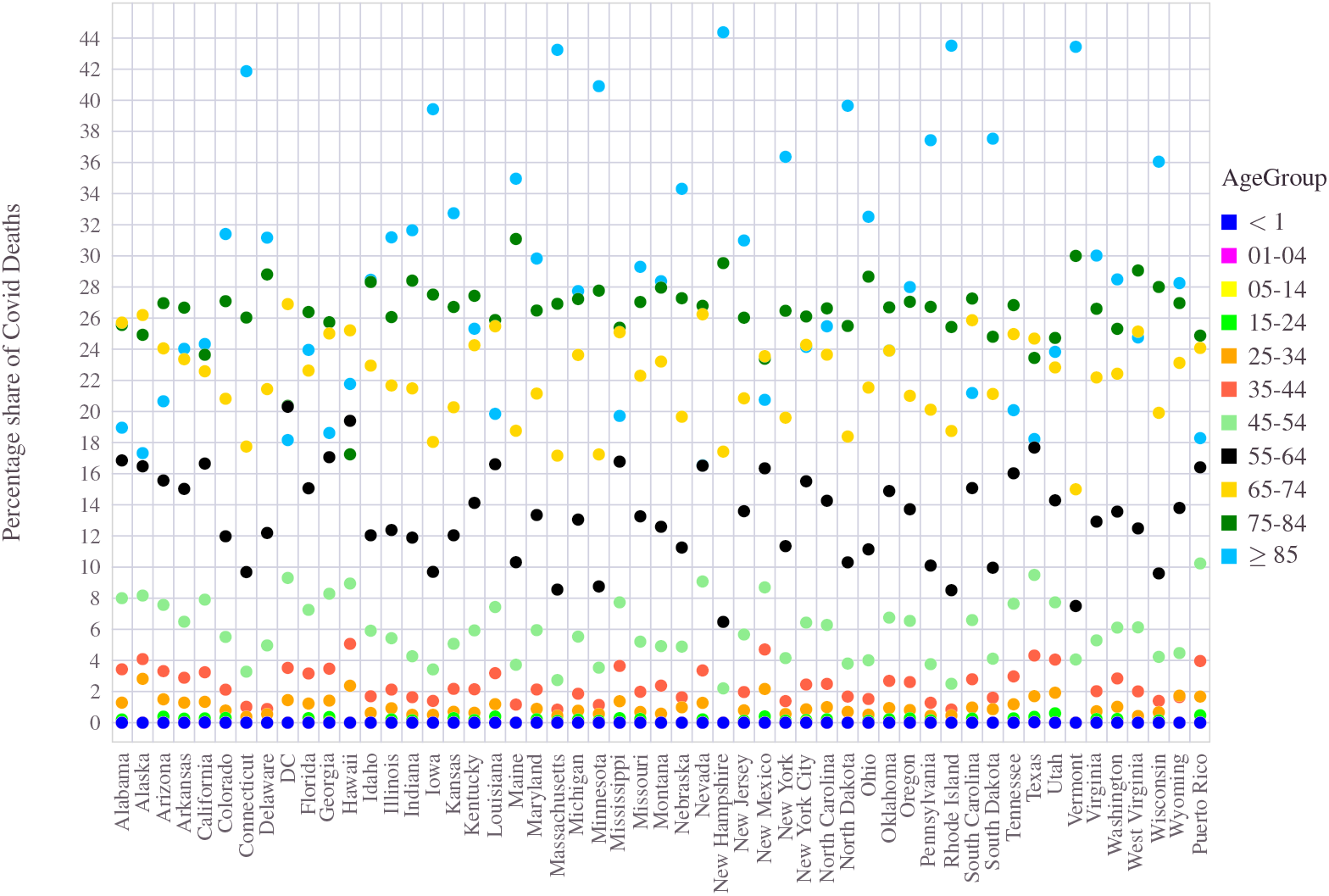
Share of COVID-19 deaths by Age group across the U.S states.

In terms of the absolute number of deaths, the three big states, California, Texas, and Florida, stood out. As of 10 November 2021, California led the pack with a record 76, 000 people succumbing to the pandemic. Going by the Census 2020, California has a total population of 39, 538, 223, which translates these figures to an average CSR *μ*_CSR_ 1.9 deaths per 1000 people. The state average CFR, *μ*_CFR_ = 1.45% for California is less than the U.S national average value 1.62%.

Stacking the mortality figures across the United States geography, one could draw some interesting connections with a few dominant factors which may have played a role in the mortality figures. Here we use CFR as the mortality metric to establish this relationship. Firstly, we can club the states into three different compartments. Those states with CFR *<* 1.4 have managed the situation relatively well, whereas those with CFR *>* 1.6 are classified as the worst hit. The list of states which fell in between (i.e., 1.4 ≤ CFR ≤ 1.6) are the intermediate ones.

A remarkable finding is that the states with relatively high population density (*ρ >* 170 people/sq.miles), a higher percentage of older age population (age ≥ 50), and a significant presence of Hispanic and Black population all have ended up with the high CFR values. The population density *ρ* expressed as the (average) number of people per square miles of land loosely describes the proximity between individuals in a given state/locality^57^. There are 28 states which met these criteria (*ρ >* 170 and with more than 12% of the population belonging to people aged 50 ≥ years) have found themselves with CFR *>* 1.6%. There are 20 states which ended up with CFR *<* 1.4, among which 19 of them had either low population density or less than 12% of the older population (aged 50 and above). Conditioning further with a combined population share of 27% or higher share of the Hispanic or Black population, we can see a strong correlation between these factors and high CFR value. There is, however, an exception to this rule in the form of North Carolina, which held a CFR = 1.2%, despite having a lower population density and a smaller than 17% of inhabitants who are 70 years or older. The depiction in Figure 13 uses the color codes with the orange-circle mark for the high CFR group of states and the green-circles for the least CFR. In the later section, when we look at vaccines’ impact on mortality, we will observe the drastic increase in death counts among states with poor vaccination rates, especially during the delta variant phase. These increased mortality rates from wave-4 have elevated the average CFR figures for Kansas, West Virginia, and Arkansas.

**Figure 13.**
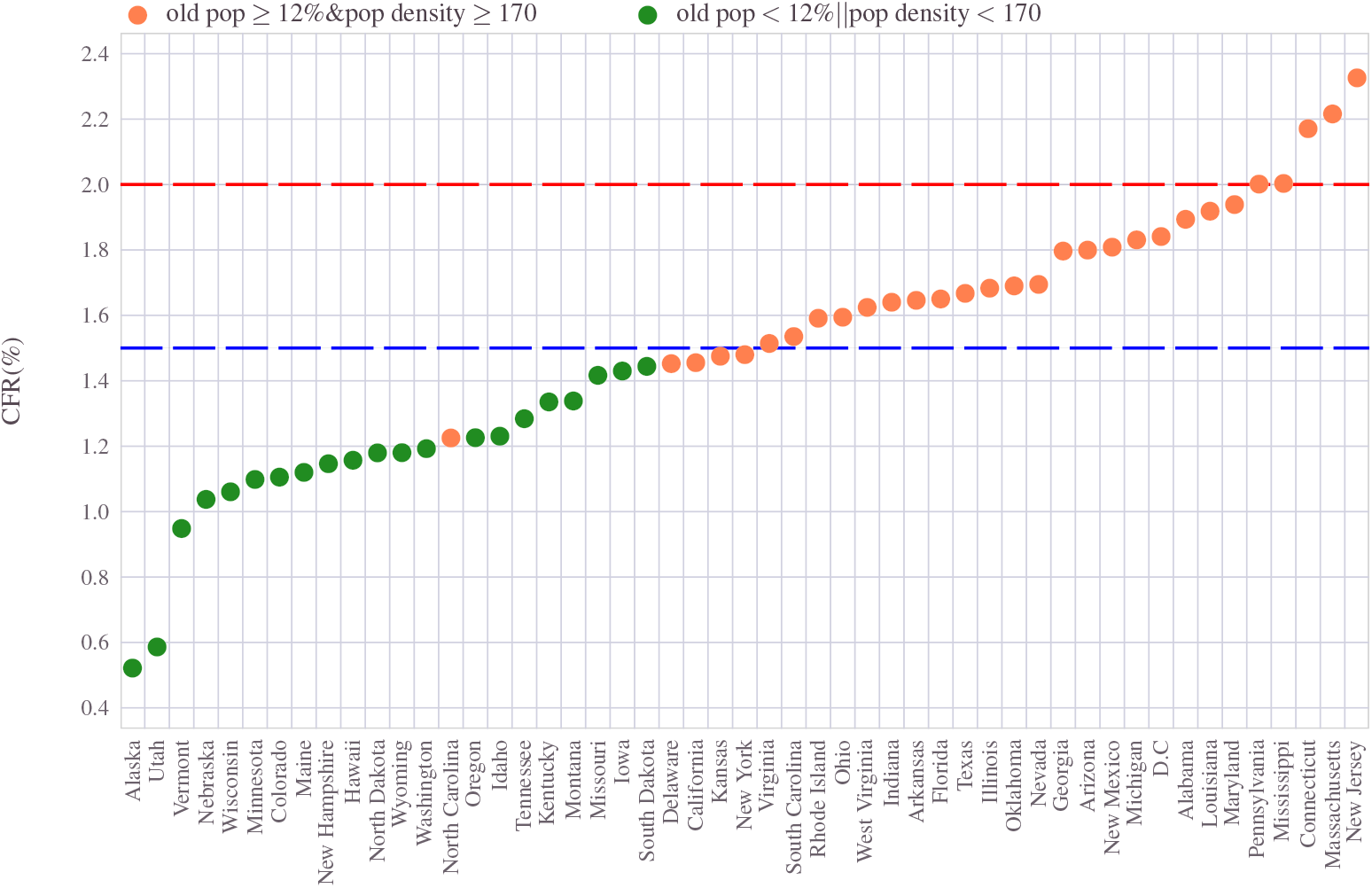
Correlation of mortality rate (CFR) with population density and old age population: States with higher population density and a relatively more significant percentage share of more senior age residents have higher mortality figures than those in sparsely populated states.

This connection is not entirely surprising. It reflects the vulnerability that older people had to deal with this pandemic. Because COVID-19 is a highly infectious disease, having a higher population density often results in an increased chance of interaction between people and thus a higher likelihood of getting infected and potentially succumbing to the illness.

## 5 Gender imbalance in COVID-19 mortality

We now look at the role played by gender or sex in the COVID mortality figures. Sex is a biological attribute linked to human anatomy and hormones that identify individuals as male, female, or others. Although it is linked to physical and biological traits, sex may rationalize an individual’s immune response to dealing with infections such as a virus. Despite having a broader spectrum of variations between individuals, publicly available data often classify them in binary as male or female. Therefore, we treat individuals as male or female in this paper. Although the term gender is a multi-dimensional social construct, which sometimes differs from the formal definition of sex, for this work, we loosely equate them to be the same and classify individuals whose gender is male or female. Thus sex and gender are used interchangeably throughout this paper.

The COVID-19 inflicted deaths separated by gender as a percentage share is depicted in Figure 15.

The demographic nature of the United States is such that there is near equitable share enjoyed by the male and female genders. A closer look at the state-wise data shows a slight advantage for females regarding the population share. Except for very few states such as Alaska, North Dakota, and Wyoming, the female: male ratio stays better than 1 : 1 throughout the country. The gender split of the population percentage across all states is presented in the Figure 14. Despite having equitable - or slightly higher females than males-gender distribution, the COVID-19 disproportionally burdened the males more than the females. Throughout the U.S, except Pennsylvania, where the numbers stacked almost equally between the two, the percentage of deaths remained higher for males than females. In states such as Hawaii, Nevada, and Utah, the male: female deaths reported numbers as high as 60 : 40.

**Figure 14.**
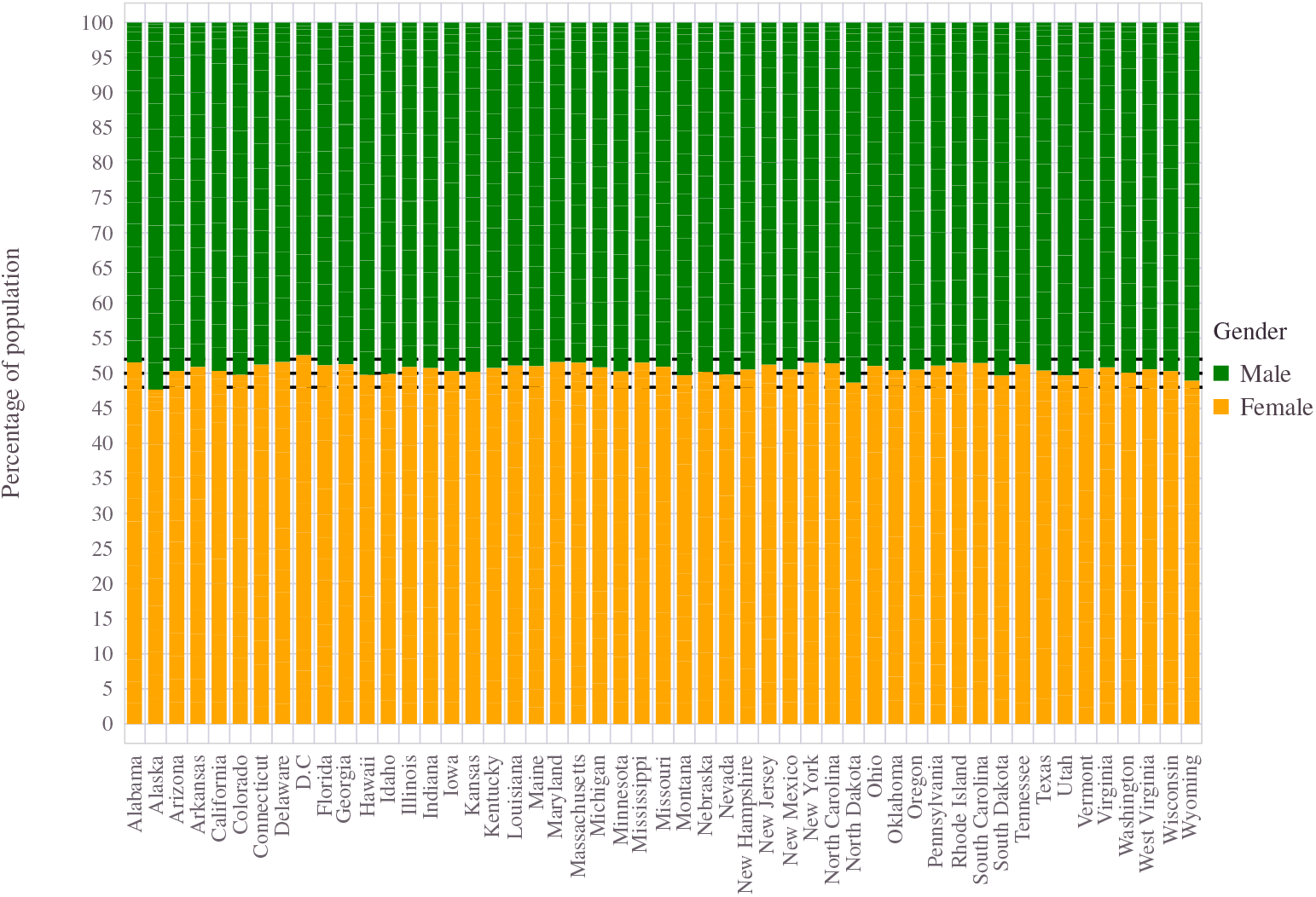
U.S state population share by gender.

**Figure 15.**
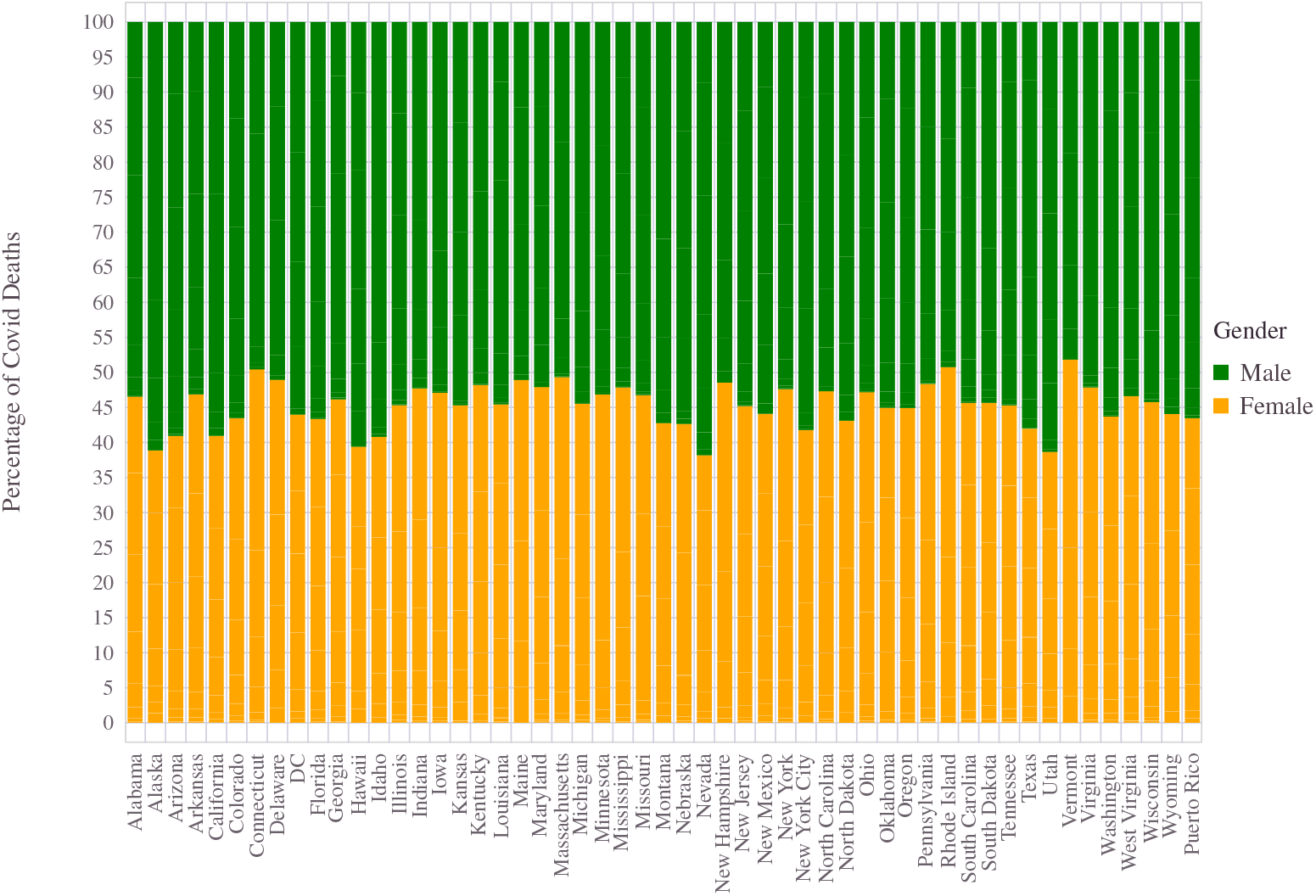
Gender wise percentage split of COVID-19 deaths across U.S states.

So far, when we considered gender, we were looking at men and women of all ages. Viewed agnostic to age, we have seen that among the deceased set, there were more males than females. There is, however, an interesting flip in this behavior when we classify the population into two groups who are on either side of age 85. Among the deceased who were 85 years or older, a significant portion of them were females. With this lone exception, all other age groups witnessed a higher number of male to female deaths. Instead of looking at the total deaths, it would widen the gender imbalance if we consider only those people of age more than 85 years. At the national level, in this age group (age *<* 85), males accounted for 42.0845% and females 28.6059% of total deaths. This gap of 13.4786% grew wider from the age agnostic male to female difference at the national level (The male to female ratio of deaths in the United States due to COVID is 54.6007− 45.3993), which is 54.6007−45.3993 = 9.2%. When it comes to the oldest age group (age ≥ 85), the scenario switched the trend from a net 12.5162 males to 16.7934 females. Two states, namely Nevada and Utah, stood as an exception to this rule (i.e., flipping the trend for 85 years), where the male: female ratio continued a similar trend across all age groups. The gender classification across all the states, illustrating the above-mentioned split behavior, is shown in Figure 16.

**Figure 16.**
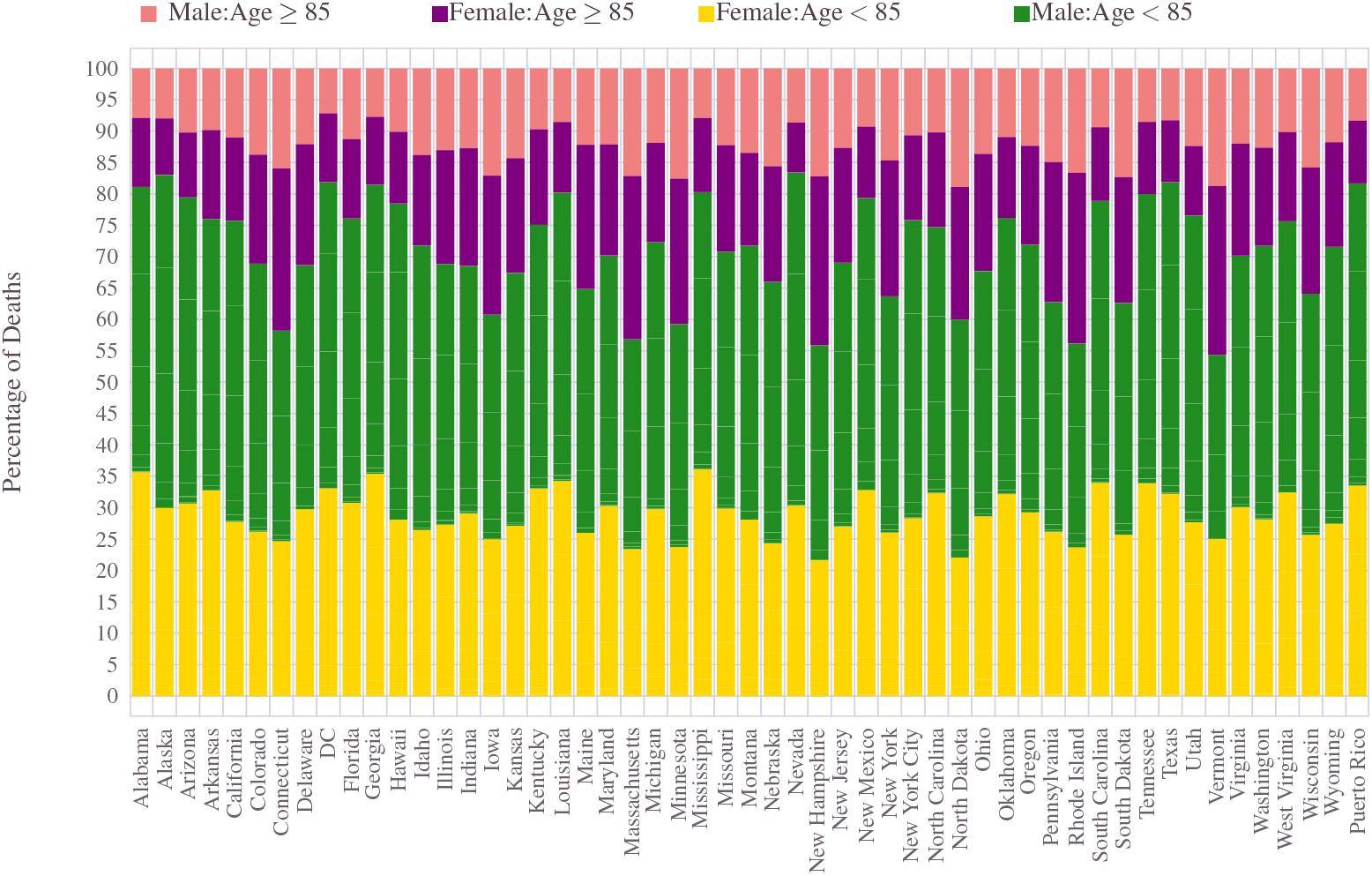
Gender impact: There is a flip in the behavior for 85 year and older versus those less than 85 years. Fatality statistics across different states of USA.

Once again, while the broader trend held across the states, the exact numbers varied considerably between geographical boundaries. For example, states like Connecticut, Iowa, Massachusetts, Minnesota, New Hampshire, North Dakota, Rhode Island, and Vermont had nearly 40% of deaths belonging to individuals aged 85 years or higher. Among this, about 63% are females compared to the 37%. So the states which had to deal with a larger share of deaths among the old age population also had to witness the loss of more females.

Although the mortality rate of COVID-19 infection is lower for women than men, several studies have shown that women across the spectrum have been disproportionately impacted in terms of psychological and physical distress. Some studies such as^39^ also suggest that women had to face higher levels of struggle when it comes to managing their careers around the COVID-19 pandemic, as compared to that men. These aspects are outside the scope of this paper, and we refer to references such as these^20, 22, 38, 39, 58^.

## 6 Ethnic divide

In terms of ethnicity and race, the population of the United States is very diverse. More than 40% of Americans identify themselves as people of color (Latino or Hispanic, Black, Asian American, Native Hawaiian or Pacific Islander, Native American, and two or more races). As per the official definition of the U.S Census, the population is classified into the following racial/ethnic groups.

1. Hispanic or Latino.
2. White alone non-Hispanic.
3. Black or African American alone non-Hispanic.
4. American Indian and Alaska Native alone non-Hispanic.
5. Asian alone non-Hispanic.
6. Native Hawaiian and Other Pacific Islander alone non-Hispanic.
7. Some Other Race alone non-Hispanic.
8. Multiracial non-Hispanic.

Here, White refers to those individuals whose ancestry can be traced back to any of the original peoples of Europe, the Middle East, or North Africa. The Black or African Americans are people whose genetic lineage can be tracked down to the Black racial groups of sub-Saharan Africa. American Indian and Alaska Native people are descendants of original peoples who lived in North and South America (including Central America) and maintain tribal affiliation or some form of community attachment with them. Asian is a collective name for people whose biological traits go back to the Far East’s original residents, Southeast Asia, or the Indian subcontinent. Finally, native Hawaiian and Other Pacific Islanders have ethnic connections to early inhabitants peoples of Hawaii, Guam, Samoa, or other Pacific Islands.

The term Hispanic or Latino identifies a Cuban, Mexican, Puerto Rican, South or Central American, or other Spanish culture or origin regardless of race.

Beyond the lists mentioned above, other ethnic groups are prevalent in the U.S, but based on the Census bureau’s assessment, further classifications did not substantially impact the overall findings. Furthermore, the CDC classified the U.S population along the same lines as Census while recording the COVID-19 data. The CDC nomenclature and the mapping to Census classifications (in parenthesis) are as follows:

1. Hispanic (i.e., Hispanic or Latino)
2. N-Hisp White (i.e., White alone non-Hispanic)
3. N-Hisp Black (i.e., Black or African American alone non-Hispanic)
4. N-Hisp Native Indian and Alaskan (i.e., American Indian and Alaska Native alone non-Hispanic)
5. N-Hisp Asian (i.e., Asian alone non-Hispanic)
6. N-Hisp Pacific Islander (i.e., Native Hawaiian and Other Pacific Islander alone non-Hispanic)
7. Other (All other races combined, including Some Other Race alone non-Hispanic and Multiracial non-Hispanic)

We will follow this shortened naming convention for the rest of the paper while discussing race/ethnicity.

Based on the latest estimates, U.S has a population of 332, 866, 152 (see^50^). According to the 2020 Census data, with an aggregate count of 204.3 million, the White population remains the largest race or ethnic group in the United States. Although in combination with other groups, 235 million people have identified themselves as White, going further by Hispanic and Non-Hispanic classifications, about 204 million among them is Non-Hispanic Whites. The Hispanic (this also considers the Latino population), which assumes people of any race, totaled 62 million in 2020. The Black or African American population tallies to 46.9 million, out of which the Non-Hispanic Black people alone is 41.25 million^51^. The Non-Hispanic Black group covers both the single-race Black and multi-race (excluding Hispanic) Black people. The next largest racial group identified is the Asian (alone or in combination) with 24 million. The American Indian and Alaska Native aggregated to 9.7 million in all, whereas the Native Hawaiian and Other Pacific Islander to 1.6 million.

The relative standing of ethnic cohorts, in U.S population is shown in Figure 17. Nearly 60% of individuals are Non-Hispanic White. The Hispanic community comes second with a 17% stake. The Non-Hispanic Black and Non-Hispanic Asian race occupy 13% and 6% share of the population. The remaining cohort altogether sums to less than 2%, which is factored further among Non-Hispanic Indians, Alaskan natives, pacific islanders, and others.

**Figure 17.**
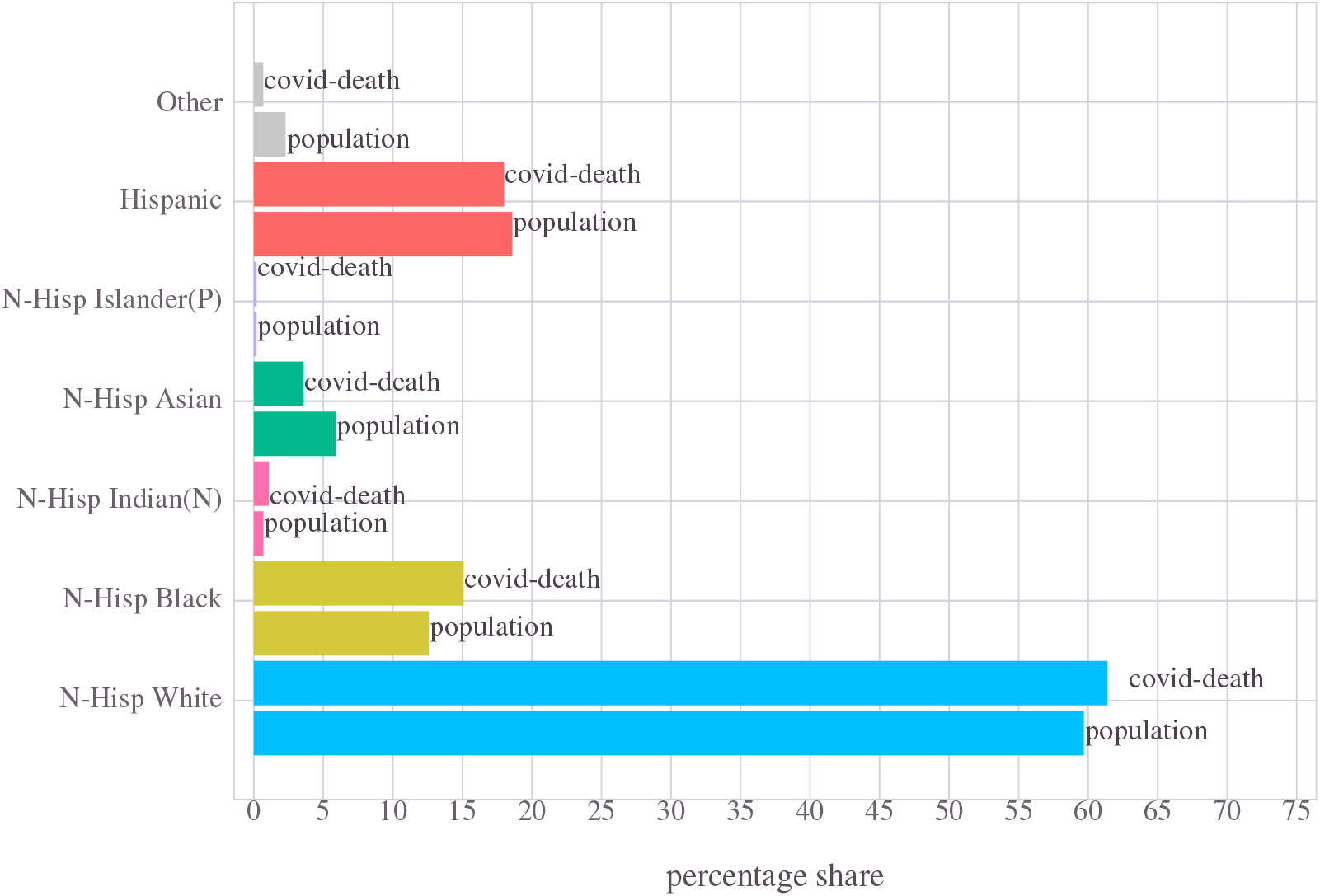
The distribution of deaths due to COVID-19 and U.S population, based on race/ethnicity. Each color code identifies with a given ethnic group.

The total number of people deceased due to the pandemic, categorized by their ethnicity, placed alongside the U.S population distribution is depicted in Figure 17. Out of the 736000 deaths (as of October 23, 2021), nearly 62% came from Non-Hispanic Whites. The next highest casualty fell on the Hispanic community, accounting for 18% of the total U.S deaths. They are followed by the Non-Hispanic Black, aggregating 15% share of the total people who have fallen to the pandemic. Out of the remaining 5%, the Non-Hispanic Asians emerged as the majority shareholder (effectively 3% of the U.S deaths), leaving the rest to the other ethnic groups, including the Indians and the Pacific Islanders. This is depicted in Figure 17.

Comparing the ethnic divide reflected in the population to that of the pandemic triggered death figures, at first glance, it may appear that the COVID-19 mortality has fairly well correlated with the population statistics. The side-by-side comparison in Figure 17 is a visual affirmation of this argument. However, an important aspect of this match-up is that the underlying data in play has ignored all the other demographic parameters(e.g., age, gender). This way, the comparison drawn is such that, for the average age of the U.S residents, the ethnic divide observed on the COVID-19 death is more or less in sync with the population distribution. As we shall soon discover, this assessment is highly misleading.

Rather than treating the data from an ethnic perspective alone, it is helpful to approach it as a multi-dimensional statistical inference problem. For example, the data can be viewed as an object spanned by different dimensions such as race/ethnicity, age, and geography. Instead of taking a global approach, if we take a more granular approach and view data in two or three dimensions, a clear demographic pattern seems to surface. Therefore, we begin by considering two attributes (ethnicity and geography/states, followed by race/ethnicity and age) before taking up the joint three-dimensional (ethnicity, age, geography/states) analysis.

### 6.1 Ethnic divide across geography/state

Here we compare the ethnic composition in each state’s population and draw a comparison with the state level COVID-19 mortality rates. In a way, this effectively projects the data into the ethnicity and geography/states space by ignoring the age distinction. In other words, all age groups clubbed into one.

#### 6.1.1 Ethnic variation in population across states

Across the 50 states, the share of the population owned by ethnic groups considerably varied. The majority of states have Non-Hispanic Whites as the dominant ethnic group. In more than 50% of the states, they represent higher than 70% share of the population as Figure 18 illustrated.

**Figure 18.**
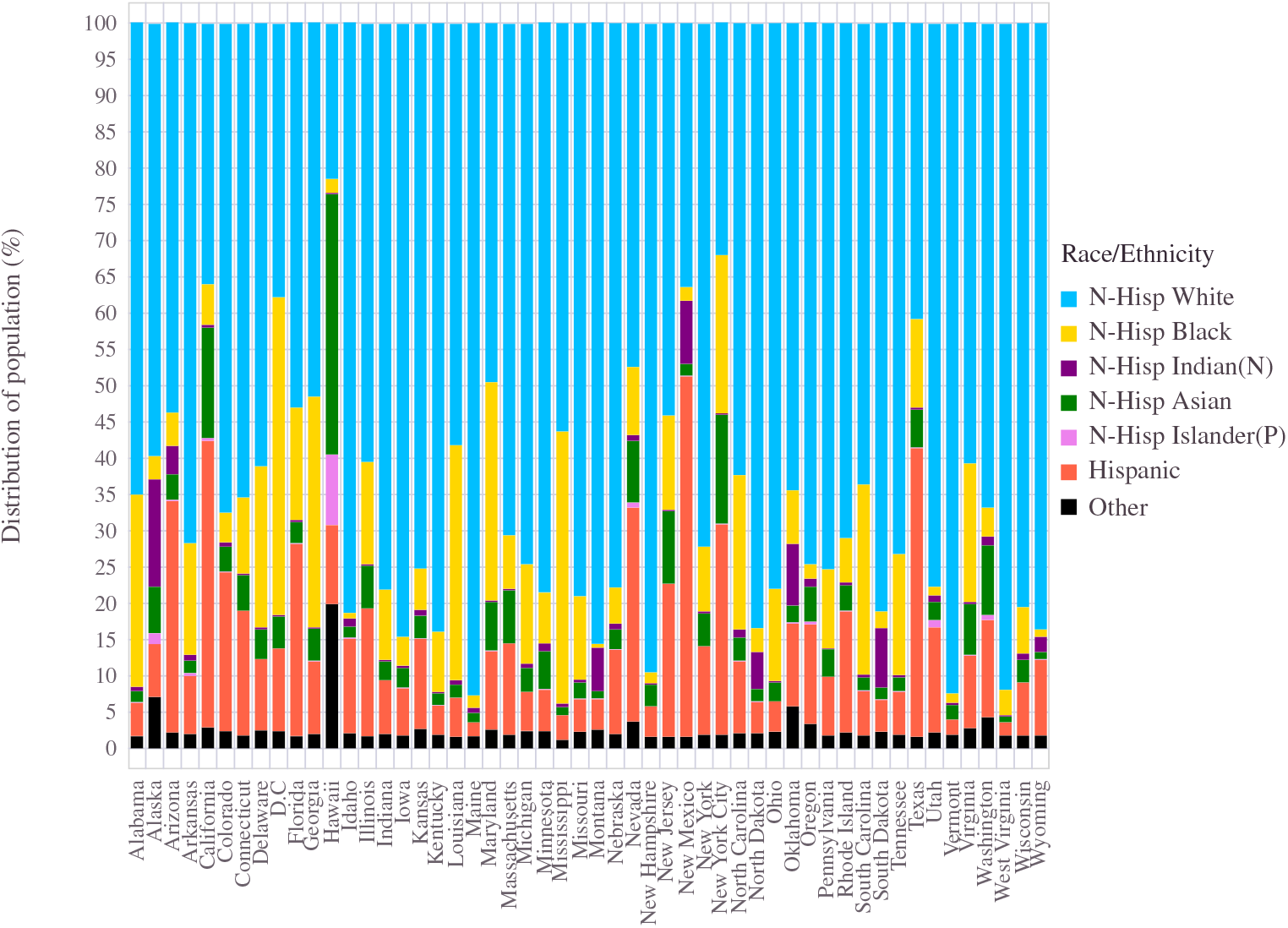
Census 2020: Distribution of U.S state population among ethnic groups.

States like Arizona, California, Florida, New Mexico, and Texas possess a significant Hispanic population share. States with notable presence of Non Hispanic black are Virginia (19%), North Carolina (21%), Delaware(22%), South Carolina (26%), Alabama (26.5%), Maryland(30%), Georgia (32%), Louisiana (32.5%),Mississippi (37.5%) and District of Columbia(44%).

Alaska has 15 percent of the Native Indian population, followed by South Dakota, Oklahoma, and New Mexico, each of which possesses a 9% Native Indian people.

The Southern states of the country are home to nearly 56% of the U.S Black population. The Midwest and Northeastern states each have 17% of the U.S black population, whereas the Western states only hold 10% of the Black population. The most significant number of Black people in the U.S. state is in Texas with 3.9 million. Florida (3.8 million) and Georgia (3.6 million) are the second and third in the top list of Black community presence. New York in the east has 3.4 million, and California in the west has 2.8 million black residents. These five states altogether hold 37% of the total U.S Black population.

The Hispanic population predominantly resides in California, Texas, Florida, Mountain West states (Arizona, Colorado, Idaho, Nevada, New Mexico, and Utah) and smaller pockets in the interior and eastern states. For example, new Mexico has 49% of its population identified to be Hispanic. Texas and California have approximately 39% share of Hispanic residents in the states. The remaining states with the highest percentage share of Hispanic presence are Arizona (31%), Nevada (29%), Florida (26%), Colorado (22%), New Jersey (20%), New York (19%), and Illinois (17%).

#### 6.1.2 Statewide COVID-19 deaths across Ethnic groups

A notable demographic contrast in pandemic mortality regarding race and ethnicity has been visible between regional locations in the United States. The proportional number of deaths due to COVID-19 across the U.S states is shown in Figure 19.

**Figure 19.**
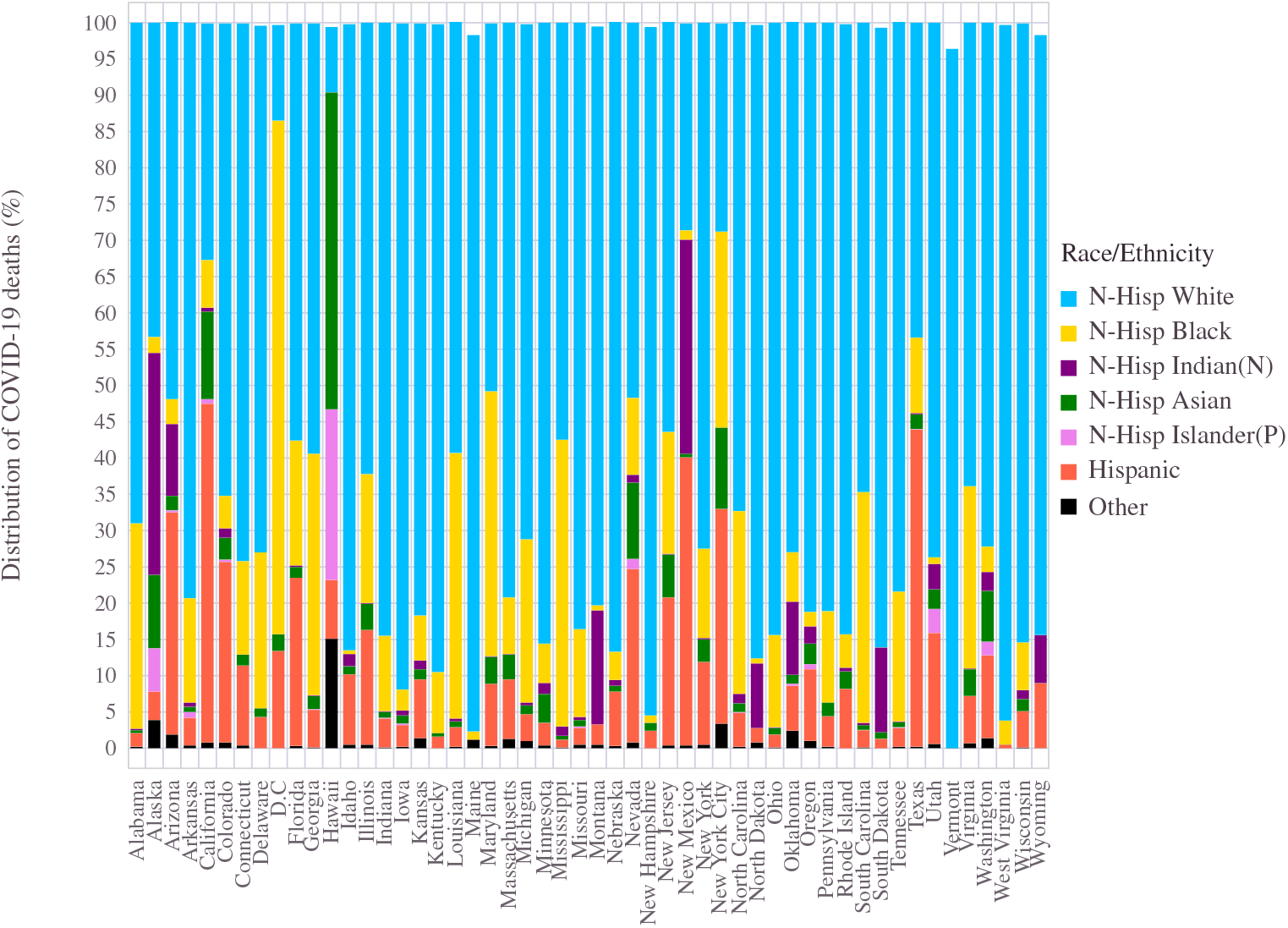
The distribution of deaths due to COVID-19 among different ethnic groups in each U.S state.

Arizona (30%), California (46%), Colorado (25%), D.C (15%), Florida (23%), Illinois (16%), Nevada(24%), New Jersey (20%), New Mexico (40%), Texas (44%) and Utah (15%) are the states which registered 15 or higher percentage share of the COVID-19 deaths reported by individuals from the Hispanic group. The people belonging to the Black community from the following states held 15% or higher percentage of COVID deaths. Alabama (28%), D.C (70%), Delaware (22%), Florida (18%), Georgia (33%), Illinois (18%), Louisiana (36%), Maryland (36%), Mississippi (40%), New Jersey (17%), North Carolina (25%), South Carolina (32%), Tennessee (18%), Virginia (25%). For the Native Indians, the community witnessed significant stress in states Alaska (30%), Montana(16%), and New Mexico (29%) in terms of the pandemic deaths.

More than half of the states saw more than 70% deaths coming from Non-Hispanic White men and women. Kentucky (90%), Iowa (91%), New Hampshire (95%), West Virginia (96%), Maine (96%), and Vermont (97%) had this number higher than 90%. A lower proportion (i.e., ≤ 30% of total deaths) of White individual deaths took place in Hawaii (9%), D.C (13%), New York City (29%) and New Mexico (29%). In California, 34% of the COVID triggered deaths belonged to the White community.

#### 6.1.2 State wise statistics against the U.S national average

The age agnostic, pandemic mortality data and its correlation with population distribution across the U.S geography are illustrated as a scatter plot in Figure 20. Each dot (different color indicates different ethnicity) displays the age-masked data from a state (i.e., total 7 dots for each state, one per each of the seven ethnic groups).

**Figure 20.**
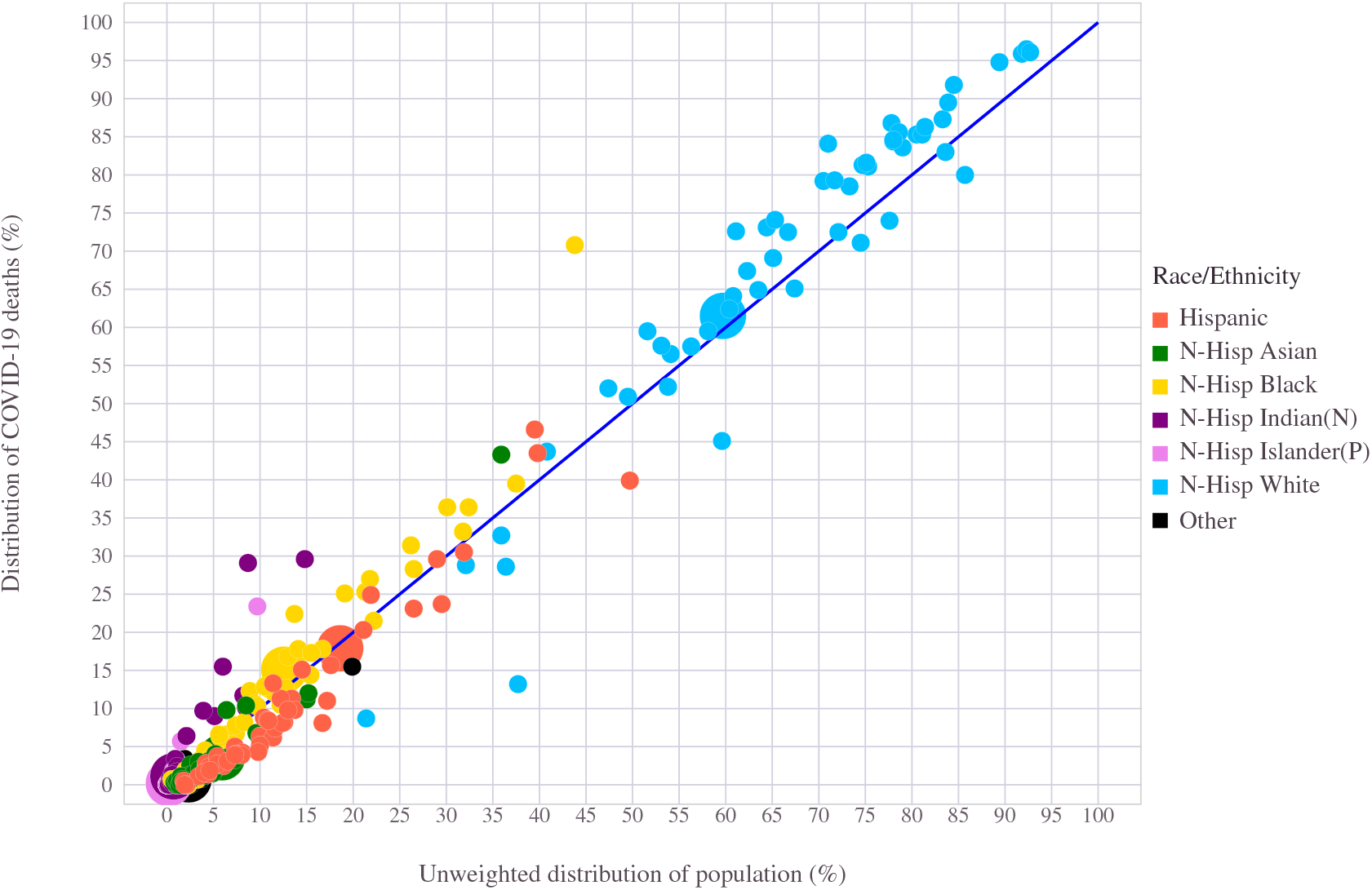
Total number of deaths due to COVID-19 versus aggregate population in United States: For each ethnic community, the percentage share of COVID-19 deaths in proportion to its population share is shown.

At the national level, the age ignored pandemic deaths correlate with how the population is divided into ethnic groups. The blue line is the reference line reflecting a perfect correlation between the two data sets. Although the absolute distribution has considerably varied across locations, the mortality and population remained fairly well correlated and thus stayed closer to the reference line. Although many states witnessed a severe loss of Non-Hispanic White communities, the normalized population statistics remained remarkably similar across all ethnic groups when age is not factored separately.

### 6.2 Ethnic divide across age groups

Here the U.S national level data is treated (i.e., skip the state level division and thereby drop the geographic/state dimension) as a two dimensional object. Each data has age (first dimension) and ethnicity (dimension 2) as a co-ordinate pair. For each age range (0 ≤ age ≤ 24 years, ≤ 25 age 34 ≥ years,35 ≤ age ≤ 44 years, 45 ≤ age ≤ 54 years,55 ≤ age ≤ 64 ≤ years, ≤ 65 age ≤ 74 years,75 ≤ age ≤ 84 years and age ≤ 34 years), the ethnic composition of people (normalized to a total size of 100) is used to infer dominant patterns.

#### 6.2.1 Ethnic divide of U.S Population across age groups

The underlying distribution of the U.S population into these two demographic coordinates is represented in Figure 21. One can draw a stunning contrast from these age-split statistics. In every age group, the dominant shareholder is the Non-Hispanic Whites, followed by people of the Hispanic race. However, the relative difference between these two ethnic groups reduces while traversing from higher to lower age groups. For example, in the 85 or more years old segment, the Whites hold a high of 76% and the Hispanic a modest 8% share of the population (In this particular group, the black population share is similar to the Hispanic numbers). As we move down in age, the percentage of Whites reduced, and the Hispanic community increased. For example, in the 0 − 24 age group, the White and Hispanic respectively balance the share percentage to 50 and 25%, of that given segment population. The ethnic black population also has an increased percentage share towards younger age groups.

**Figure 21.**
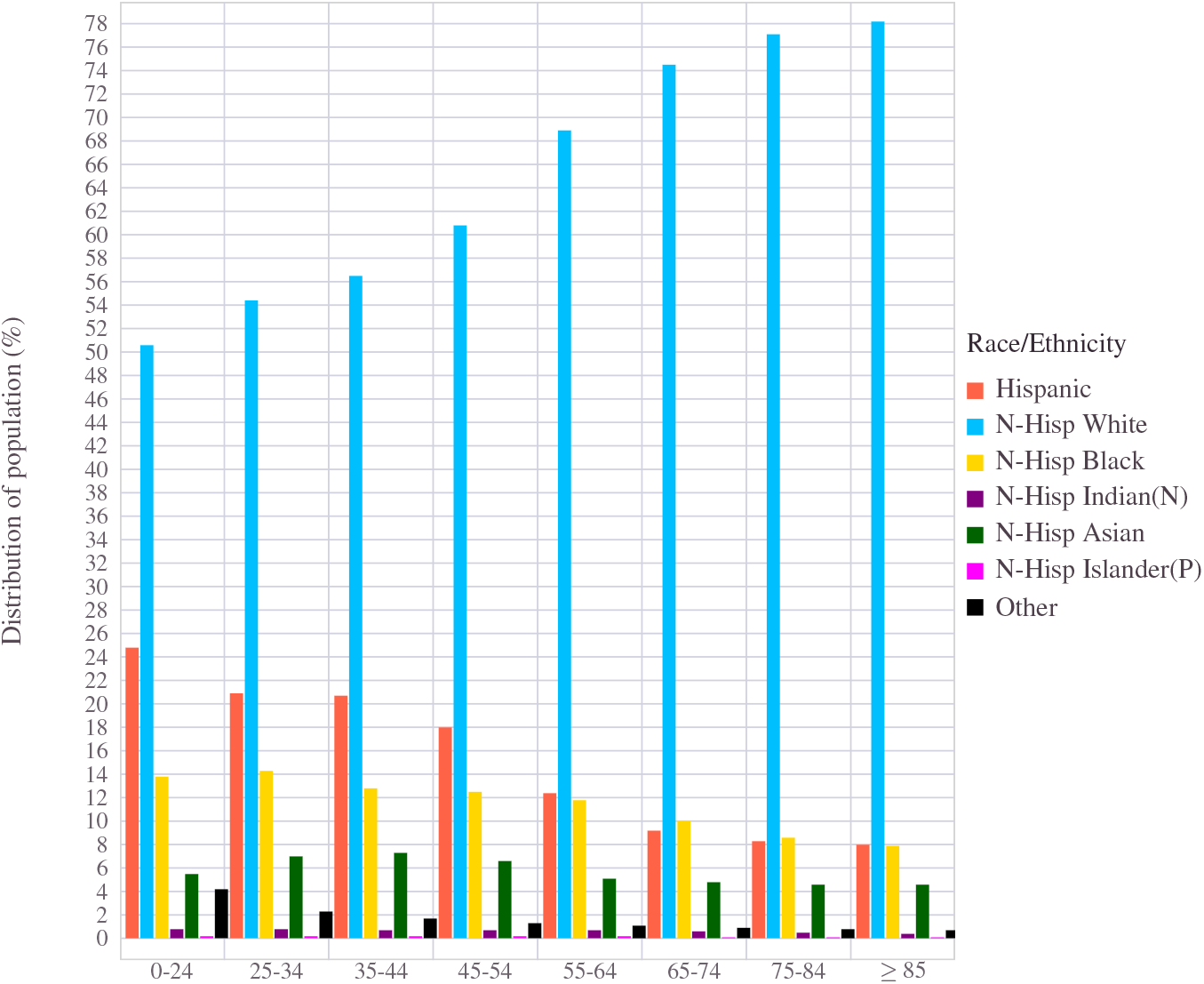
The U.S national population divided into specific age groups and within each age-cohort, the ethnic distribution is shown.

#### 6.2.2 Ethnic divide of COVID-19 deaths across age groups

Likewise, the distribution of mortality figures split into race and age is shown in Figure 22. While the general trend of the dominant race being White stayed intact in the higher age groups, a dramatic percentage increase of the death count can be observed in the Hispanic population at low to middle age groups. As a matter of fact, in every age group, the percentage share of the Hispanic race stood above their percentage share in the population. More Hispanic individuals of less than 45 years old died due to the pandemic than individuals from the White community.

**Figure 22.**
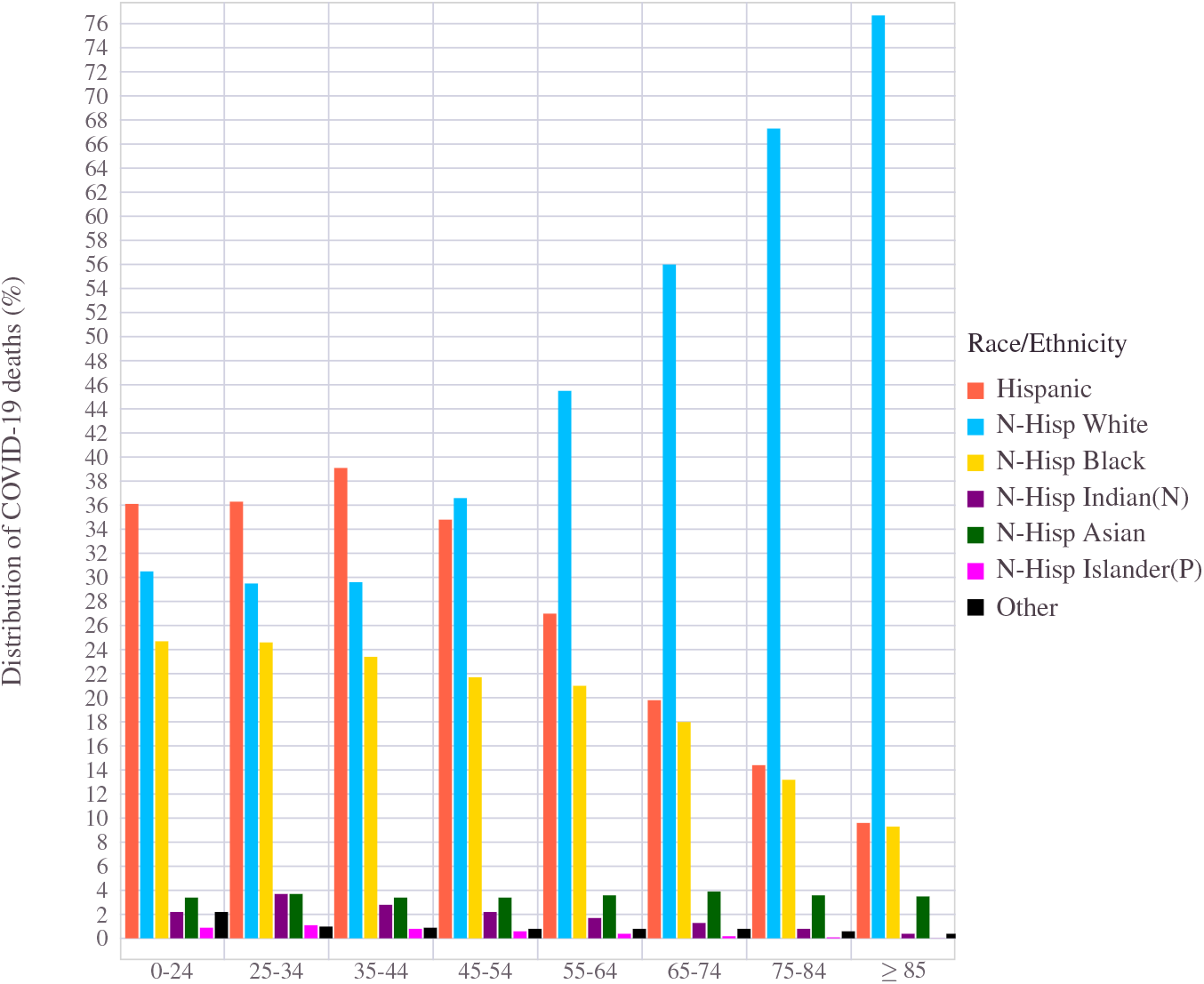
The total number of deaths due to COVID-19, divided into non-overlapping specific age groups and within each age-cohort, the ethnic distribution is shown.

Almost in identical fashion to that of the Hispanic race, the Black community appeared to have a disproportional increase in percentage share of deaths across all age groups, a drastic increase visible particularly towards the younger side. Among the deceased, belonging to the sub 55 years old group (i.e., age *<* 55), the White individuals accounted for less than that from the Hispanic and Black race combined. Even though a more significant number of White deaths took place in the older age category, it remained comparatively lower than their population strength. In all, a clear disadvantage is noticeable in the Hispanic and Black communities.

An interesting observation is on the Non-Hispanic Asian population. Across all age groups, this race has fared slightly better in terms of COVID-19 mortality in comparison to their population presence. The absolute number of Native Indian people is lower at the federal level. However, when it came to COVID death, they too had to face a disproportional increase in burden due to the pandemic. Towards the younger side of the population, this phenomenon gets exacerbated.

### 6.3 Joint analysis: Ethnicity, Age and Geography

As discussed in the Section 6.1.3, from the age-masked data (i.e., view the state-wise data bundled by ignoring the age information altogether), we could establish a first-degree correlation between race/ethnicity and the COVID-19 mortality rate. The scatter plot in Figure 20 has the mapping of the age-agnostic COVID-19 death counts against the corresponding population demographic data. The state-level and the country have this mean trend where many points align closer to the reference line. From the Figure 20 the second-degree inference that can be derived is the higher than expected death counts that people of the Black ethnic community had to suffer. Statistically speaking, this has more to do with the variance. Even though their percent share per state population remained small, the burden of death caused by COVID-19 has been remarkably higher for this race. On the other hand, the Hispanic community did better than its average population share in many places, except two of its stronghold states, California and Texas. The Non-Hispanic White population was also hit by slightly worse than expected mortality numbers in many states. The states with moderately lesser populations statistically had the mortality figures raked higher than the population average.

Skipping over the age information from the analysis and looking purely at a state level has a significant drawback. Firstly, as we have seen already, the COVID-19 mortality skewed heavily towards older individuals. While this is a visible situation in diverse groups, the stark impact of age is particularly present among White individuals. Therefore, to assess the inherent pattern in the data, we have to view the data at a more granular level by considering age, ethnicity, and geography.

Figure 23 illustrate the correlation between mortality and population distribution as a scatter plot representation from the raw age-ethnicity demographic data from all U.S states. The line (affine equation with slope = 45 degrees) is a boundary reflecting 1 : 1 mapping between mortality rate and population share, which in a way is the expected reference line. Staying above (left side) suggests a worse than expected mortality situation, whereas below signifies better mortality figures. Compared to Figure 20, the emergence of age as the dominant aspect of COVID mortality is clearly visible in Figure 23. For the country as a whole, the Non-Hispanic Whites ended up with significantly better than expected COVID-19 mortality figures across all age groups. This increased gap is more significant than the age-agnostic data presented earlier in Figure 20. Among the agewise results, the younger people stood out with incredibly better statistics than expected based on its population proportion. This de-masking of age’s role in the pandemic dynamics has also exposed the inequality levied upon certain ethnic groups.

**Figure 23.**
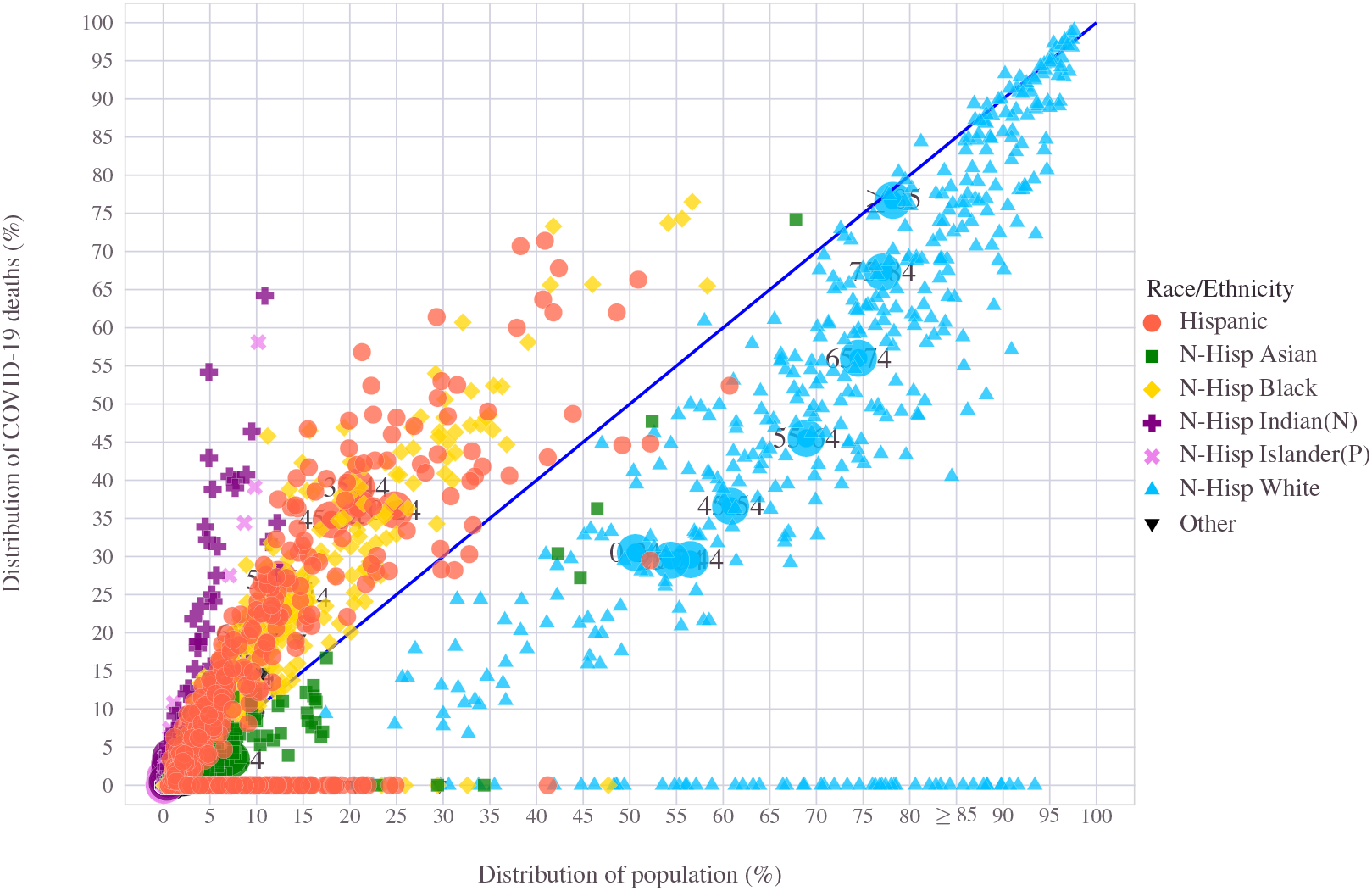
COVID-19 deaths across ethnicity in proportion to population

On the other hand, the Hispanic and Black (Non-Hispanic Black) found themselves on the other side of the reference line. In every age group, these ethnic groups ended up with a disproportionally higher number of deaths than their share. Other communities such as Asian and Native Indian and Pacific islanders have reduced presence compared to national numbers. While Asian individuals fared better, the ethnic people had a higher than average loss. In every ethnic cohort, the age-wise average mortality measure is extracted (from Figure 23) and explicitly shown in Figure 24. The figure shows that the Non-Hispanic White community, the dominant shareholder in every age group, has stayed much better than their proportional presence in the population. Within this group, a gradual improvement is seen towards the lower age group. For the Hispanic and Black communities, the overall situation pretty much reversed. The damage done by the pandemic is much more pronounced on the lower to the middle-aged segment of the community than the oldest lot, as seen in Figure 24. The situation for people from other ethnic background shown separately in Figure 25 clearly illustrate the severe damage the pandemic has done on other communities. Other than the Asian race, all other Hispanic, native, or Black ethnicity have suffered disproportionate mortality rates.

**Figure 24.**
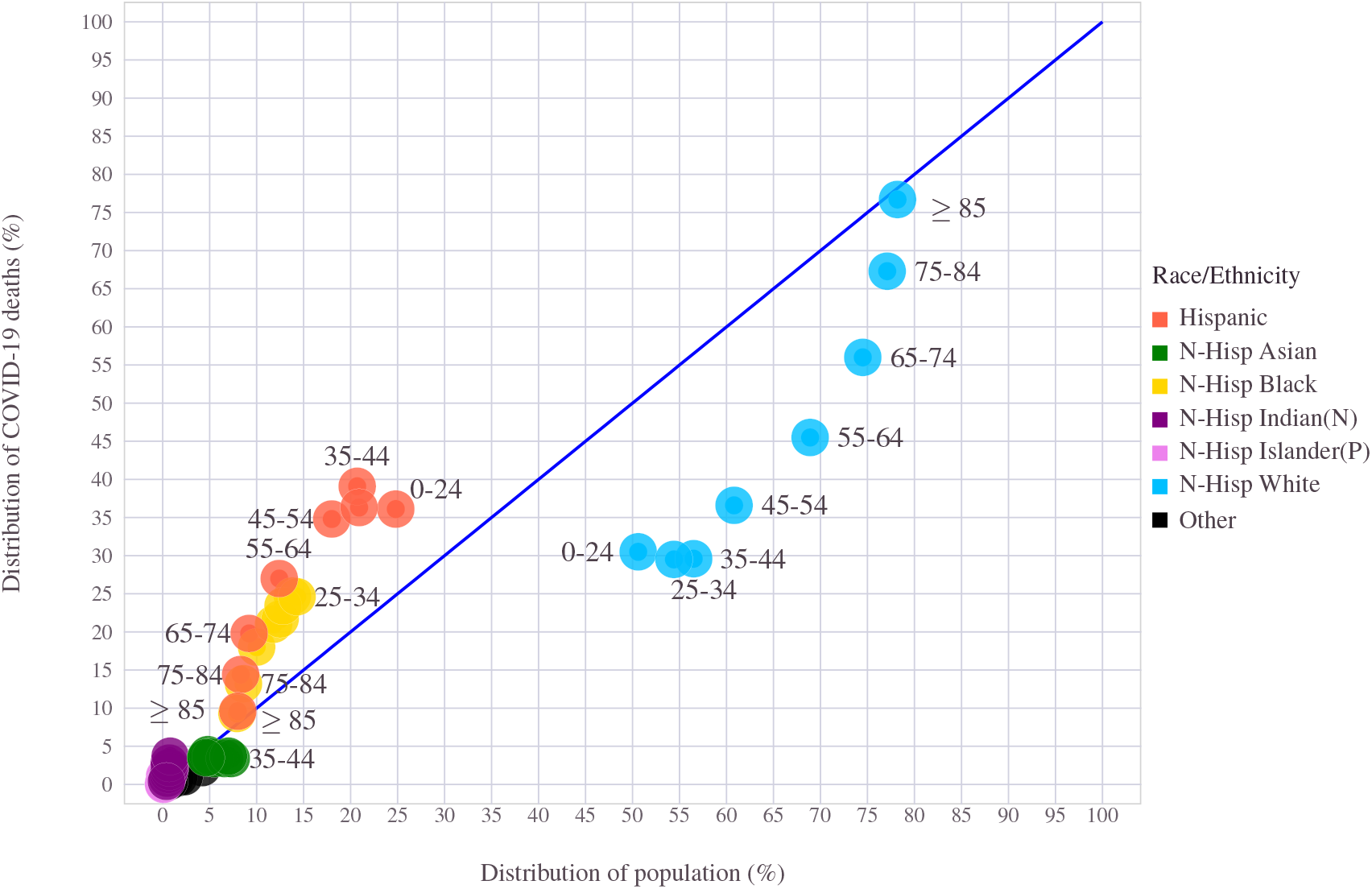
COVID-19 death across ethnicity in proportion to population. Averaged over the country data per each age group

**Figure 25.**
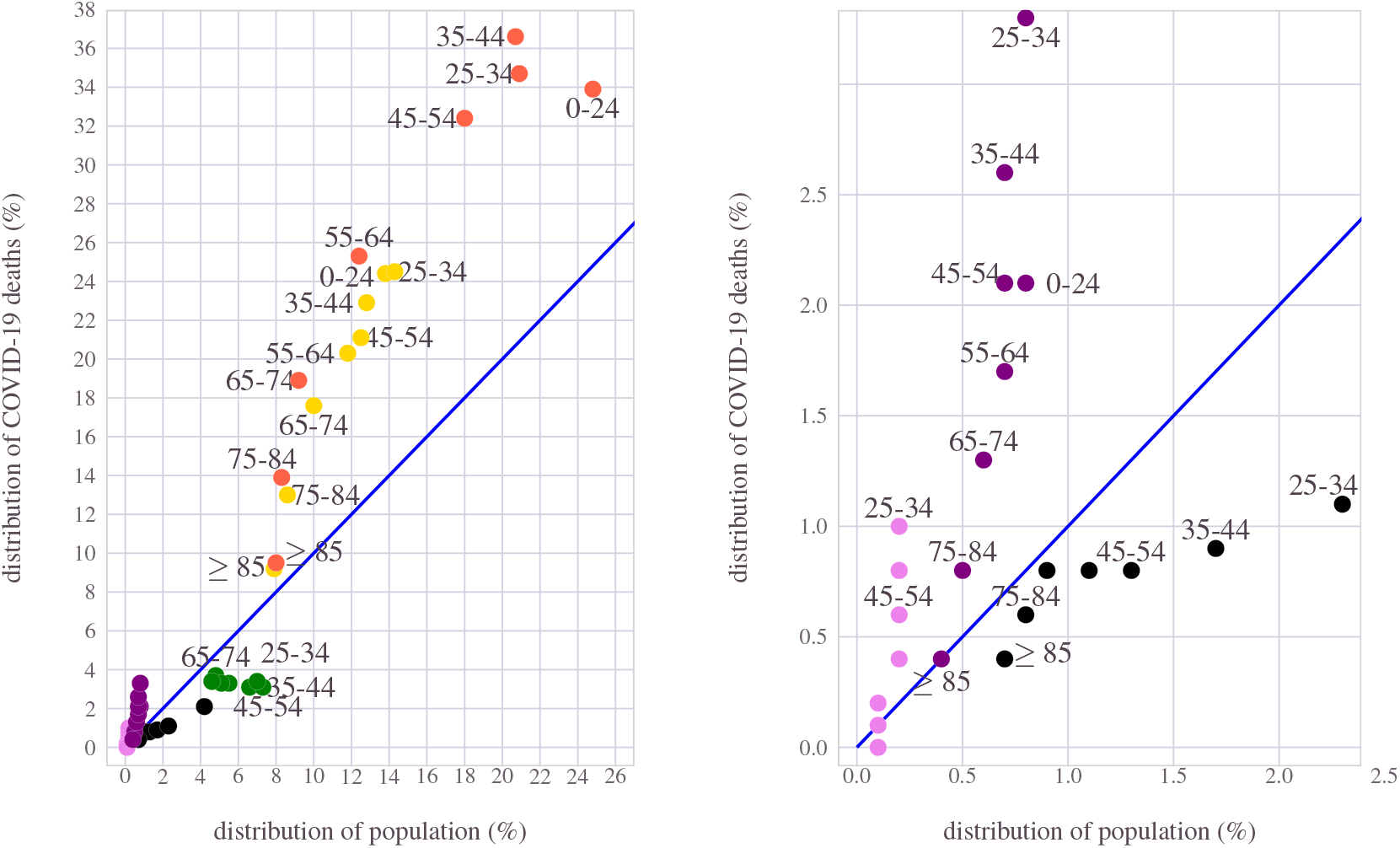
COVID-19 death versus population in U.S. Ethnic groups other than Non-Hispanic White groups are highlighted.

Another illustration of this disproportional demographic variation is shown in Figure 26, where Δ, which measures the difference between the age-wise COVID mortality rate and the corresponding population, is illustrated. The underlying information used here is age-ethnicity divided data, collected from individual states. A negative value of Δ implies lower than average deaths w.r.t the population strength, and a positive value reflects a condition of relatively higher mortality. The Non-Hispanic White and Asian communities, for the most part, have found themselves doing somewhat better than other communities.

**Figure 26.**
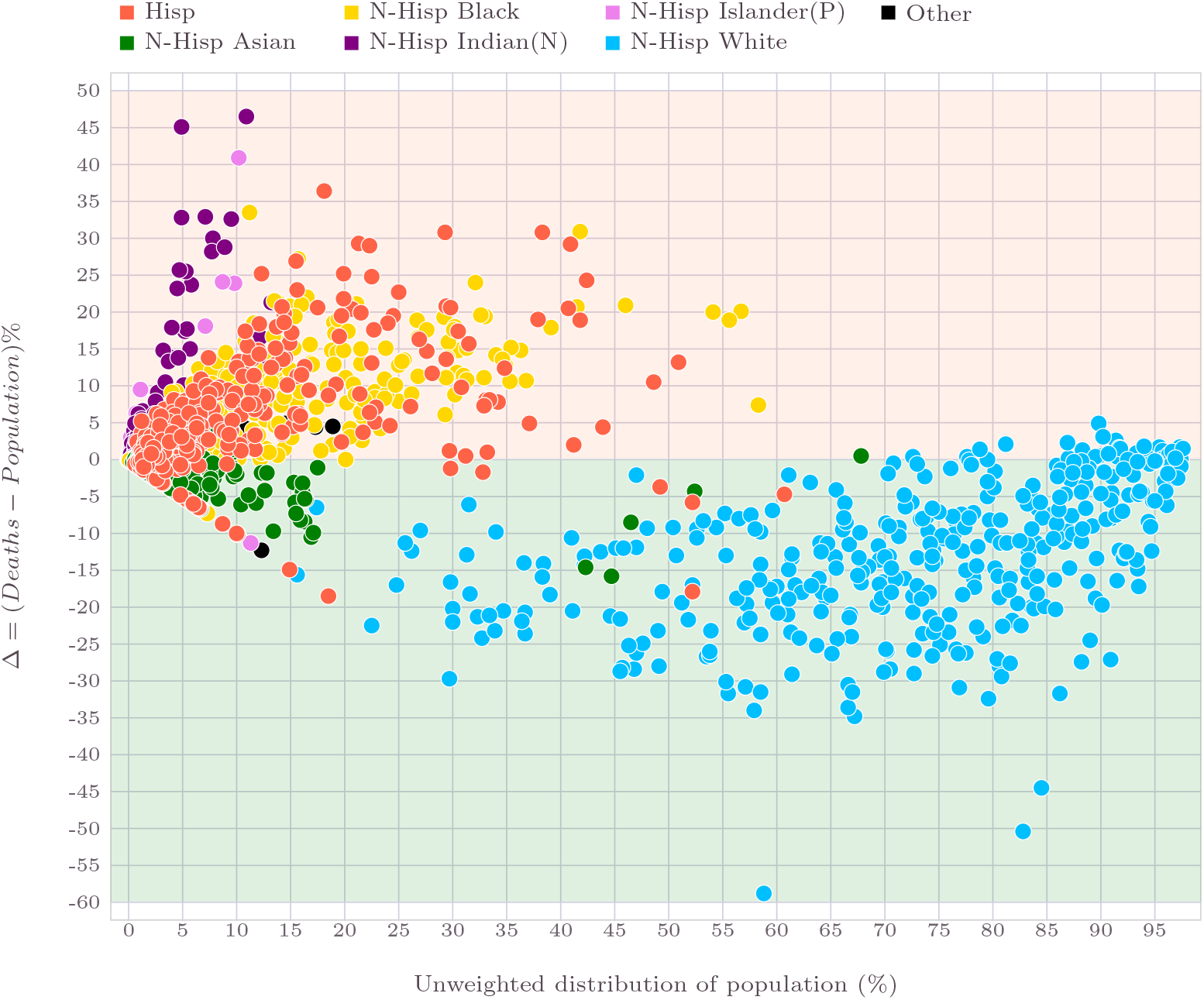
COVID-19 death among ethnic groups. The difference Δ between the percentage share of COVID-19 deaths and the corresponding population proportion is shown. Δ *<* 0 is an indication for better than expected mortality rates based on population. A positive value of Δ implies a dis-proportionally higher number of deaths than mandated by the population situation.

Given that various ethnic groups occupy vastly different shares when it comes to the population, the normalized difference between the mortality rate and population mix may serve as a better illustration of this phenomenon. For this purpose, we define the metric *ν*(*s*) as follows:

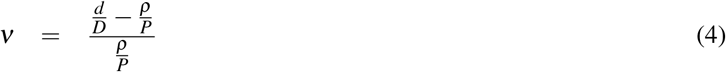

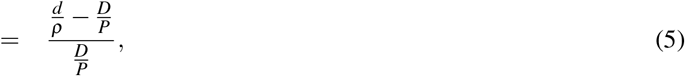

Where *d*≡*d*(*s*) is the number of deaths matching to a specific demographic subset *s* whose population is *p*≡*p*(*s*). The aggregate number (union over the entire demography 𝓁 = ⋃ *s*) of deaths is denoted by *D* (note that *D ≡ D*(*s*) ⋃ *d*(*s*)) and with a total population *P* (also *P*≡*P*(*s*) = *ρ*(*s*)). Recall that the numerator in Eq.4 is the difference metric Δ discussed earlier. That is, 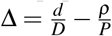, which is also depicted in Figure 26.

In this section, we considered the demographic subset *s* = (age, ethnicity). This metric *ν ×*100 is plotted in Figure 26. A positive value suggests a dis-proportionally higher (worse) mortality than expected based on its population. A negative value is an indication of having had better death statistics than predicted by the population scenario. The disproportional damage done by COVID-19 to ethnic Hispanic and Black people is also clear in Figure 26 with higher *ν* values. People of the White race braced with better statistics overall, as seen with the negative *ν* values.

The *ν* values computed age-wise for each of the ethnic group with the data illustrated in Figure 24, is shown in Figure 27. The shocking impact of COVID-19 driven mortality that has fallen on the Native Indians and the Pacific Islanders, especially the lower and middle-aged people, only emerges when viewed from this normalized approach. For the oldest age group, pretty much all ethnic communities faced a population accorded mortality Figure The White and Asian communities have ended up with better *ν* numbers for all age groups less than 75 years old. The Hispanic and Black communities have a fairly similar *ν* behavior across most age groups. Individuals of age less than 74 from these communities have witnessed nearly 75% more deaths than dictated by their population share. The Native Indians and Pacific Islanders have drastically higher mortality rates in the low and middle age groups. For example, for the younger people between 25 and 34 years old, the pandemic resulted in almost 4 times more deaths than the cohort’s population strength.

**Figure 27.**
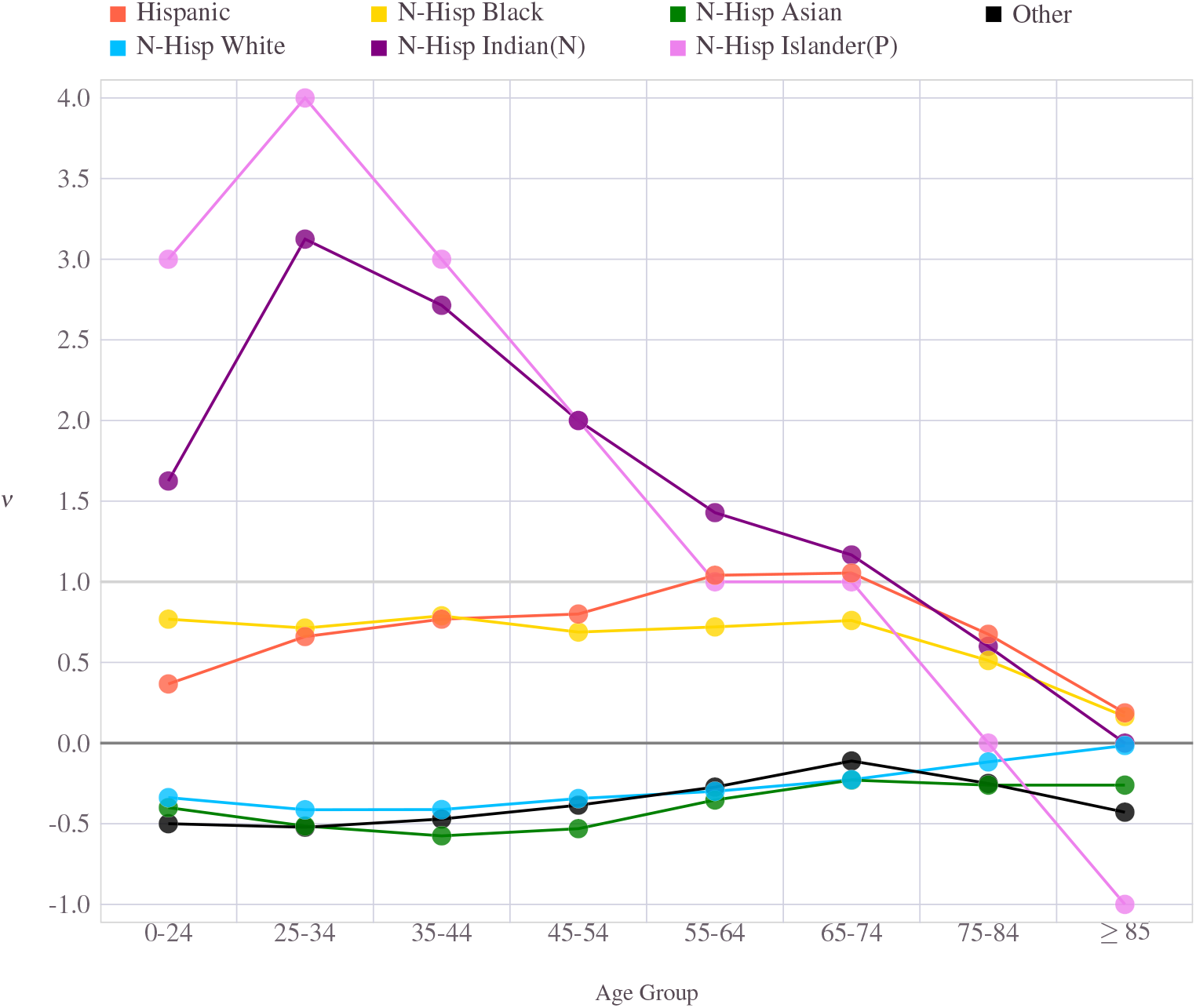
Ethnic groups and their *ν* values

## 7 Comparison with other respiratory diseases

Fundamentally, COVID-19 is a respiratory illness. Majority of the people infected with the COVID-19 virus exhibit mild to moderate symptoms such as elevated body temperature, fever, coughing, and shortness of breath. Some infected subjects get pneumonia and advance to severe illness, which can spiral into fatal situations. In the early stage of the virus breakout, some parallels were drawn between COVID-19 and pneumonia. The World Health Organization (WHO) first referred to the virus as *novel coronavirus-infected pneumonia* (NCIP) before adopting the name COVID-19. The SARS-CoV-2 virus in Wuhan has initially identified as *cases of pneumonia of unknown etiology*, as per the WHO report in January 2020. Influenza is another common respiratory illness. In this section, we will attempt to compare the mortality statistics of COVID-19 with these two diseases.

### 7.1 Pneumonia and COVID-19

Pneumonia refers to a broad range of lung infections caused by viruses, bacteria, and, in some cases, triggered by certain types of fungus. A bacterial or viral infection can trigger inflammation, leading to significant damage to one or both lungs. The resulting fluid and infection debris build-up often cause serious breathing trouble to patients, necessitating ventilator support and or oxygen therapy. Irrespective of the underlying agent that triggers the inflammation, pneumonia can turn into a dire, life-threatening health situation.

Pneumonia caused as a result of the COVID-19 virus is sometimes also referred to as COVID-Pneumonia. Early evidence suggests that COVID-pneumonia often causes damage to both the lungs. Some people with COVID-pneumonia get acute respiratory distress syndrome (ARDS), a severe lung failure condition.

Not all COVID-19 infections lead to COVID-pneumonia. People with obesity, diabetes, older age, cardiovascular or underlying lung illness are considered high-risk patients for COVID-Pneumonia.

Stacking up the total deaths due to pneumonia (including COVID-pneumonia and those who died due to pneumonia without any COVID-19 connections) and the total deaths reported due to COVID-19, it emerges that the latter outnumbered the former. A gender-wise comparison, for each age group is presented in Figure 28. While the COVID-19 deaths are higher for every age group and both males and females, the females showed larger deviation among the older age category, particularly those with 85 years and higher.

**Figure 28.**
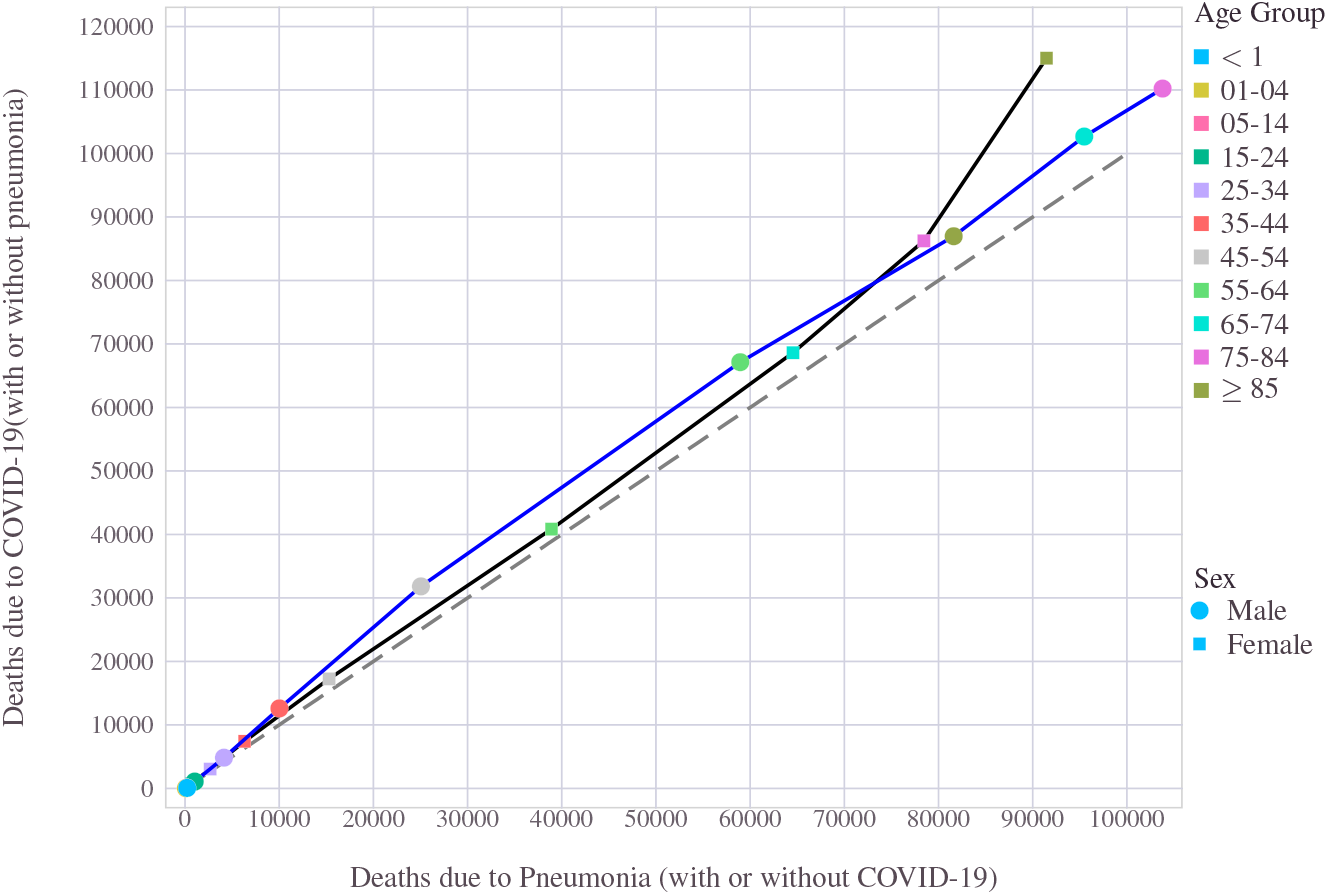
Pneumonia vs. COVID-19: Comparison of the total number of the absolute number of deaths attributed to pneumonia or COVID-19. Those patients infected with both COVID-19 and pneumonia are included in pneumonia and COVID-19 categories.

Isolating the COVID-pneumonia cases from the mix and thereby comparing the non-covid caused pneumonia deaths with that of total number deaths due to Covid-19 alone(but without any symptoms of pneumonia), we can observe that the gap has widened further. In other words, the number of deaths due to COVID-19 (even discounting COVID-pneumonia subsets from it) is significantly higher than that caused by Non-COVID pneumonia. The severity of COVID-19 also gets exacerbated towards older age and most noticeably for females with age 85 years or older. This behavior is illustrated in Figure 29.

**Figure 29.**
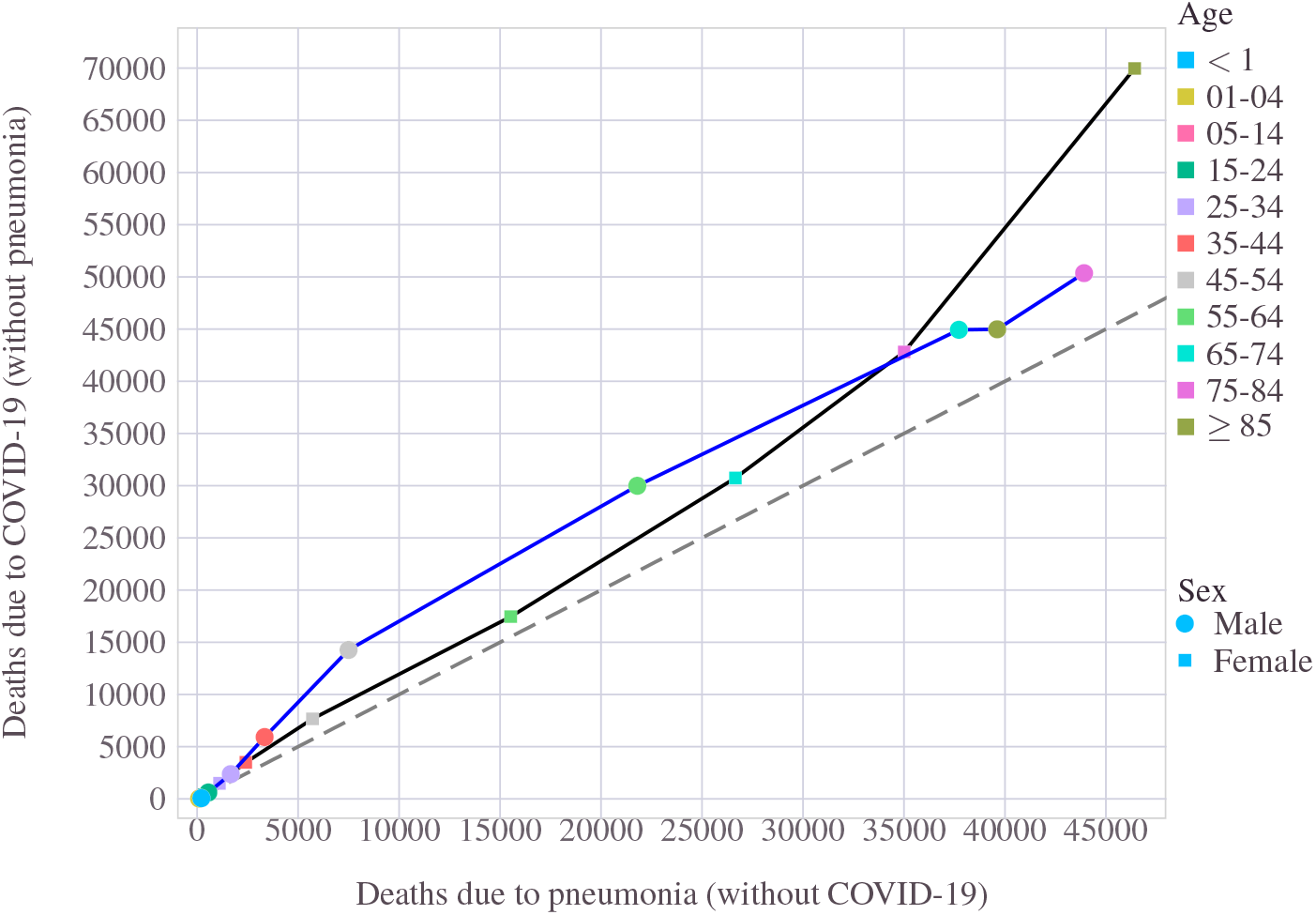
Pneumonia vs. COVID-19: The number of of people who died due to COVID-19 without any pneumonia symptoms is compared with the total number of non-COVID pneumonia deaths.

Given that a good number of COVID-19 patients advanced to pneumonia, it is worthwhile to quantify the percentage of individuals(among the deceased) who found to have advanced to COVID-pneumonia. This ratio is presented in Figure 30. Among the younger people who fell due to COVID-19, relatively less number had advanced to COVID-pneumonia. From the deceased subjects who were under 25 years old, more than 70% of the deaths were not due to pneumonia complications. However, the adult population (i.e., age range 25 − 74 years) witnessed nearly 50% of its death count attributed to pneumonia. When it came to the oldest age groups, one could observe a slight drop in COVID-pneumonia share. An interesting gender bias that emerged is how females in the oldest age category perished without exhibiting any pneumonia symptoms. For the 85+ years old groups of individuals, when 48% of the COVID-19 deaths among males reportedly also had pneumonia, the corresponding fraction stayed below 39% among the females.

**Figure 30.**
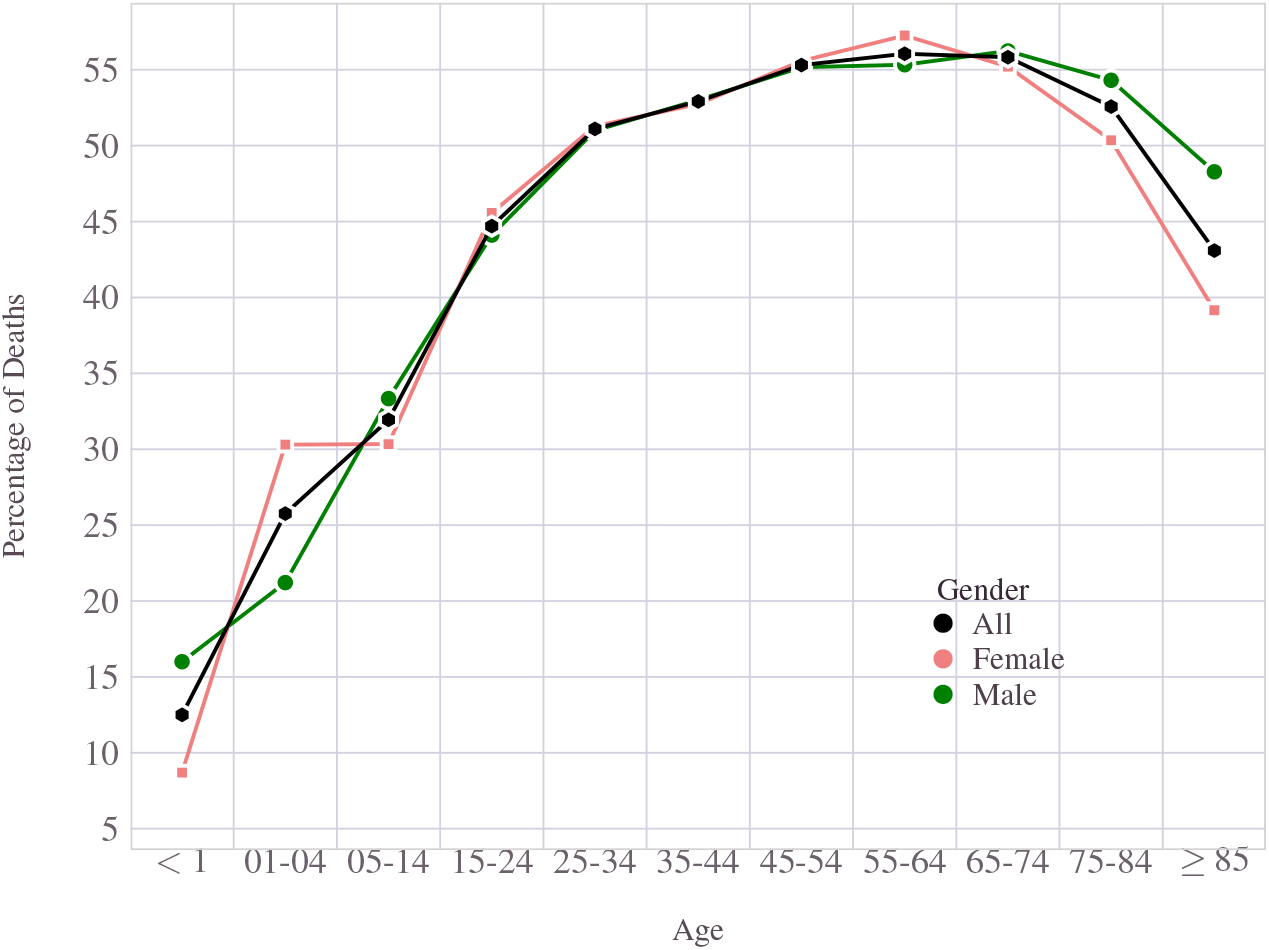
Age-wise distribution of people died due to COVID-19 who also had pneumonia.

The mortality distribution across age groups and the role gender played in the statistics is worth mentioning. The age-wise behavior of COVID-19 deaths was discussed in Section 4. Here, we place pneumonia and COVID-19 side by side and contrast their statistics. A striking similarity between pneumonia mortality and COVID-19 induced deaths soon became obvious. Figure 31 depicts the overall deaths attributed to the two infections across different age groups and gender. The death counts in each category include the joint headcounts (i.e., COVID-pneumonia). That is, the COVID-19 category also includes those individuals who had advanced from COVID-19 with pneumonia symptoms and vice versa. To isolate this dependency, we also analyze the disjoint category sets and show the comparison in Figure 32. Other than the slight variation triggered by the higher fraction of female deaths in the ≥ 85 years old category, the statistical similarity in mortality across age is well established. Like COVID-19, the most significant share of deaths among males took place among 75 − 84 years. This group attributed to 27− 28% of fatalities due to COVID-19 as well as pneumonia. Thus, a slight drop in percentage among the males among the oldest adult group. Statistically, females, on the other hand, exhibited a monotonic increase in percentage share as they progressed age-wise. Females who were ≥ 85 years old attributed to about 35% of the total (female) non-COVID pneumonia deaths. The corresponding figure for non-pneumonia triggered COVID-19 deaths stood higher at 42%. For the COVID-19 deaths caused by pneumonia, the numbers are 36% and 32%, respectively.

**Figure 31.**
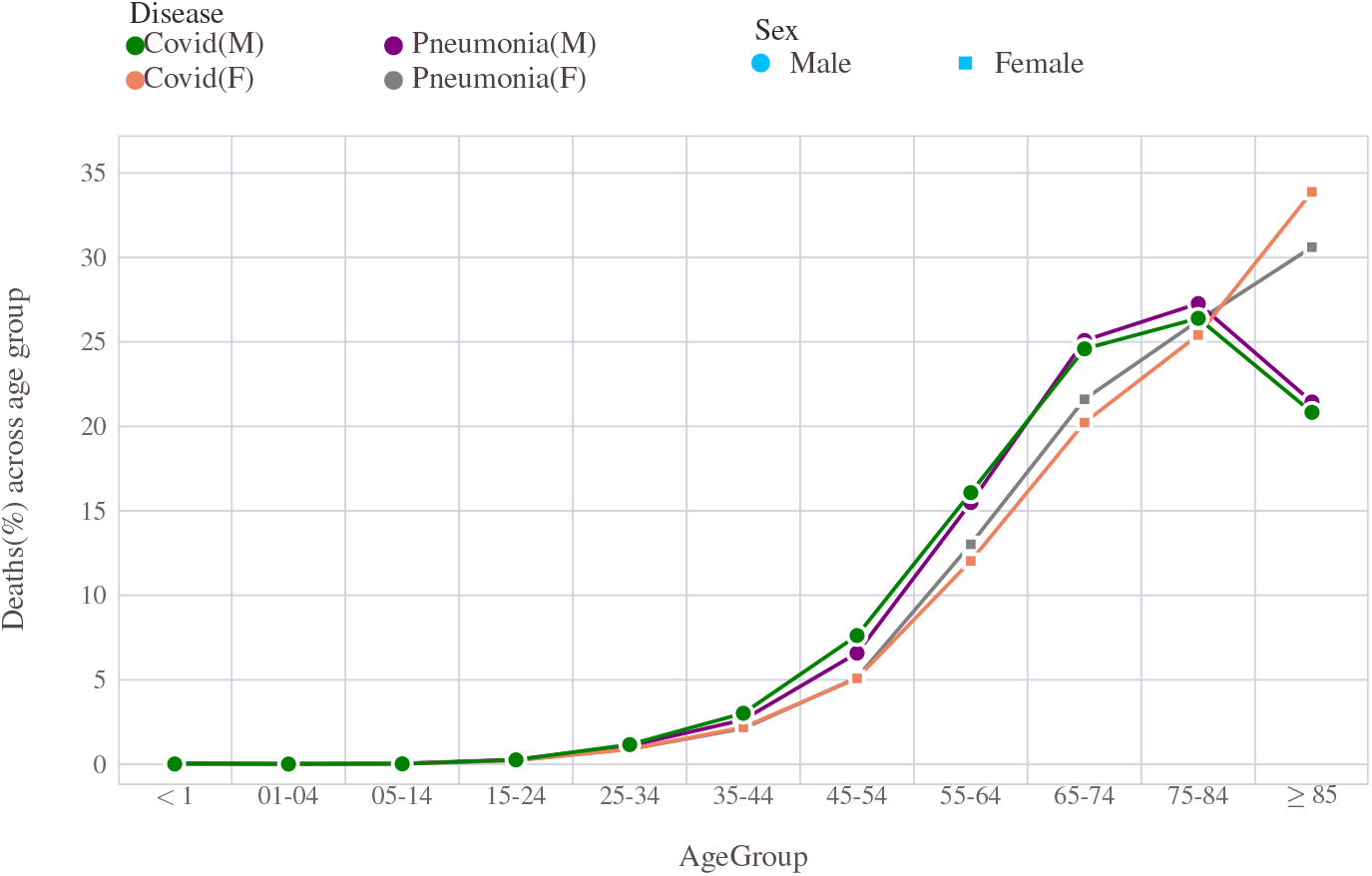
Pneumonia or COVID-19: Age-wise distribution of people died due to COVID-19 or pneumonia. A close similarity in age profile exists between the mortality figures of the two diseases. In this figure, people infected with both COVID-19 and pneumonia are included in both the sets.

**Figure 32.**
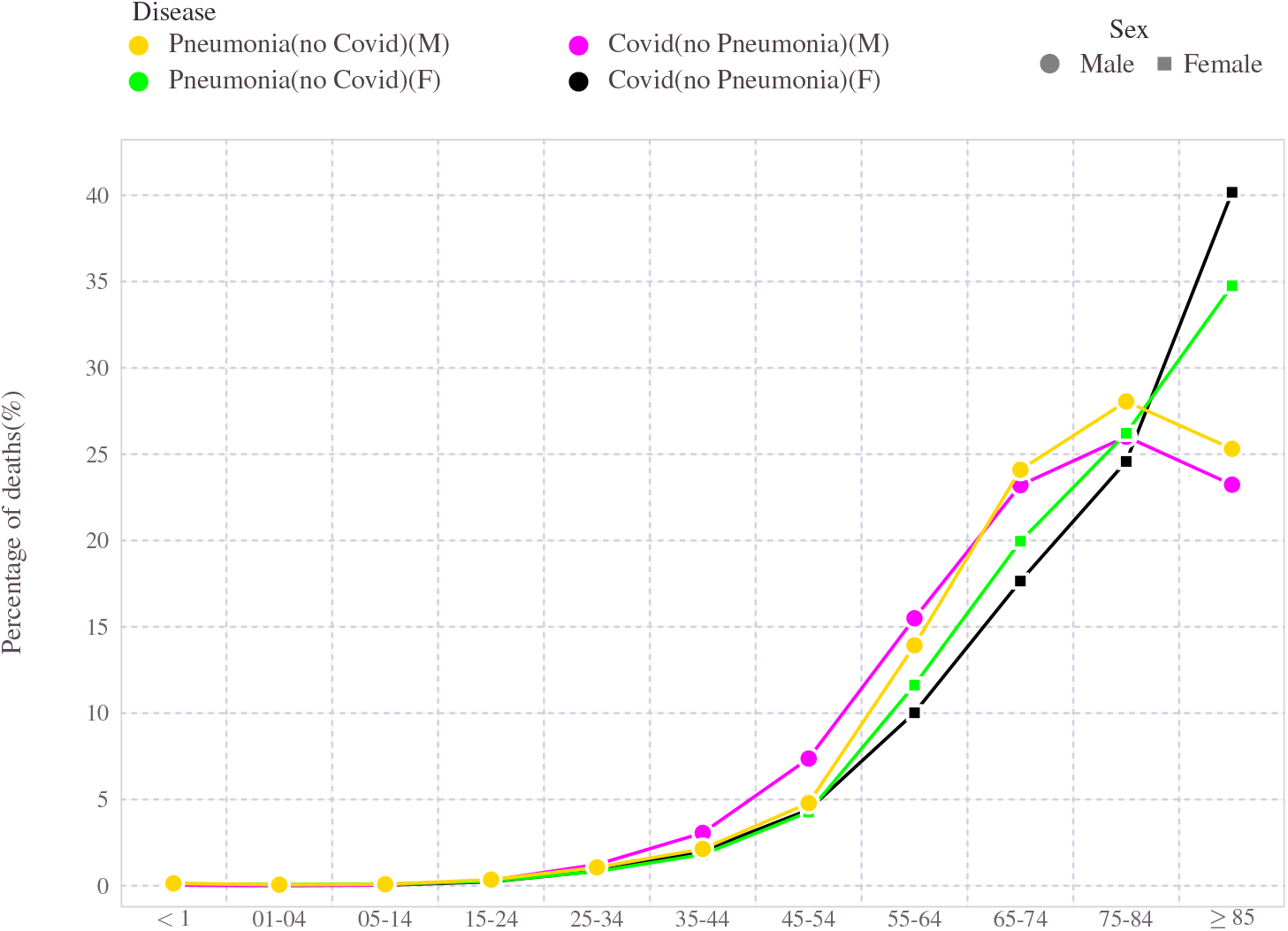
Pneumonia or COVID-19: Age-wise distribution of people died due to COVID-19 or pneumonia. A close similarity in age profile exists between the mortality figures of the two diseases. For a fair comparison, the COVID-19 deaths here do not include diseased people who had pneumonia. Similarly, the pneumonia death discards any individuals who also had a confirmed COVID-19 infection.

### 7.2 Influenza and COVID-19

In the early days when the COVID-19 pandemic hit the world, there were reports drawing parallels to commonly known flu, aka Influenza Influenza^36,37,52^. Influenza and COVID-19, have many things in common. They both are contagious respiratory illnesses. However, they are caused by different viruses. The COVID-19 virus variants are generally more virulent than flu viruses, resulting in an accelerated spread of diseases across populations over a broader geographical span.

Compared to previous years, in the United States, the Influenza cases and deaths were drastically more minor in 2020 and 2021. Between January 2020 and October 1, 2021, throughout the USA, the total number of fatalities attributed to Influenza is only 9343 (8934 of them were from 2020). Statistically, the mortality due to Influenza distributes itself to slightly lower age groups than COVID-19. The highest share of COVID-19 deaths belonged to 75 − 84 years old individuals, whereas the highest-ranked males were ten years younger (65 − 74) on average. Influenza also appears to be less severe than Covid-19 most senior age groups. To a certain degree, the deaths caused by Influenza are spread broader and less skewed and spread across a wider age group than COVID-19. However, a common theme in these two respiratory illnesses is the relative immunity enjoyed by the younger population. For those under 45 years old, their combined share accounted for less than 10% of the total death counts from these illnesses. Figure 33 has the comparison drawn. Like COVID-19 and pneumonia, the females managed to push the vulnerable age higher by 10 years approximately.

**Figure 33.**
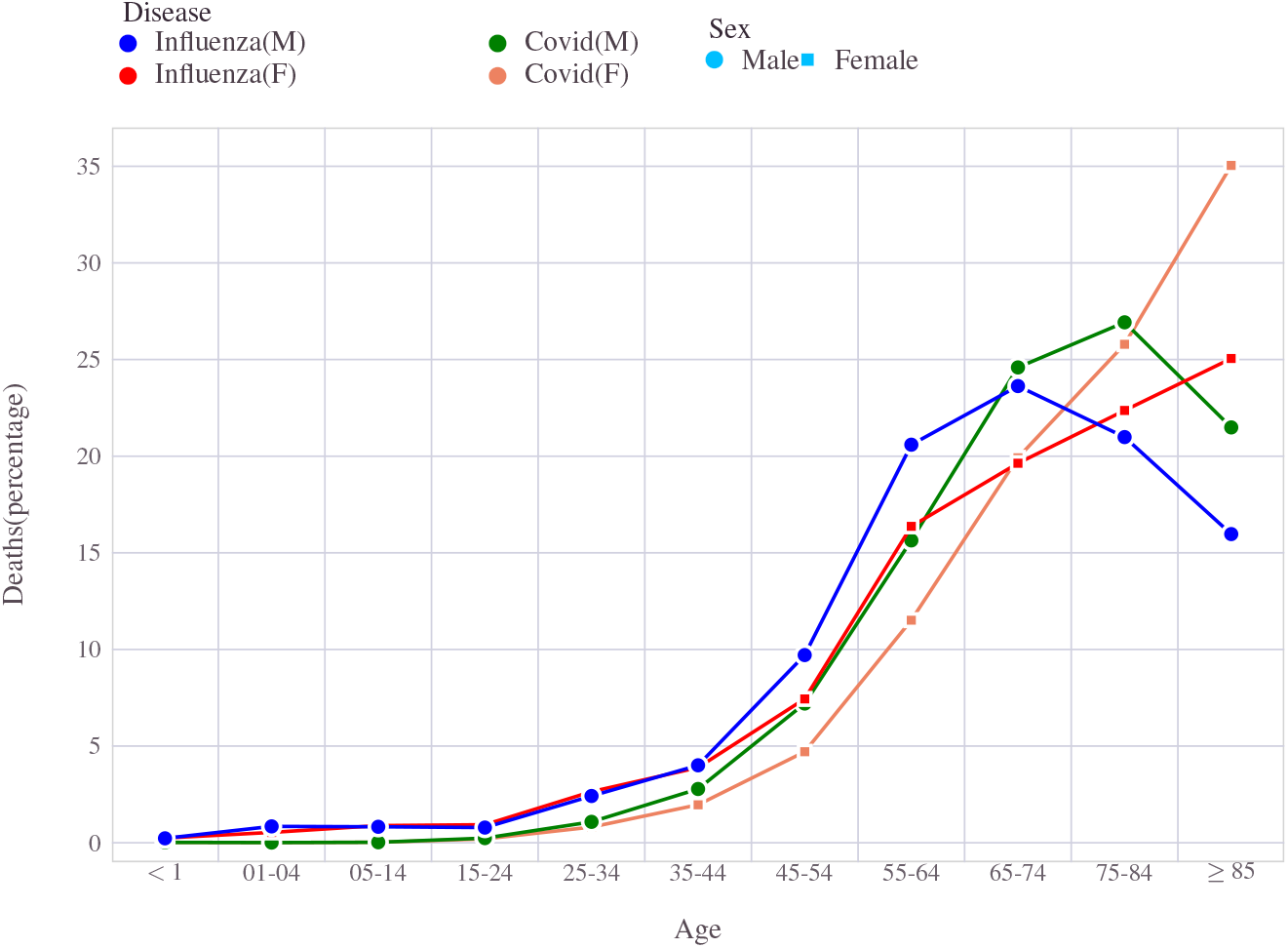
Influenza and Covid: Age profile of the individuals who died of Influenza compared with COVID-19 statistics.

## 8 Virus genealogy and impact due to variants

Coronaviruses come from a family of lineages^64^. A group of related viruses with a common ancestor is termed a lineage. For example, the virus SARS-CoV-2 has many lineages, all of which cause COVID-19. Like all viruses, coronaviruses adapt themselves to their needs by constantly changing through mutation. A mutation refers to the phenomenon where a change in a virus’s genetic code (gnome) takes place. These mutations occur very frequently, and such events sometimes lead to changes in the virus’s characteristics. This phenomenon leads to virus variants. A variant is a formal name for the underlying viral genome (genetic code) containing mutations. Thus, new variants emerge over time. A few variants come to the scene and disappear quickly, without making their presence felt in a big way. A few of them, however, emerge stronger and survive long enough. Some of the persisting viruses that cause COVID-19 have been identified globally, classified, and documented.

Coronaviruses are named for the crown-like spikes on their surfaces. Scientists monitor changes in the virus, including modifications to the spikes on the surface of the virus. These studies, including genetic analyses of the virus, are helping scientists understand how changes to the virus might affect how it spreads and what happens to people infected with it. Some of the dominant variants of the SARS-CoV-2 circulating the world which made significant inroads within the United States are B.1.1.7 (Alpha), B.1.351 (Beta), P.1 (Gamma), and B.1.617.2 (Delta), among which the Delta (B.1.617.2) variant is classified as variants of concern. The B.1.526 (Iota) is another variant that has been identified to cause an increased mortality rate among older adults^53^. The WHO maintains a website listing a known list of variants, their new names, and details^56^.

These variants seem to spread more easily and quickly compared to other variants. For instance, the newer variants like Alpha and Delta are very contagious and infect far more people than the virus that triggered the pandemic. According to recent research, a plausible explanation for this is that these new variants can spread more efficiently through the air. The major coronavirus variants confirmed to have a presence in the U.S are shown in Figure 34 along with their timeline during the pandemic.

**Figure 34.**
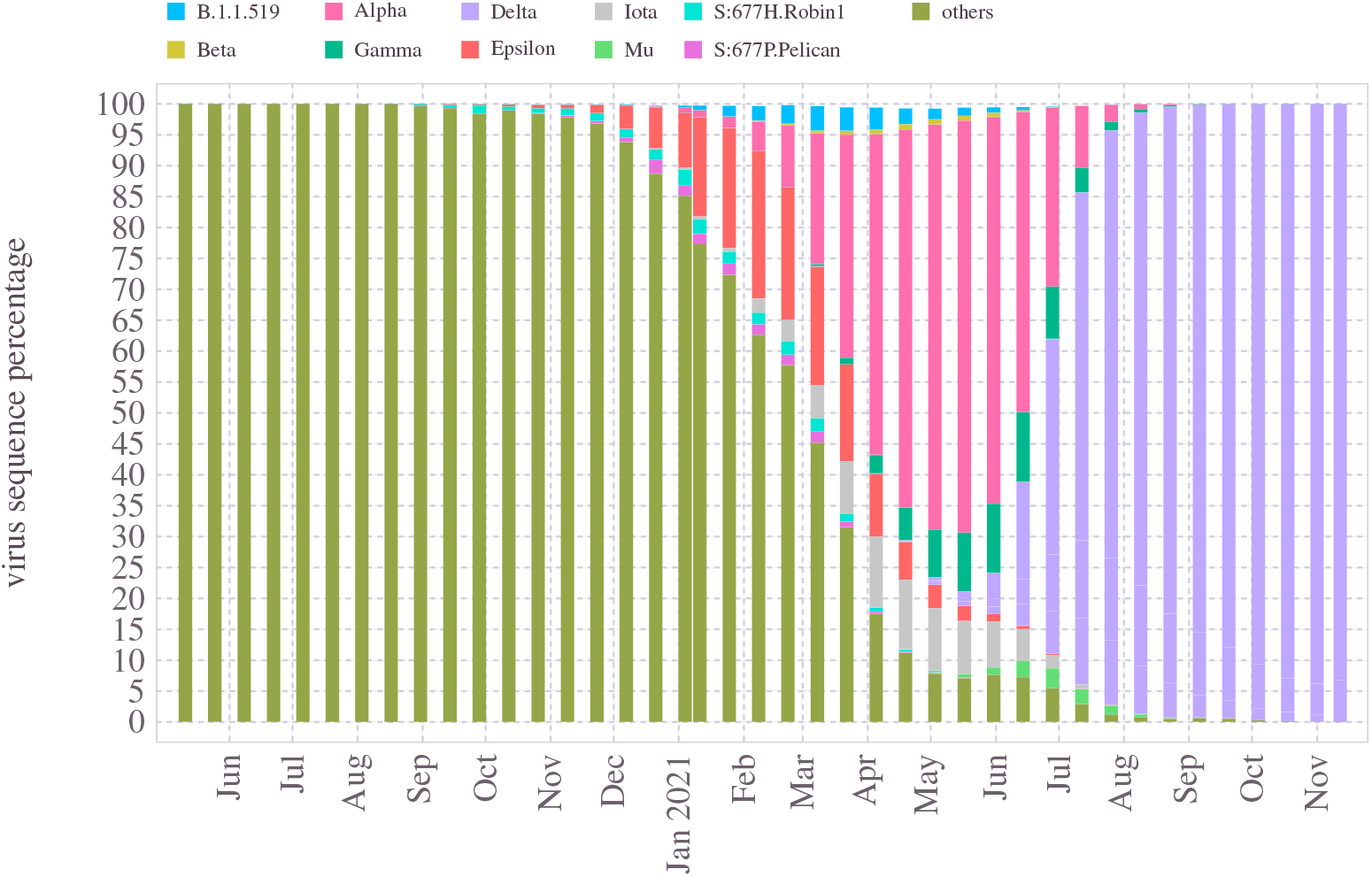
The dominant coronavirus variants confirmed in the United States and their timeline.

## 9 Vaccines

Immediately after the virus began its surge in the early months of 2020, a quest for an effective treatment also began. As a result, experts have almost immediately concluded the urgency of developing effective vaccines to deal with the virus and thus eventually curb and prevent further damages^42^.

Developing a vaccine entails a long and involved process, which demands enormous resources, research, coordination between diverse scientific expertise, and time^47^. Going by the history of earlier vaccines, developing a new indication would take anywhere from 4-10 years. As a reference, the previously fastest vaccine developed from viral sampling was for mumps in the 1960s, which took four years^62^. Nevertheless, in the case of COVID-19, scientists were able to come out with effective vaccines within less than a year, thereby enabling a clear path to deal with the pandemic finally.

Research laboratories, universities, and pharmaceutical companies worldwide have been actively developing medicines, including vaccines for COVID-19^49^. On 11 December 2020, in the United States, Food and Drug Administration(FDA), the central agency authorized to decide on matters such as drugs and health products, issued the emergency use authorization (EUA) of the Pfizer-BioNTech COVID-19 vaccine for persons aged 16 years and older. A week later, the FDA issued the second EUA for the Moderna COVID-19 vaccine. In addition, the Moderna vaccine EUA requirement placed the lower age requirement as 18 years. These emergency approvals paved the way for the vaccines to be available to the public in the United States. In the early months, vaccines were administered to people identified to be on the critical path. By April 2020, widespread availability of the vaccines was in place throughout the country for everyone who turned 16 and older. Even though the full FDA approval of the Pfizer-BioNTech vaccine came only later (August 2021), the quick deployment of vaccines to the public helped to arrest the pandemic to a considerably lower level from the peak number of cases and deaths in January 2021. With the recent FDA approval, the lowest age for vaccines is lowered to children 5 years of age or older.

Both Pfizer-BioNTech and Moderna vaccines belonged to a class of vaccines using mRNA^19,40,48^. The fascinating history of mRNA and its eventual emergence as a pandemic reliever is detailed in the Nature article^61^. In May 2021, FDA extended the emergency use of Pfizer-BioNTech vaccine to people who are minimum 12 years of age. Eventually, on August 23, FDA approved this vaccine, formally titled Comirnaty (COVID-19 Vaccine, mRNA), replacing the previously known name Pfizer-BioNTech COVID-19 Vaccine, for COVID-19 disease prevention in individuals 16 years of age and older. The FDA maintains this vaccine’s emergency use authorization (EUA) status for individuals 12 through 15 years of age^43,44^. In April 2021, a third vaccine, made by Johnson & Johnson (Jansen pharmaceuticals), also received EUA designation from FDA.

### 9.1 Type of COVID-19 Vaccines in the United States

The primary goal of a COVID-19 vaccine is to provide a means to acquire immunity against SARS-CoV-2. Depending on the underlying mechanism of action by which this goal is achieved, vaccines are classified differently.

All vaccines try to stimulate an immune response to an antigen, a molecule found on the virus. For the COVID-19, the characteristic spike protein found on the virus’s surface was identified to be the main driving force by which it invades the human cells. Therefore, this spike protein is used as the antigen for most of the COVID-19 vaccines. While some vaccines rely on smuggling the antigen into the human cell, others use clever methods by instructing the cellular engine within the human body to produce the viral antigen.

Among other things, prior research findings on the structure and mechanism of coronaviruses causing diseases such as severe acute respiratory syndrome (SARS) and the Middle East respiratory syndrome (MERS) have immensely helped in the accelerated development of COVID-19 vaccines. The SARS-CoV-2 genetic sequence data was made available by 10 January 2020. A detailed discussion on the science behind all COVID-19 vaccines is beyond the scope of this work. Therefore, we limit our attention to a high-level difference between the three vaccines administrated in the United States.

The Pfizer–BioNTech and Moderna vaccines belong to what is known as nucleic acid vaccines. Nucleic acid vaccines use genetic material – either RNA or DNA – to provide cells with the instructions to make the antigen. The Pfizer–BioNTech and Moderna vaccines rely upon RNA to stimulate an immune response. When induced into a human body, the RNA contained in the vaccine serves as messenger RNA (mRNA), triggering and instructing the cells to build the SARS-CoV-2 spike protein.

This clever mechanism of teaching the body how to identify and destroy the corresponding pathogen was adapted from the long line of breakthrough research on mRNA on other indications such as cancer. The advantages of nucleic acid-based vaccines such as these are that they are cheap and easier to make. In addition, the immune response would be strong because the antigen being produced resides well inside the human cells, usually in abundant levels.

The other significant type of vaccine is called viral vector vaccines, which use a carrier virus vector to deliver instructions to human cells. A virus vector is a modified version of a different virus. For example, the Johnson & Johnson vaccine adds the spike protein gene to a common virus known as adenovirus (a virus that causes colds or flu-like symptoms). This virus is modified to enter cells but without any ability to replicate or cause illness. Once the vaccine enters the human body, adenovirus delivers instructions that teach cells to make the spike protein. In turn, this will trigger the immune system to react by preparing antibodies. When exposed to COVID-19, these cells’ antibodies will attack the SARS-CoV-2 spike protein and build an effective defense mechanism against the virus. An overview of the various vaccine development and targeted therapies for COVID-19 is discussed in^18,41^.

### 9.2 Effectiveness of COVID-19 vaccines

There are many subtle differences between these vaccines in terms of storage and administration requirements. However, in analyzing the vaccine’s impact in controlling mortality and the overall pandemic situation, the primary information of interest is the efficacy data. Although each vaccine has unique attributes, all of them offer excellent protection against severe complications and deaths from the COVID-19.

Vaccine efficacy is a term used in epidemiology, which measures the effectiveness of the vaccine^17^. It is expressed as the percentage reduction of disease cases in a group of vaccinated people compared with a group in which no one is vaccinated. For example, vaccine efficacy of 90% suggests the situation where 90% decreases in the number of disease cases in the vaccinated group compared to a group not vaccinated at all.

Mathematically the vaccine efficacy(VC) is defined by the formula,

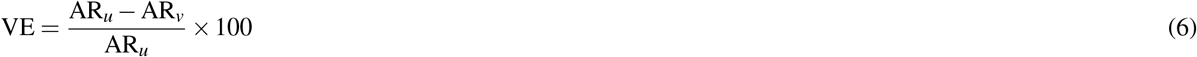

Where AR_*u*_ and AR_*u*_ are the attack rates of unvaccinated and vaccinated groups, respectively. The term *attack rate* refers to the fraction of the population considered at-risk (based on a chosen criterion such as death or severe illness) by the disease of interest during a specified time interval. Depending on the criterion used, the efficacy number varies. For instance, efficacy calculated for death will be different from that measured for a defined illness threshold.

Accurate estimates on efficacy data for the vaccines are still under active study. Initial data from Pfizer’s Phase 3 clinical trial, published in December 2020, listed the vaccine to hold efficacy of 95%. Subsequent data presented by the company in April 2021 had updated efficacy against COVID-19 to 91.3%, a measure of preventing symptomatic infection up to six months after the second dose. In terms of preventing severe disease as defined by the CDC guidelines, this vaccine was found to be 100%, and as per the FDA criterion of preventing severe disease, they reported efficacy equal to 95.3%. Other studies reported lower than 90 as the efficacy when measured after 6 months. However, all studies so far pointed to very high values of efficacy figures against severe diseases. The CDC studies revealed in August 2021 that mRNA vaccine immunity against infection might be waning, even though the vaccines stay highly effective against hospitalization and deaths. Citing the New York state data, the CDC showed that vaccine effectiveness against infection has dropped from 91.7% to 79.8%.

As for the other mRNA vaccine, somewhat similar data was reported in terms of its effectiveness. Initial data from Phase 3 reported 95% efficacy, which was later found to be 90% against infection, and 95% against critical illness when measured six months post vaccinations.

The reported efficacy of the Johnson & Johnson COVID-19 vaccine is slightly lower than the mRNA-based options. However, the overall efficacy number equal to 72% against mild illness and a higher value, 86% against severe disease are reported for this vaccine.

There is ongoing research assessing the efficacy figures of these vaccines over a different time duration. However, at this stage, the available research is still limited. Therefore, to make a concrete comparison of effectiveness between these drugs, more data is required.

Regarding the effectiveness of these vaccines against virus mutations, there is not much scientific evidence available to conclude it. Some studies claim that all three of these vaccines are less effective against the delta variants than the earlier versions of the virus. Despite these concerns on reduced effectiveness, as we shall see in section, the mortality statistics strongly suggest the superior benefit of vaccination even against the threats of delta variant^45,46^.

### 9.3 Impact of Vaccines

Vaccine roll-out in the United States began at a full scale around January 2021, at a time the weekly death count reached an all-time high figure of 26000 people. Initial administration of the vaccine was carried out by giving preference to healthcare staff, people who are older, and those identified to be at higher risk of disease complications. By the end of March, a total of about 107 million doses, predominantly the mRNA vaccines, have been administered in the country, which is about 32.25% of the US population. The impact of this was seen in the rapid reduction in the death count. In 3 months, when 30% of the population got vaccinated, the weekly death counts fell by 70%, as can be seen in Figure 35. With the increased vaccination rate, the number of deaths continued to drop until the middle of April 2021. The trajectory of vaccination rate has slowed down in April, which among other things, appears to have reflected in the death rates during April. During May-July, the vaccine access opened up, and when the rate of administration increased, the death rates also improved. By July 2021, the aggregate number of weekly deaths due to COVID-19 in the country was brought down to less than 2000, for the first time since April 2020, when the pandemic launched its first wave of assault in the United States. A visual explanation of this correlation between the rate of vaccine administration and its impact on controlling the death rates is presented in Figure 35. The weekly aggregated number of deaths in the US is plotted against the time window from January 2021 through the first week of July 2021. Color code maps to the percentage of people who are 65 years or older (recall that the vaccines were initially made available to this age group before opening it up for younger people). As shown in the illustration, a significant improvement in COVID-19 mortality happened when the vaccine rates for this age group progressed from 0 to 30%.

**Figure 35.**
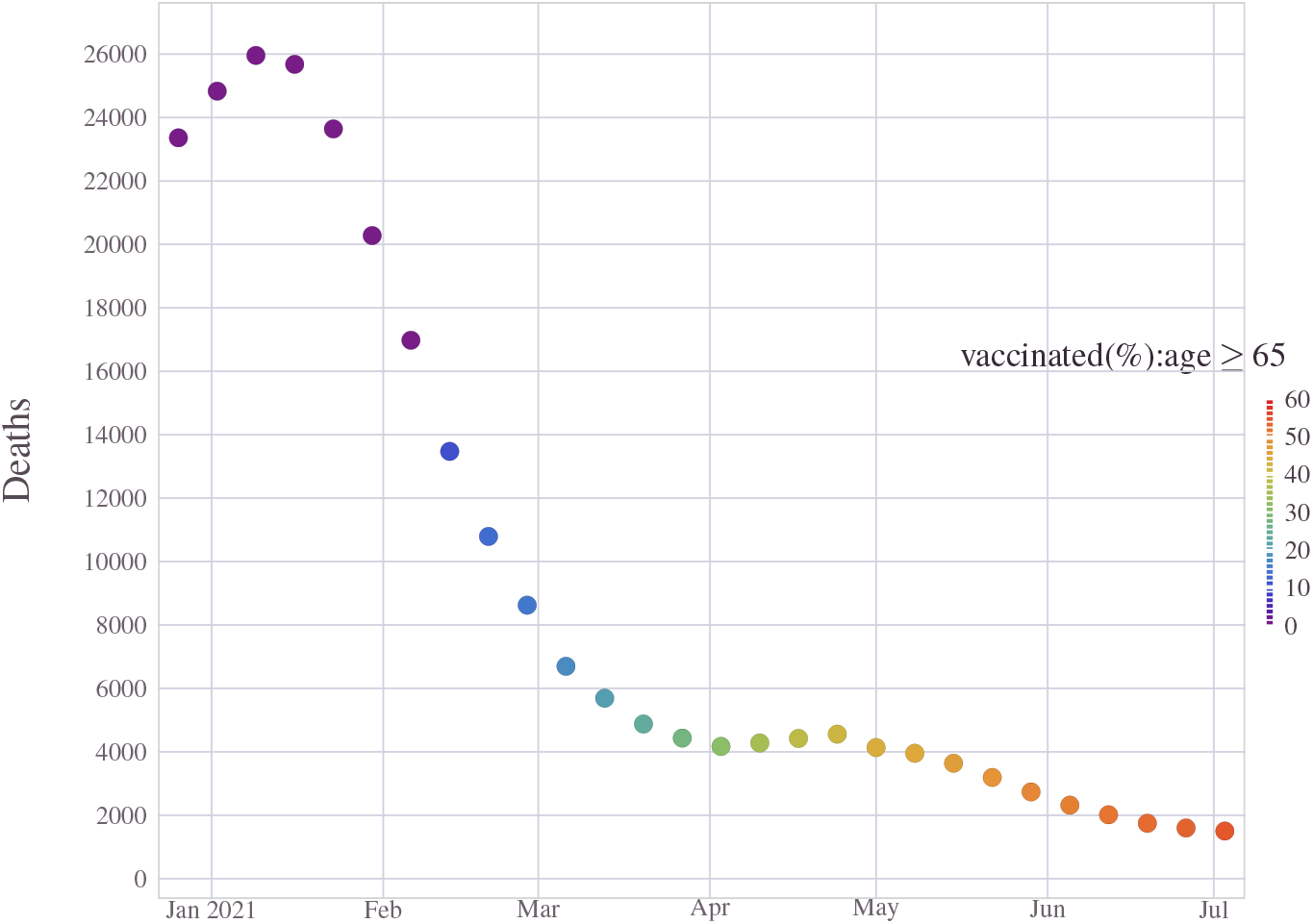
Vaccine administration began in early 2021 in a phased manner. By May 2021, access to the vaccine was widely available for individuals 16 years or older. As the vaccination rate increased, the death rates did fall considerably, as can be seen here. The color gradient indicates the fraction of people who are 65 years or older who are fully vaccinated.

With the roll-out of vaccines, and the steady decline in death counts, optimism surrounding the end of the pandemic grew brighter. However, just around the time when things were looking steady and in control, the threat of the delta variant emerged, which eventually triggered a fresh wave of the pandemic. The decision to relax restrictions and partial opening during the summer months also contributed to accelerating this wave. However, looking closely at the data, a major contributing factor for the surge in deaths during this wave was an inadequate level of vaccination.

During the three months from July through September, 100,000 people died around the country due to COVID-19 when the delta variant dominated the virus landscape in the United States. Most of these deaths are attributed to people who remained un-vaccinated when vaccines were plentiful and accessible to everyone 12 years or older. Despite the abundant supply of vaccines, the United States moved up the ranks to become the country with the highest rate of recent deaths.

We examined the correlation between vaccination rates as of July 2021 and death rates among adults (of age 18 and older) during the Delta surge (covering July 1, 2021, through October 10, 2021). In addition, we analyze the strong correlation between un-vaccinated states and the increased mortality percentage from the data available across states. A recent analysis^65^ brought up the distinct nature of the virus deaths during the Delta surge, compared to the earlier chapters of the pandemic. According to this study, “the people who died in the month of July-October were concentrated in the South of United States, a region that has lagged in vaccinations; many of the deaths were reported in Florida, Mississippi, Louisiana, and Arkansas. Moreover, those who died were younger: In August, every age group under 55 had its highest death toll of the pandemic”,^65^. Our analysis quantitatively validates this thesis. The correlation between vaccination rate among the eligible population and the cumulative deaths during the Delta variant induced wave-4 are pictorially depicted in Figure 36 and Figure 37.

**Figure 36.**
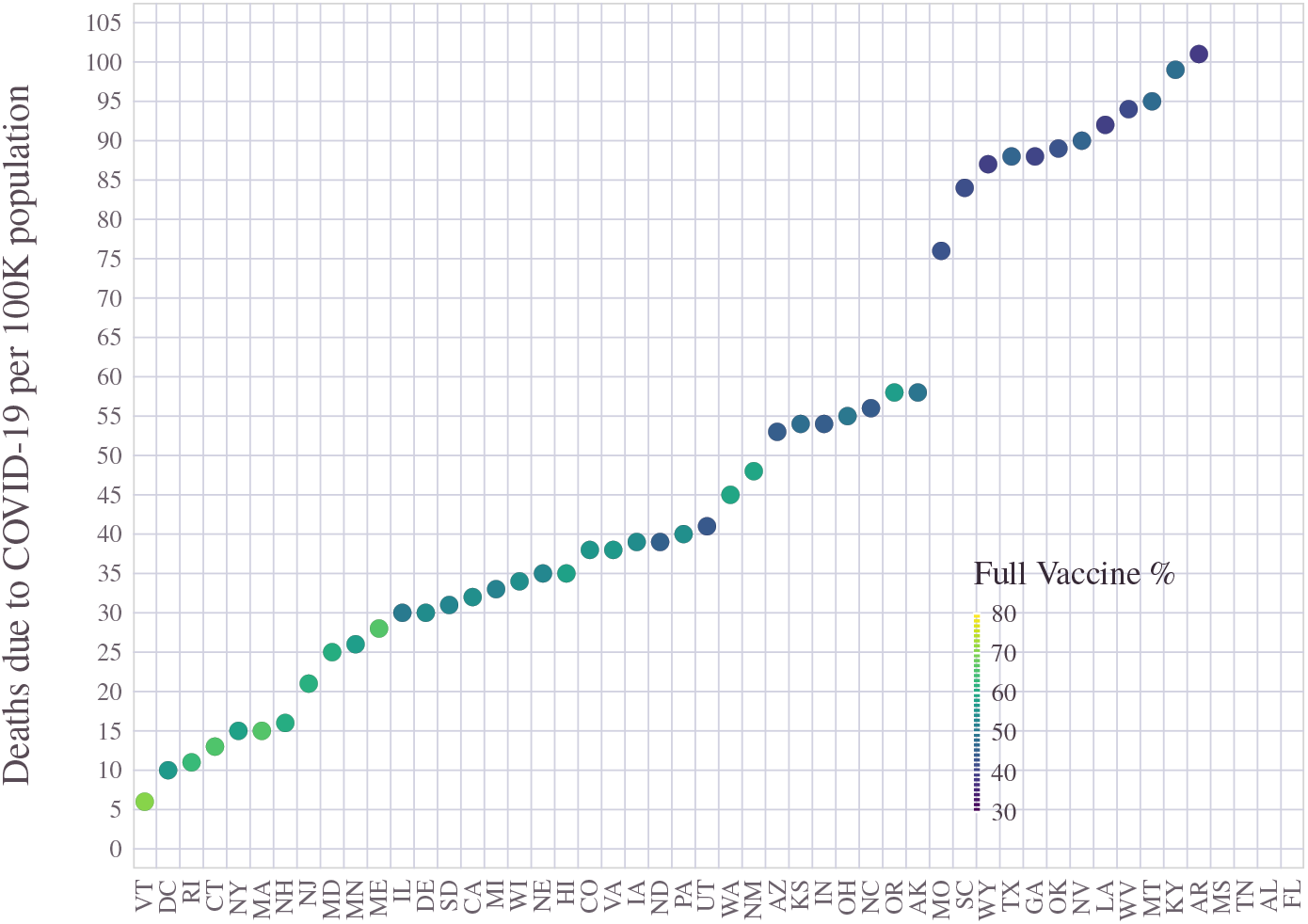
Vaccine impact during Delta variant. Correlation between vaccination rate and deaths across states during delta variant surge (wave-4). Color code indicates the percentage of people with age ≥ 12 who are fully vaccinated. Those states which lagged in vaccination have witnessed a higher proportion of deaths during the fourth wave of the pandemic.

**Figure 37.**
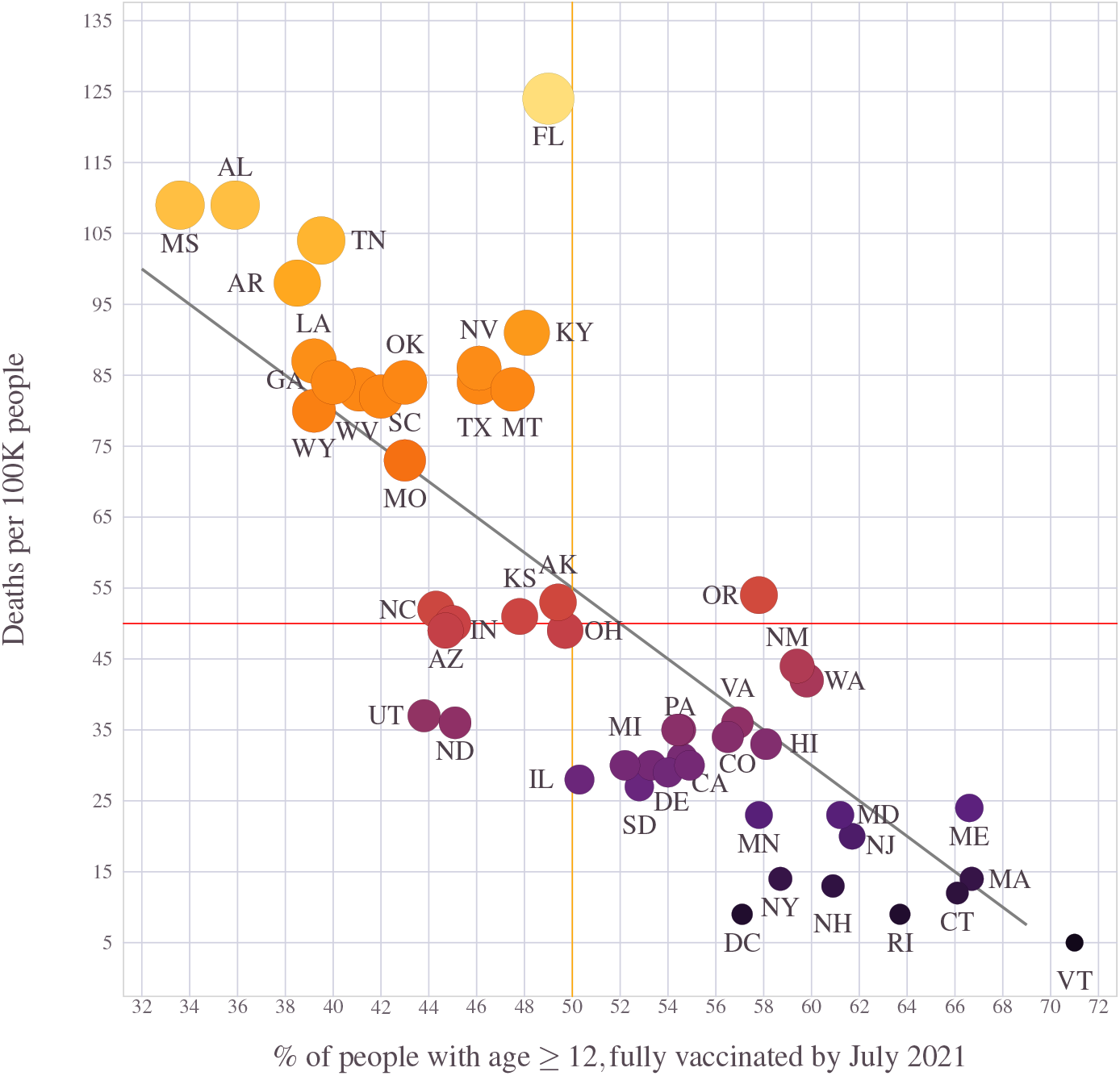
Vaccine impact during Delta variant. Correlation between vaccination rate and deaths across states during delta variant surge (wave-4). Darker color (or smaller marker size) indicates a proportionally lesser number of deaths by COVID-19 during the delta variant surge in wave-4. Those states which lagged in vaccination have witnessed a higher proportion of deaths during the fourth wave of the pandemic.

**Figure 38.**
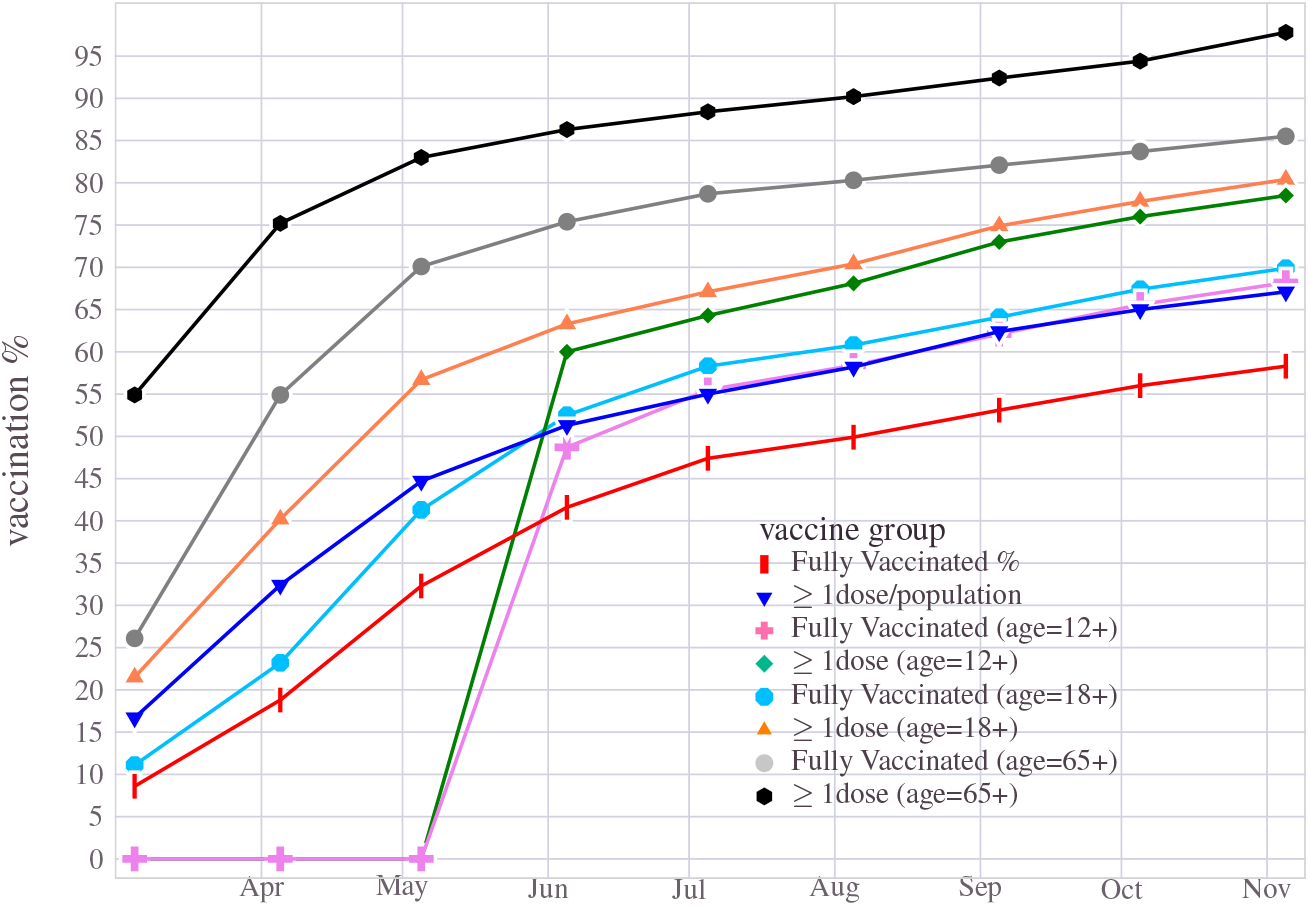
Vaccine deployment and progress over time. In 6 months over 50 percent of the population got vaccinated

## 10 Temporal dynamics of the COVID-19 pandemic

One of the fascinating aspects of the COVID-19 pandemic is its degree of dynamism as it evolved. As the virus underwent mutations, it traveled geographically, found new homes, triggered new waves, and marched on. In this section, we discuss how the pandemic’s demographic characteristics changed over its journey as it traversed across the U.S geographical landscape.

### 10.1 Agewise statistics over time

The COVID-19 induced deaths in the American population across different age groups exhibited a remarkable statistical consistency over a long time. Until the vaccines arrived at the scene, both the absolute number of deaths and age-wise distribution behaved monotonically from younger to the older section of the U.S populace. When the vaccine administration gathered momentum, post-January 2021, a drastic decline in death counts across all age groups was noticed. However, the situation changed around August 2021 with the emergence of the delta variant, triggering the fourth pandemic surge. A more significant than expected number of younger individuals succumbed to the disease during this period, tilting from the age trend hitherto witnessed. During the latest wave, the most significant chunk of deaths came from people in the 65 − 74 age group, along with a substantial increase in mortality among the younger adults belonging to the 25 − 54 years old category. This is pictorially illustrated in Figure 40, where the absolute number of monthly deaths by age cohort is shown. The percentage share of deaths distributed across age groups in Figure 39 indicates the relative change in the age trend before and during wave-4. In December 2020, while at the peak of Wave-3, the ≥ 75 years old people had accounted for 31.5% of the total deaths, which dropped to 17% share when it came to September 2021 during wave-4. All across the age groups below 75 years old registered an increase in percentage share of deaths during the delta variant surge. 22.5% of the September deaths from COVID-19 reported are from people between 55 − 74 years old, an increase of 40% from their corresponding figures from the wave-3 peak. The younger adults (age between 25 and 54), on the other hand, ended up with a 3 times higher percentage share of deaths during delta variant wave than what it was during wave-3. This particular cohort had a cumulative share of 3.5% in December 2020, which increased to a high of 10.5% in September 2021.

**Figure 39.**
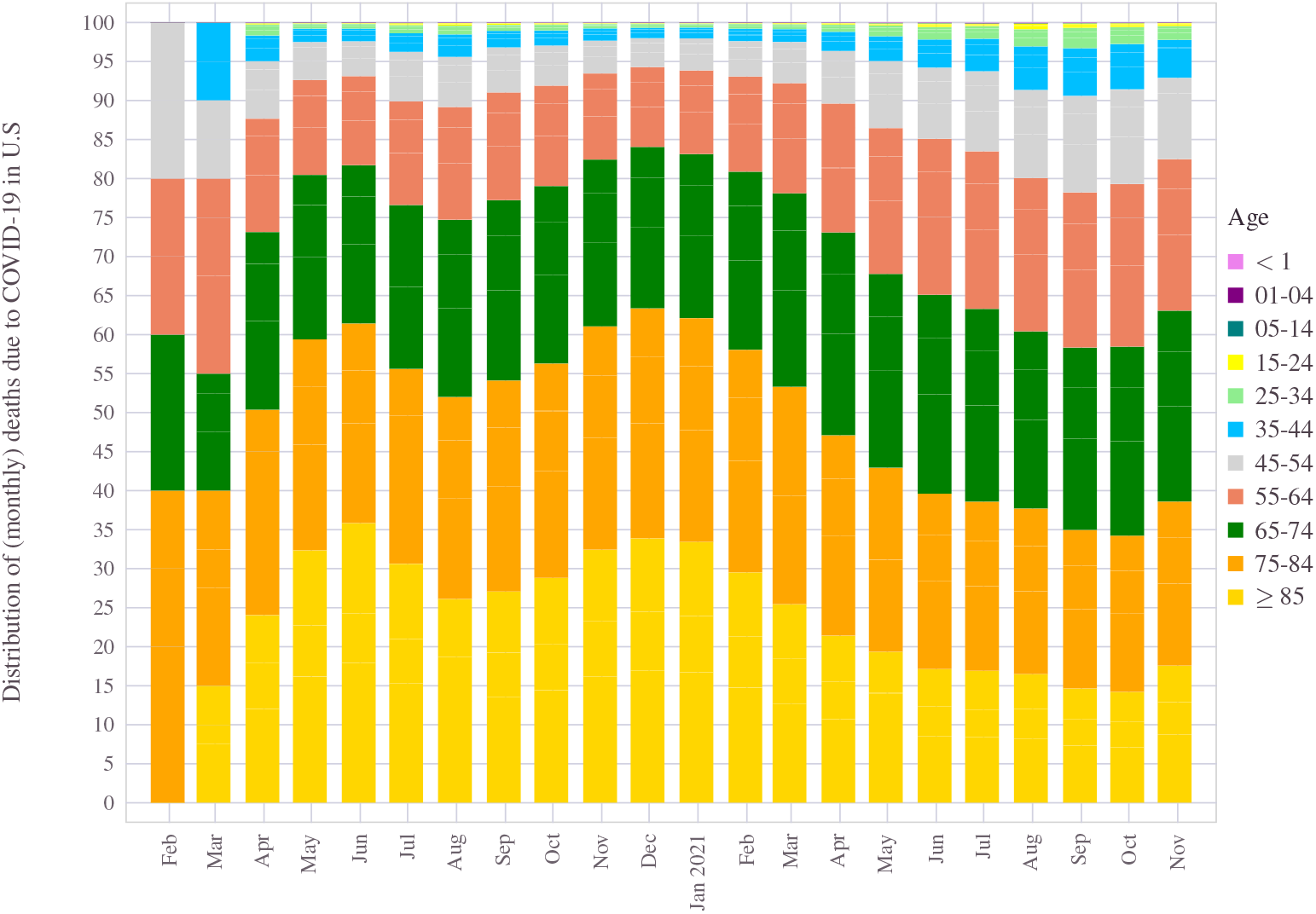
Vaccine impact: Change in the distribution of deaths across age profiles over time. Since the deployment of the vaccine, a noticeable improvement was seen in the mortality rates of oldest adults and gradually moving towards an equal distribution of deaths among broader sets of adult age groups.

**Figure 40.**
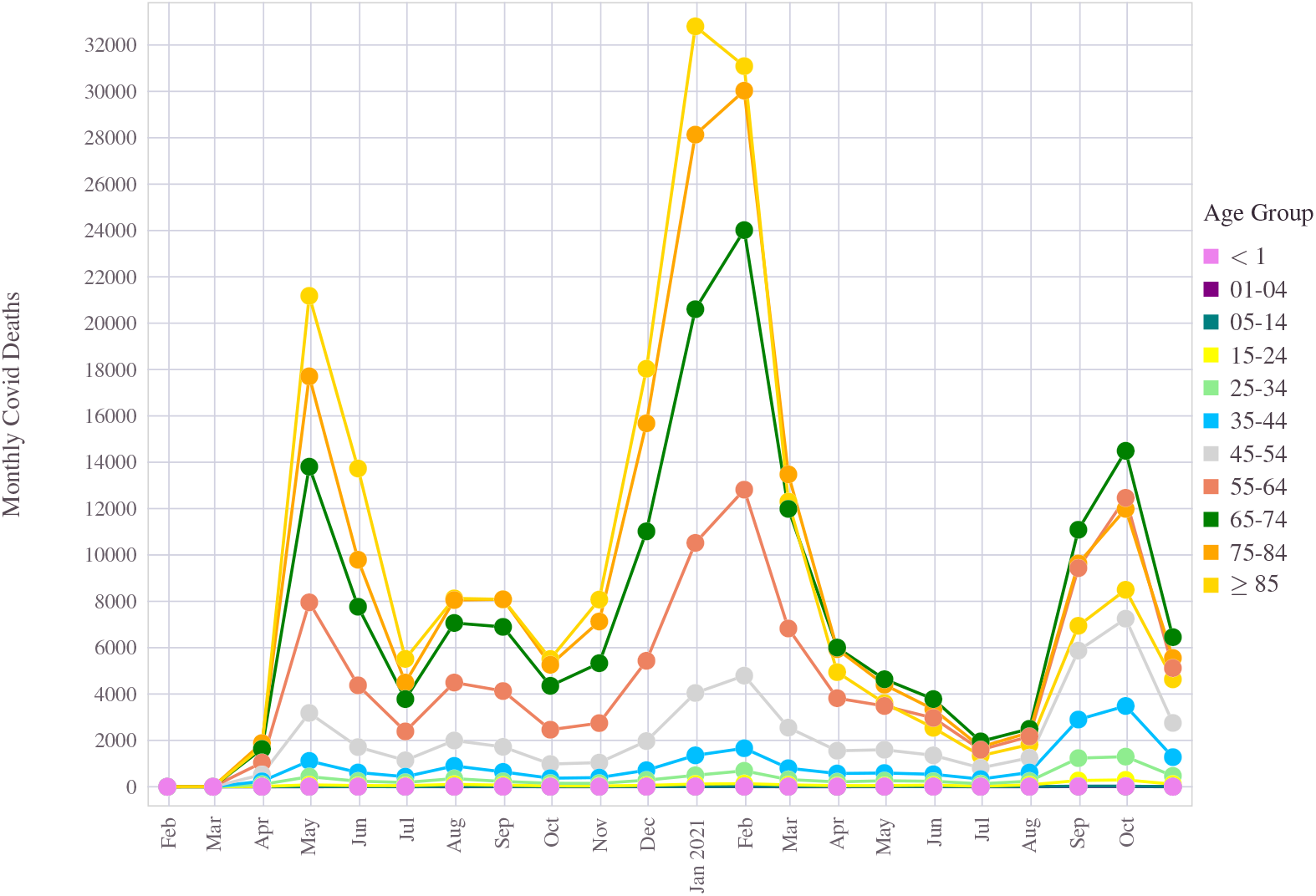
Monthly aggregate number of deaths in U.S due to the COVID-19 pandemic. Age-wise numbers are shown.

### 10.2 Gender statistics over time

Overall, from the American population, more males than females have died due to COVID-19. This phenomenon largely stayed the same for any short or long period, sliced during the last 22 months, as shown in Figure 41. Although the relative standing on the absolute death counts between the two gender (male/female deaths) has varied occasionally over the pandemic’s life span, the ratio almost always remained to be no less than 1. Even during the delta variant surge, this trend continued.

**Figure 41.**
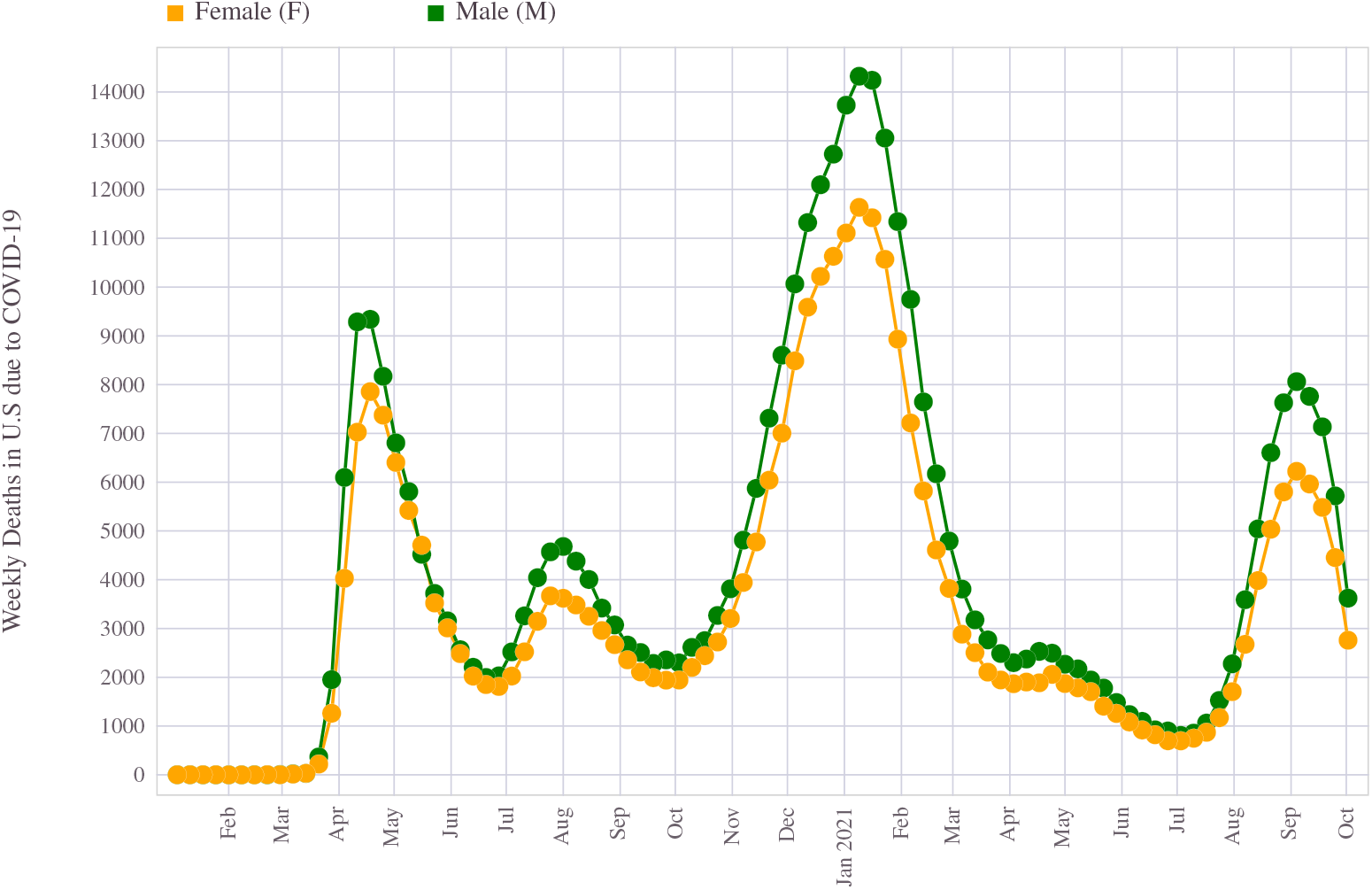
Gender-wise mortality figures from COVID-19: The cumulative weekly number of deaths classified into male/female throughout the pandemic’s timeline is shown.

### 10.3 Ethnicity behavior over time

The relative positions of the different ethnic communities also stayed somewhat the same throughout the pandemic. In terms of absolute aggregate death counts of the country, the white individuals accounted for a higher number of deaths, followed by Hispanic people and then the Black community. During the first wave, from March to July 2020, more ethnic black people than those of the Hispanic race have died from COVID-19 disease. For the black community, this initial outbreak triggered the highest weekly deaths that were ever registered during the entire pandemic. At the height of this initial surge, as many as 4100 black lives were lost to the virus. The second wave and the giant tide during the latter half of 2020 consumed the most considerable amount of people from all ethnic communities. The Hispanic people stood second, behind the Non-Hispanic White population in terms of the most significant number of registered deaths during this period. The Delta wave resulted in an almost equal number of Black and Non-Hispanic people when the White cross-section of the society had to brace twice as many deaths as all other races combined. Figure 42 depicts the relative comparison of absolute death counts per week as the outbreak advanced in time and thus creating these death mountains.

**Figure 42.**
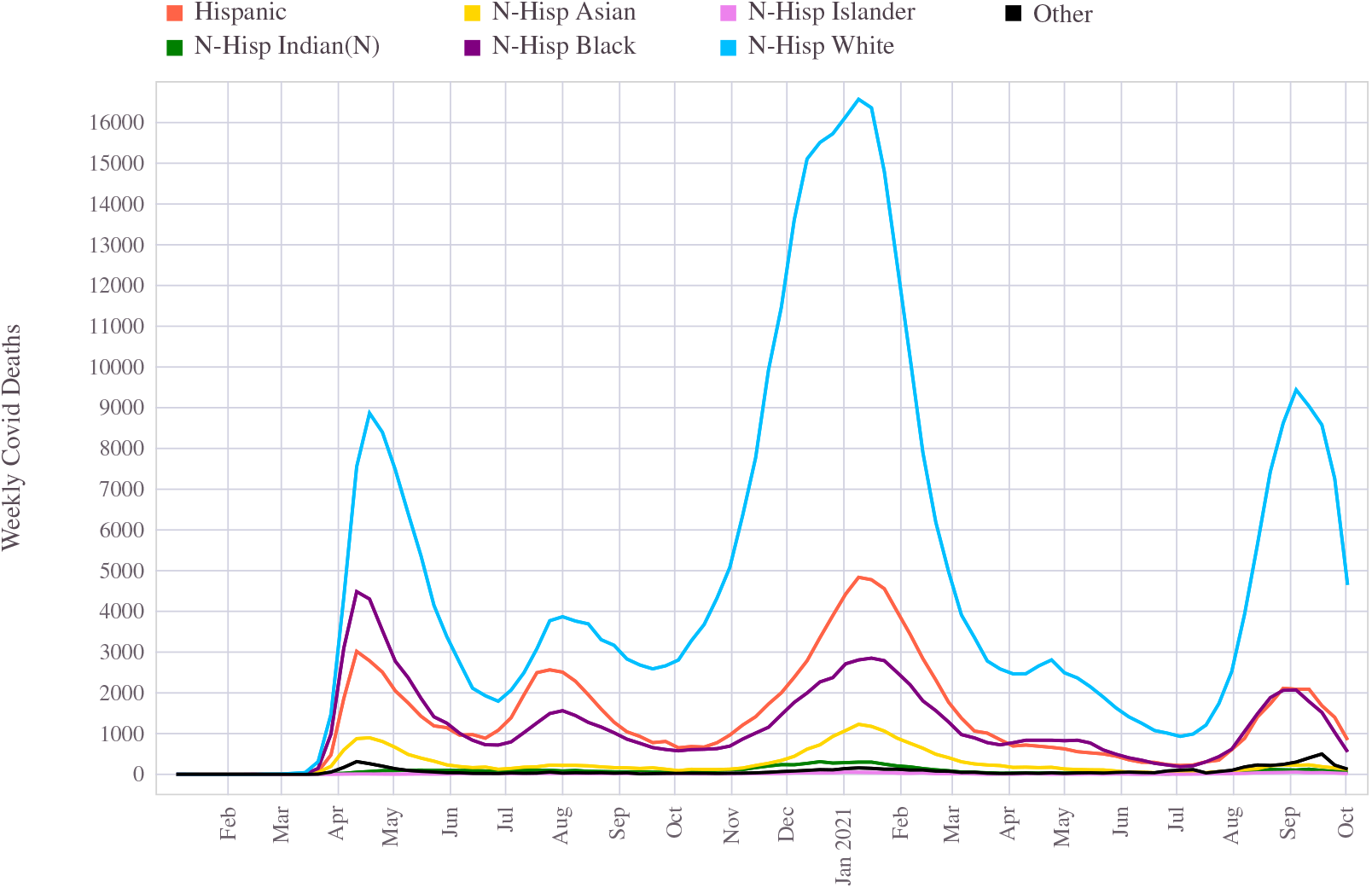
Trend in weekly death counts from COVID-19 for each race/ethnic group in U.S

### 10.4 Temporal view of COVID-19 deaths mountain in states

At different epochs, the COVID-19 virus has found different homes. Even though it all began in Washington State, the initial wave wreaked havoc on the east coast of the United States. New York, New Jersey, Pennsylvania were among the most hit states between January to June 2020. This is reflected in the Figure 43. During that period, New York City, in particular, was the epicenter of the pandemic, with the weekly deaths soured to the highest level during mid-April 2020 with as many as 4700 people fallen. During the same window, New Jersey accrued its highest peak with 2500 deaths per week. Although the west coast of the United States, including its biggest state California, was hit by the pandemic, the scale of the situation was much lower than what was felt at the eastern front.

**Figure 43.**
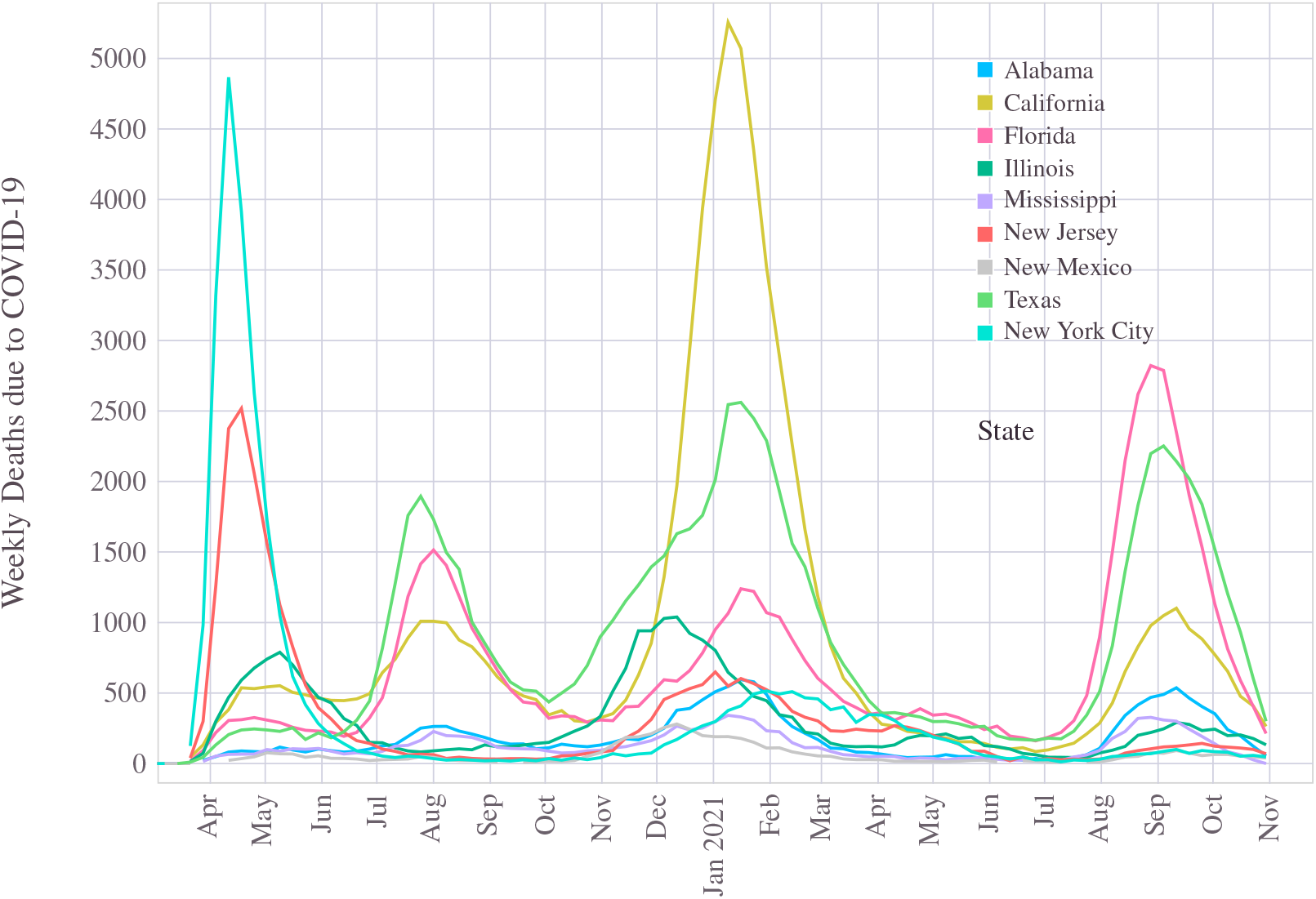
The COVID-19 pandemic waves in selected states of the U.S: The surge in cases and their timeline have varied across different states.

The second wave was relatively mild compared to the first. During this stage of the outbreak, a larger number of cases and deaths were reported from the bigger states, namely, Texas and Florida, followed by California.

The holiday season into the end of the year 2020 and through the new year of 2021 saw the pandemic at its full vigor, presenting itself with a massive death mountain which aggregated to 26000 deaths in a single week across the United States. As many as 20% of those died accounted in California alone. The large states California, Texas, and Florida, yet again were heavily impacted during this phase.

Thanks to the vaccine roll-out, the situation began to improve until the fourth wave, which coincided with the arrival of the delta variant. This leg of the pandemic, which started its ascend in July through November 2021, saw a weekly peak level of 10000 deaths in September 2021. Florida, in particular, was struck and, together with Texas, has dominated the death counts in the Delta variant surge. Many southern states are also impacted during the fourth surge. Although California had a much lesser impact than it had during the third wave, it stood as the third worst-hit state in this period on the absolute number of deaths.

## 11 Concluding Remarks

This paper analyzed the COVID-19 situation in the United States and brought up several exciting mortality patterns using empirical data evidence. A closer look at the data helped us reveal several hidden trends that have impacted specific demographics in a dis-proportional way. We have introduced new metrics to quantify better some of these hidden patterns, especially the age-ethnicity correlation. We have found that higher population density and old age population strongly correlate with high mortality figures. In addition, discussions on connections between COVID-19 and other respiratory diseases showed some statistical similarities and differences. Finally, vaccines’ role in controlling the pandemic to a manageable situation and how well the mortality rate followed the vaccine administration are discussed in detail.

The pandemic has been active for nearly 2 years, the dynamics of which have considerably varied during the last several months. Nevertheless, we have covered some of the specific temporal dynamics of the COVID-19 outbreak in reasonable detail. We believe that the statistical inference such as the ones gathered in this paper would be helpful for better scientific understanding, policy preparation and thus adequately prepare should a similar situation arise in the future.

## 12 Limitations and Notes on the Data sources

For much of the work, we have relied on the publicly available data from CDC^26,27^. Some of these data are live and constantly updated, thus subject to specific errors. For example, the death estimates are provisional, as the final mortality data after validation gets released only later. The population data for the U.S is sourced from the U.S Census 2020. The exact number is likely to have changed during the pandemic, and thus the normalized figures quoted are subject to such variations. In the computation of mortality metrics, both the death counts and the number of infected cases are required. Usually, there is a delay between these counts, which is not addressed in our metric computations. Although these caveats do not change the overall conclusions from our analysis, we wish to point out that an accurate assessment will need to address these aspects. While the tools developed for this work is extendable to future data, the ones analyzed here are based on the available data up to 12 November 2021.

## 13 Reproducible research

We strongly believe in the philosophy of faithful reproducible research and open sharing of scientific data^84^. The data used in this work is all from publicly available verified sources. Therefore, all of the results, including the figures and statistical measures in this paper, are accurately reproducible. We have used Julia Programming Language (and several scientific libraries in Julia ecosystem such as LinearAlgebra, DataFrames, Statistics, DifferentialEquations, JuMP, Gadfly, Plots, Query, CSV, Pluto, and PlutoUI)^85,86^ for the majority of the analysis in this paper.

## Data Availability

Julia/Pluto notebook is open source. Plan to upload the source code to GitHub
Available any time

## Acknowledgements

The work on this began in the summer 2020 as a side project while attempting to develop a mathematical and computing framework using a delay differential equation-based model of the COVID-19 pandemic. Part of the mathematical modeling work was submitted and presented at the Greater San Diego Science and Engineering Fair 2021. We appreciate the guidance and support, and encouragement received from the San Diego science committee. This work’s bulk of computational and statistical analysis is performed using Julia’s remarkable scientific programming ecosystem. The online Julia community has been a valuable go-to place for getting all help related to Julia. Summer research at UCSD and helpful guidance from Dr. Valentina Kouznetsova and Dr. Igor Tsigelny have spurred my interest in research and the joy of scientific presentation. Professor Joshua Botica has helped me with statistical analysis and Professor John Harland in clarifying Linear Algebra related topics at Palomar college. Many thanks to my friends Sumana Srinivas at U.C Berkeley and Anjali Koganti at UCLA for several helpful suggestions in getting this paper to a much better shape than it originally was. Ms. Buehler at Del Norte HS has been my source of constant encouragement for much of the last two years, and many thanks to her for encouraging me to write this up.

## Footnotes

1 There are many different types of corona-viruses. Some of them cause colds or other mild respiratory illnesses; others can trigger serious diseases, including Severe Acute Respiratory Syndrome (SARS) and the Middle East respiratory syndrome (MERS). For example, the SARS outbreak in 2003-2004 affected a significant population in several countries. The coronavirus that causes COVID-19 has many similarities to the one that caused the 2003 SARS outbreak. While many aspects of these viruses are still unknown, the SARS-CoV-2 spread faster than the 2003 SARS-CoV-1 virus

## References

1. Ksiazek TG, et al. 2003. A novel coronavirus associated with severe acute respiratory syndrome. N Engl J Med 348: 1953–1966.

2. Taubenberger, J. K., Kash, J. C., & Morens, D. M. (2019). The 1918 influenza pandemic: 100 years of questions answered and unanswered. Science Translational Medicine, 11(502). https://doi.org/10.1126/scitranslmed.aau5485

3. Morens DM, Breman JG, Calisher CH, Doherty PC, Hahn BH, Keusch GT, Kramer LD, LeDuc JW, Monath TP, Taubenberger JK. The Origin of COVID-19 and Why It Matters. Am J Trop Med Hyg. 2020 Sep;103(3):955–959. doi: 10.4269/ajtmh.20-0849. PMID: 32700664; PMCID: PMC7470595.

4. Guo YR, Cao QD, Hong ZS, Tan YY, Chen SD, Jin HJ, Tan KS, Wang DY, Yan Y. The origin, transmission and clinical therapies on coronavirus disease 2019 (COVID-19) outbreak - an update on the status. Mil Med Res. 2020 Mar 13;7(1):11. doi: 10.1186/s40779-020-00240-0. PMID: 32169119; PMCID: PMC7068984.

5. Zhou P, Yang XL, Wang XG, Hu B, Zhang L, Zhang W, Si HR, Zhu Y, Li B, Huang CL, Chen HD, Chen J, Luo Y, Guo H, Jiang RD, Liu MQ, Chen Y, Shen XR, Wang X, Zheng XS, Zhao K, Chen QJ, Deng F, Liu LL, Yan B, Zhan FX, Wang YY, Xiao GF, Shi ZL. A pneumonia outbreak associated with a new coronavirus of probable bat origin. Nature. 2020 Mar;579(7798):270–273. doi: 10.1038/s41586-020-2012-7. Epub 2020 Feb 3. PMID: 32015507; PMCID: PMC7095418.

6. Worldometer. COVID-19 coronavirus pandemic. Data retrieved: 12 November, 2021. https://www.worldometers.info/coronavirus/

7. Lucas Rodes-Guirao Cameron Appel Charlie Giattino Esteban Ortiz-Ospina Joe Hasell Bobbie Macdonald Diana Beltekian Hannah Ritchie,Edouard Mathieu and Max Roser. Coronavirus pandemic (covid-19).OurWorld in Data, 2020. https://ourworldindata.org/coronavirus.

8. Cucinotta D, Vanelli M. WHO Declares COVID-19 a Pandemic. Acta Biomed. 2020 Mar 19;91(1):157–160. doi: 10.23750/abm.v91i1.9397. PMID: 32191675; PMCID: PMC7569573.

9. Vellas C, Delobel P, de Souto Barreto P, Izopet J. COVID-19, Virology and Geroscience: A Perspective. J Nutr Health Aging. 2020;24(7):685–691. doi: 10.1007/s12603-020-1416-2. PMID: 32744561; PMCID: PMC7301052.

10. Pizano-Escalante MG, Anaya-Esparza LM, Nuño K, Rodríguez-Romero JJ, Gonzalez-Torres S, López-de la Mora DA, Villagrán Z. Direct and Indirect Effects of COVID-19 in Frail Elderly: Interventions and Recommendations. J Pers Med. 2021 Oct 2;11(10):999. doi: 10.3390/jpm11100999. PMID: 34683141; PMCID: PMC8539433.

11. Hussien H, Nastasa A, Apetrii M, Nistor I, Petrovic M, Covic A. Different aspects of frailty and COVID-19: points to consider in the current pandemic and future ones. BMC Geriatr. 2021 Jun 27;21(1):389. doi: 10.1186/s12877-021-02316-5. PMID: 34176479; PMCID: PMC8236311.

12. Dai M, Liu D, Liu M, Zhou F, Li G, Chen Z, Zhang Z, You H, Wu M, Zheng Q, Xiong Y, Xiong H, Wang C, Chen C, Xiong F, Zhang Y, Peng Y, Ge S, Zhen B, Yu T, Wang L, Wang H, Liu Y, Chen Y, Mei J, Gao X, Li Z, Gan L, He C, Li Z, Shi Y, Qi Y, Yang J, Tenen DG, Chai L, Mucci LA, Santillana M, Cai H. Patients with Cancer Appear More Vulnerable to SARS-CoV-2: A Multicenter Study during the COVID-19 Outbreak. Cancer Discov. 2020 Jun;10(6):783–791. doi: 10.1158/2159-8290.CD-20-0422. Epub 2020 Apr 28. PMID: 32345594; PMCID: PMC7309152.

13. Liu C, Zhao Y, Okwan-Duodu D, Basho R, Cui X. COVID-19 in cancer patients: risk, clinical features, and management. Cancer Biol Med. 2020 Aug 15;17(3):519–527. doi: 10.20892/j.issn.2095-3941.2020.0289. PMID: 32944387; PMCID: PMC7476081.

14. Erener S, “Diabetes, infection risk and COVID-19”, Mol Metab, 2020 Sep;39:101044. doi:10.1016/j.molmet.2020.101044. Epub 2020 Jun 23. PMID: 32585364; PMCID: PMC7308743.

15. Azevedo RB, Botelho BG, Hollanda JVG, Ferreira LVL, Junqueira de Andrade LZ, Oei SSML, Mello TS, Muxfeldt ES. Covid-19 and the cardiovascular system: a comprehensive review. J Hum Hypertens. 2021 Jan;35(1):4–11. doi: 10.1038/s41371-020-0387-4. Epub 2020 Jul 27. PMID: 32719447; PMCID: PMC7384729.

16. Gutierrez L, Beckford J, Alachkar H. Deciphering the TCR Repertoire to Solve the COVID-19 Mystery. Trends Pharmacol Sci. 2020 Aug;41(8):518–530. doi: 10.1016/j.tips.2020.06.001. Epub 2020 Jun 20. PMID: 32576386; PMCID: PMC7305739.

17. Lipsitch M, Dean NE. Understanding COVID-19 vaccine efficacy. Science. 2020 Nov 13;370(6518):763–765. doi: 10.1126/science.abe5938. Epub 2020 Oct 21. PMID: 33087460.

18. Cascella M, Rajnik M, Aleem A, Dulebohn SC, Di Napoli R. Features, Evaluation, and Treatment of Coronavirus (COVID-19). 2021 Sep 2. In: StatPearls [Internet]. Treasure Island (FL): StatPearls Publishing; 2021 Jan–. PMID: 32150360.

19. Jackson LA, Anderson EJ, Rouphael NG, Roberts PC, Makhene M, Coler RN, McCullough MP, Chappell JD, Denison MR, Stevens LJ, Pruijssers AJ, McDermott A, Flach B, Doria-Rose NA, Corbett KS, Morabito KM, O’Dell S, Schmidt SD, Swanson PA 2nd, Padilla M, Mascola JR, Neuzil KM, Bennett H, Sun W, Peters E, Makowski M, Albert J, Cross K, Buchanan W, Pikaart-Tautges R, Ledgerwood JE, Graham BS, Beigel JH; mRNA-1273StudyGroup. An mRNA Vaccine against SARS-CoV-2 - Preliminary Report. N Engl J Med. 2020 Nov 12;383(20):1920–1931. doi: 10.1056/NEJMoa2022483. Epub 2020 Jul 14. PMID: 32663912; PMCID: PMC7377258.

20. Park CL, Russell BS, Fendrich M, Finkelstein-Fox L, Hutchison M, Becker J. Americans’ COVID-19 Stress, Coping, and Adherence to CDC Guidelines. J Gen Intern Med. 2020 Aug;35(8):2296–2303. doi: 10.1007/s11606-020-05898-9. Epub 2020 May 29. PMID: 32472486; PMCID: PMC7259430.

21. Bertsekas DP and Tsitsiklis JN, Introduction to probability, Athena Scientific, 2000.

22. Ruprecht MM, Wang X, Johnson AK, Xu J, Felt D, Ihenacho S, Stonehouse P, Curry CW, DeBroux C, Costa D, Phillips Ii G. Evidence of Social and Structural COVID-19 Disparities by Sexual Orientation, Gender Identity, and Race/Ethnicity in an Urban Environment. J Urban Health. 2021 Feb;98(1):27–40. doi: 10.1007/s11524-020-00497-9. Epub 2020 Dec 1. PMID: 33259027; PMCID: PMC7706696.

23. Mortality Frequency Measures, Center for Disease Control and Prevention (CDC) https://www.cdc.gov/csels/dsepd/ss1978/lesson3/section3.html

24. Wise RP, Livengood JR, Berkelman RL, Goodman RA. Methodologic alternatives for measuring premature mortality. Am J Prev Med 1988;4:268–273.

25. The New York Times, “A Continent Where the Dead Are Not Counted “, https://www.nytimes.com/2021/01/02/world/africa/africa-coronavirus-deaths-underreporting.html, Date accessed: 02 November 2021.

26. Centers for Disease Control and Prevention (CDC). https://data.cdc.gov

27. Centers for Disease Control and Prevention. COVID Data Tracker https://covid.cdc.gov/covid-data-tracker/#datatracker-home

28. “COVID-19 Data Repository by the Center for Systems Science and Engineering (CSSE) at Johns Hopkins University”, url: https://github.com/CSSEGISandData/COVID-19.

29. Dong E, Du H, Gardner L. An interactive web-based dashboard to track COVID-19 in real time. Lancet Inf Dis. 20(5):533–534. doi:10.1016/S1473-3099(20)30120-1”

30. Ahmad FB, Cisewski JA, Miniño A, Anderson RN. Provisional Mortality Data — United States, 2020. MMWR Morb Mortal Wkly Rep 2021;70:519–522. DOI:http://dx.doi.org/10.15585/mmwr.mm7014e1

31. Erratum: Vol. 70, No. 14. MMWR Morb Mortal Wkly Rep 2021;70:900. DOI: http://dx.doi.org/10.15585/mmwr.mm7024a4

32. Gundlapalli AV, Lavery AM, Boehmer TK, et al. Death Certificate–Based ICD-10 Diagnosis Codes for COVID-19 Mortality Surveillance — United States, January–December 2020. MMWR Morb Mortal Wkly Rep 2021;70:523–527. DOI:http://dx.doi.org/10.15585/mmwr.mm7014e2

33. Spinney, L. (2017). Pale Rider: The Spanish Flu of 1918 and How It Changed the World. New York, NY: Public Affairs.

34. Morens, D. M., Taubenberger, J. K., and Fauci, A. S. (2009). The persistent legacy of the 1918 influenza virus. N. Engl. J. Med. 361, 225–229. doi: 10.1056/NEJMp0904819

35. Morens, D. M., and Fauci, A. S. (2007). The 1918 influenza pandemic: insights for the 21st century. J. Infect. Dis. 195, 1018–1028. doi: 10.1086/511989

36. Khan MS, Shahid I, Anker SD, Solomon SD, Vardeny O, Michos ED, Fonarow GC, Butler J. Cardiovascular implications of COVID-19 versus influenza infection: a review. BMC Med. 2020 Dec 18;18(1):403. doi: 10.1186/s12916-020-01816-2. PMID: 33334360; PMCID: PMC7746485.

37. Flerlage T, Boyd DF, Meliopoulos V, Thomas PG, Schultz-Cherry S. Influenza virus and SARS-CoV-2: pathogenesis and host responses in the respiratory tract. Nat Rev Microbiol. 2021 Jul;19(7):425–441. doi: 10.1038/s41579-021-00542-7. Epub 2021 Apr 6. PMID: 33824495; PMCID: PMC8023351.

38. Pinho-Gomes A, Peters S, Thompson K, et al, “Where are the women? Gender inequalities in COVID-19 research authorship”, BMJ Global Health 2020;5:e002922.

39. National Academies of Sciences, Engineering, and Medicine. 2021. The Impact of COVID-19 on the Careers of Women in Academic Sciences, Engineering, and Medicine. Washington, DC: The National Academies Press. https://doi.org/10.17226/26061.

40. Gill K, Minall L, Nassif AR. pDNA and mRNA vaccines. InPractical Aspects of Vaccine Development 2022 Jan 1 (pp. 157–205). Academic Press. https://www.sciencedirect.com/science/article/pii/B9780128143575000076.

41. Marian AJ. Current state of vaccine development and targeted therapies for COVID-19: impact of basic science discoveries. Cardiovasc Pathol. 2021 Jan-Feb;50:107278. doi: 10.1016/j.carpath.2020.107278. Epub 2020 Sep 2. PMID: 32889088; PMCID: PMC7462898.

42. Le TT, Cramer JP, Chen R, Mayhew S. Evolution of the COVID-19 vaccine development landscape. Nat Rev Drug Discov. 2020 Oct;19(10):667–668. doi:10.1038/d41573-020-00151-8. PMID: 32887942.

43. Oliver SE, Gargano JW, Marin M, Wallace M, Curran KG, Chamberland M, McClung N, Campos-Outcalt D, Morgan RL, Mbaeyi S, Romero JR, Talbot HK, Lee GM, Bell BP, Dooling K. The Advisory Committee on Immunization Practices’ Interim Recommendation for Use of Moderna COVID-19 Vaccine - United States, December 2020. MMWR Morb Mortal Wkly Rep. 2021 Jan 1;69(5152):1653–1656. doi: 10.15585/mmwr.mm695152e1. PMID: 33382675.

44. Oliver SE, Gargano JW, Marin M, Wallace M, Curran KG, Chamberland M, McClung N, Campos-Outcalt D, Morgan RL, Mbaeyi S, Romero JR, Talbot HK, Lee GM, Bell BP, Dooling K. The Advisory Committee on Immunization Practices’ Interim Recommendation for Use of Pfizer-BioNTech COVID-19 Vaccine - United States, December 2020. MMWR Morb Mortal Wkly Rep. 2020 Dec 18;69(50):1922–1924. doi: 10.15585/mmwr.mm6950e2. PMID: 33332292; PMCID: PMC7745957.

45. Polack FP, Thomas SJ, Kitchin N, Absalon J, Gurtman A, Lockhart S, Perez JL, Pérez Marc G, Moreira ED, Zerbini C, Bailey R, Swanson KA, Roychoudhury S, Koury K, Li P, Kalina WV, Cooper D, Frenck RW Jr, Hammitt LL, Türeci Ö, Nell H, Schaefer A, Ünal S, Tresnan DB, Mather S, Dormitzer PR, S. ahin U, Jansen KU, Gruber WC; C4591001 Clinical Trial Group. Safety and Efficacy of the BNT162b2 mRNA Covid-19 Vaccine. N Engl J Med. 2020 Dec 31;383(27):2603–2615. doi: 10.1056/NEJMoa2034577. Epub 2020 Dec 10. PMID: 33301246; PMCID: PMC7745181.

46. Baden LR, El Sahly HM, Essink B, Kotloff K, Frey S, Novak R, Diemert D, Spector SA, Rouphael N, Creech CB, McGettigan J, Khetan S, Segall N, Solis J, Brosz A, Fierro C, Schwartz H, Neuzil K, Corey L, Gilbert P, Janes H, Follmann D, Marovich M, Mascola J, Polakowski L, Ledgerwood J, Graham BS, Bennett H, Pajon R, Knightly C, Leav B, Deng W, Zhou H, Han S, Ivarsson M, Miller J, Zaks T; COVE Study Group. Efficacy and Safety of the mRNA-1273 SARS-CoV-2 Vaccine. N Engl J Med. 2021 Feb 4;384(5):403–416. doi: 10.1056/NEJMoa2035389. Epub 2020 Dec 30. PMID: 33378609; PMCID: PMC7787219.

47. Asselah T, Durantel D, Pasmant E, Lau G, Schinazi RF. COVID-19: Discovery, diagnostics and drug development. J Hepatol. 2021 Jan;74(1):168–184. doi: 10.1016/j.jhep.2020.09.031. Epub 2020 Oct 8. PMID: 33038433; PMCID: PMC7543767.

48. Jackson, N.A.C., Kester, K.E., Casimiro, D. et al. The promise of mRNA vaccines: a biotech and industrial perspective. npj Vaccines 5, 11 (2020). https://doi.org/10.1038/s41541-020-0159-8

49. Lurie, N., Saville, M., Hatchett, R., & Halton, J. (2020). Developing Covid-19 Vaccines at Pandemic Speed. New England Journal of Medicine, 382(21), 1969–1973. https://doi.org/10.1056/nejmp2005630

50. U.S. Census Bureau, “U.S. and World Population Clock”, https://www.census.gov/popclock/. Accessed, Nov 10 2021.

51. Christine Tamir, “The Growing Diversity of Black America”, Pew Research Center, Research report, Date accessed: March 2021, https://www.pewresearch.org/social-trends/2021/03/25/the-growing-diversity-of-black-america/PEW

52. V Grech and M Borg, ”Influenza vaccination in the COVID-19 era, Early Human Development”, 2020, ELSEVIER vol 148 DOI:https://doi.org/10.1016/j.earlhumdev.2020.105116,Online https://www.sciencedirect.com/science/article/pii/S0378378220303455

53. Wan Yang, Sharon K. Greene, Eric R. Peterson, Wenhui Li, Robert Mathes, Laura Graf, Ramona Lall, Scott Hughes, Jade Wang, Anne Fine, “Epidemiological characteristics of the B.1.526 SARS-CoV-2 variant”, medRxiv 2021.08.04.21261596; doi: https://doi.org/10.1101/2021.08.04.21261596

54. Coronavirus Resource Center, Harvard Medical School, Health Publishing Online, https://www.health.harvard.edu/diseases-and-conditions/coronavirus-resource-center

55. Leite, H., Gruber, T. and Hodgkinson, I.R. (2020), “Flattening the infection curve – understanding the role of telehealth in managing COVID-19”, Leadership in Health Services, Vol. 33 No. 2, pp. 221–226. https://doi.org/10.1108/LHS-05-2020-084

56. WHO, “Tracking SARS-CoV-2 variants, List of coronavirus variants”, https://www.who.int/en/activities/tracking-SARS-CoV-2-variants/, Accessed:12 Nov 2021.

57. Census 2020, “Historical Population Density Data (1910-2020)”, The United States Census Bureau, http://www.census.gov/data/tables/time-series/dec/density-data-text.html, DOI:October 8, 2021

58. van Staden, C. COVID-19 and the crisis of national development. Nat Hum Behav 4, 443–444 (2020). URL:https://doi.org/10.1038/s41562-020-0852-7.

59. Miller M. 2019 Novel Coronavirus COVID-19 (2019-nCoV) Data Repository: Johns Hopkins University Center for Systems Science and Engineering. Bulletin-Association of Canadian Map Libraries and Archives (ACMLA). 2020 Mar 30(164):47–51.

60. Tyrrell DA, Bynoe ML. Cultivation of viruses from a high proportion of patients with colds. Lancet 1966: 1: 76–77.

61. Dolgin, E. (2021). The tangled history of mRNA vaccines. Nature, 597(7876), 318–324. https://doi.org/10.1038/d41586-021-02483-w

62. U. Şahin et al. Nature Rev. Drug Discov. 13, 759–780 (2014)

63. X. Hou et al. Nature Rev. Mater.https://doi.org/gmjsn5 (2021)

64. Rahimi A, Mirzazadeh A, Tavakolpour S. Genetics and genomics of SARS-CoV-2: A review of the literature with the special focus on genetic diversity and SARS-CoV-2 genome detection. Genomics. 2021 Jan;113(1 Pt 2):1221–1232. doi: 10.1016/j.ygeno.2020.09.059. Epub 2020 Sep 30. PMID: 33007398; PMCID: PMC7525243.

65. “The Covid-19 Pandemic”,The New York Times, https://www.nytimes.com/news-event/coronavirus

66. Dorn, A. V., Cooney, R. E., & Sabin, M. L. (2020). COVID-19 exacerbating inequalities in the US. The Lancet, 395(10232), 1243–1244. https://doi.org/10.1016/s0140-6736(20)30893-x

67. Gaffney, A. W., Himmelstein, D. U., & Woolhandler, S. (2021). Trends and Disparities in Teleworking During the COVID-19 Pandemic in the USA: May 2020–February 2021. Journal of General Internal Medicine, 36(11), 3647–3649. https://doi.org/10.1007/s11606-021-07078-9

68. Cutler D., Deaton A., & Lleras-Muney A. (2006). The Determinants of Mortality. Journal of Economic Perspectives, 20(3), 97–120. https://doi.org/10.1257/jep.20.3.97

69. Chetty, R., Friedman, J.N., Hendren, N., Stepner, M. and The Opportunity Insights Team, 2020. How did COVID-19 and stabilization policies affect spending and employment? A new real-time economic tracker based on private sector data (pp. 1-109). Cambridge, MA: National Bureau of Economic Research.

70. Roser M, Ritchie H, Ortiz-Ospina E. Coronavirus disease (COVID-19). https://ourworldindata.org/coronavirus.

71. Centers for Disease Control and Prevention. Older adults, at greater risk of requiring hospitalization or dying if diagnosed with COVID-19; 2020. https://www.cdc.gov/coronavirus/2019-ncov/need-extra-precautions/older-adults.html.

72. Davies NG, Klepac P, Liu Y, et al. Age-dependent effects in the transmission and control of COVID-19 epidemics. Nat Med. 2020; 26(8): 1205–1211. https://pubmed.ncbi.nlm.nih.gov/32546824/

73. Omori R, Matsuyama R, Nakata Y. The age distribution of mortality from novel coronavirus disease (COVID-19) suggests no large difference of susceptibility by age. Sci Rep. 2020; 10(1): 1–9.

74. Applegate WB, Ouslander JG. COVID-19 presents high risk to older persons; 2020. Wiley Online Library.

75. T. Ford, S. Reber, R. V. Reeves, Race gaps in COVID-19 deaths are even bigger than they appear. Brookings (2020). https://www.brookings.edu/blog/up-front/2020/06/16/race-gaps-in-covid-19-deaths-are-even-bigger-than-they-appear/

76. Lauer SA, Grantz KH, Bi Q, et al. The incubation period of coronavirus disease 2019 (COVID-19) from publicly reported confirmed cases: estimation and application. Ann Intern Med. 2020; 172(9): 577–582.

77. Kandi V, Thungaturthi S, Vadakedath S, Gundu R, Mohapatra RK. Mortality rates of coronavirus disease 2019 (COVID-19) caused by the novel severe acute respiratory syndrome coronavirus-2 (SARS-CoV-2). Cureus. 2021; 13(3):e14081.

78. World Health Organization. Managing epidemics, key facts about major deadly diseases; 2020. http://www.who.int/emergencies/diseases/managing-epidemics/en/. Accessed April 14, 2021.

79. Polyakova, M., Kocks, G., Udalova, V., & Finkelstein, A. (2020). Initial economic damage from the COVID-19 pandemic in the United States is more widespread across ages and geographies than initial mortality impacts. Proceedings of the National Academy of Sciences, 117(45), 27934–27939. https://doi.org/10.1073/pnas.2014279117

80. Coibion, O., Gorodnichenko, Y., & Weber, M. (2020). The Cost of the COVID-19 Crisis: Lockdowns, Macroeconomic Expectations, and Consumer Spending. SSRN Electronic Journal. Published. https://doi.org/10.2139/ssrn.3593469

81. McLaren, J. (2021). Racial Disparity in COVID-19 Deaths: Seeking Economic Roots with Census Data. The B.E. Journal of Economic Analysis & Policy, 21(3), 897–919. https://doi.org/10.1515/bejeap-2020-0371

82. US Census Bureau 2019 National and State Population Estimates. https://www.census.gov/newsroom/press-kits/2019/national-state-estimates.html. Accessed 10 July 2021.

83. WHO, “COVID-19:Global literature on coronavirus disease”, Online database of recent research work on COVID-19,URL:https://search.bvsalud.org/global-literature-on-novel-coronavirus-2019-ncov/.

84. Fidler, F. and Wilcox, J., 2018. Reproducibility of scientific results..

85. Bezanson, J., Edelman, A., Karpinski, S., & Shah, V. B. (2017). Julia: A Fresh Approach to Numerical Computing. SIAM Review, 59(1), 65–98. https://doi.org/10.1137/141000671

86. Bezanson, J., Karpinski, S., Shah, V. B., and Edelman, A. (2012). Julia: A fast dynamic language for technical computing. ArXiv Preprint 1209.5145. download Julia:https://julialang.org/

